# Deletion of *CH25H* and *LIPA* Genes in Human Abolishes Biosynthesis of 25-Hydroxycholesterol but not of 7α,25-Dihydroxysterols and Enhances Non-enzymatic Cholesterol Oxidation: Metabolic Changes are Partially Reversed by Hematopoietic Stem Cell Transplant^‡^

**DOI:** 10.1101/2025.07.29.25330953

**Authors:** Eylan Yutuc, Arunabha Ghosh, Jonas Abdel-Khalik, Anu Goenka, Mohsen Ali Asgari, Gloria Lopez-Castejon, Glenda M Beaman, Robert F Wynn, William G Newman, Simon A Jones, William J Griffiths, Yuqin Wang

## Abstract

The *CH25H* (*cholesterol 25-hyroxylase*) and *LIPA* (*lipase A, lysosomal acid type*) genes are contiguous genes on chromosome 10. *CH25H* is translated to cholesterol 25-hydroxylase which generates 25- hydroxycholesterol (25-HC) from cholesterol, while *LIPA* codes for lysosomal acid lipase (LAL) which hydrolyses cholesteryl esters in the endosome – lysosome compartment. Here we report the effect on the oxysterol pattern in plasma of the homozygous deletion of these two genes and their restoration by allogeneic hematopoietic stem cell transplant (HSCT). In the absence of *CH25H*, 25-HC is not detected in plasma, but surprisingly, 7α,25-dihydroxysterols are present, indicating their formation by a second sterol 25-hydroxylase. As with the isolated deletion of *LIPA* seen in Wolman disease, patients with the homozygous contiguous deletion show high levels of cholesterol autoxidation products in plasma. HSCT of patients with the contiguous deletion restores both 25-HC and autoxidation products to normal levels following an initial burst in autoxidation soon after transplant.

## Introduction

One route for cells to receive cholesterol is via receptor mediated endocytosis (1), where cholesterol esters are delivered to cells in low density lipoprotein (LDL) particles (Supplemental Figure S1). The esters are hydrolysed in the endosome-lysosome compartment by lysosomal acid lipase (LAL) coded by the *LIPA* (*lipase A, lysosomal acid type*) gene. Non-esterified cholesterol is then transferred out of the endosome-lysosome compartment by Niemann-Pick C2 (NPC2) protein working in concert with membrane bound NPC1 protein, and then on to the plasma membrane before being routed to the endoplasmic reticulum (2, 3). When at high levels in the endoplasmic reticulum, non-esterified cholesterol interacts with SCAP (SREBP-cleavage activating protein) to regulate its own synthesis and LDL-receptor (LDLR) expression via repressing activation of SREBP-2 (sterol regulatory-element binding protein-2), the master transcription factor for genes coding enzymes of cholesterol synthesis and of LDLR (4). Cholesterol 25-hydroxylase coded by *CH25H,* the adjacent gene to *LIPA* on chromosome 10, is also involved in the regulation of cholesterol synthesis and LDLR expression. CH25H catalyses the conversion of cholesterol to 25-hydroxycholesterol (25-HC) (5), which can also repress the processing of SREBP-2 to its active form as a master transcription factor (6).

Wolman disease (MIM: 620151) is a rare genetic disorder characterized by a complete absence of LAL enzyme activity or biallelic loss of function of the *LIPA* gene (7, 8). This lack of activity leads to a build-up of cholesterol esters and triglycerides which results in hepatosplenomegaly, while lipid deposition along the gastrointestinal tract leads to thickened bowel walls with resultant malnutrition and wasting. Steatosis may progress to liver failure (9). Interestingly, patients with Wolman disease show high levels of cholestane-3β,5α,6β-triol (3β,5α,6β-triol) in plasma (8, 10–13). 3β,5α,6β-triol is a hydrolysis product of 3β-hydroxycholestan-5,6-epoxide, also called 5,6-epoxycholesterol (5,6-EC), formed by free radical oxidation of non-esterified cholesterol, and normally seen in Niemann-Pick type C disease where non-esterified cholesterol is trapped in the lysosomal compartment (14). Biallelic loss of function variants in *LIPA* will lead to reduced transport of non-esterified cholesterol from the lysosome to the endoplasmic reticulum, ultimately resulting in removal of one of the brakes on synthesis of cholesterol in the endoplasmic reticulum, potentially leading to cholesterol overload and non-enzymatic oxidation of cholesterol (Supplemental Figure S1). In fact elevation of both free and esterified cholesterol has been observed in Wolman disease (15). Besides 3β,5α,6β-triol we have observed its enzymatically generated metabolites i.e., sterol acids, and those of 7-oxocholesterol (7-OC, also called 7-ketocholesterol) and of 7β-hydroxycholesterol (7β-HC), two other autoxidation products of cholesterol, in plasma of patients with Wolman disease (11, 16).

*CH25H* is an interferon (IFN) stimulated gene mostly expressed in activated immune cells in response to viral or bacterial infection (17–20). 25-HC, the product of CH25H hydroxylation of cholesterol, has antiviral activity (20–22) and is also anti-inflammatory (23–25), although pro-inflammatory properties have also been ascribed to this oxysterol (26). 25-HC will repress cholesterol synthesis by restricting processing of SREBP-2 to its active form (6) and enhance cholesterol removal from cells by activating the liver X receptors (LXRs) (27, 28), with the resultant expression of the cholesterol efflux proteins ABCA1 and ABCG1 (29). 25-HC will also enhance cholesterol esterification by acyl-CoA cholesterol acyl transferase (ACAT also known as sterol O-acyltransferase SOAT) (30). In combination, 25-HC will have the effect of reducing the cellular content of unesterified cholesterol. We are, however, not aware of a specific CH25H-deficiency phenotype, as no cases of isolated CH25H deficiency have been reported and we are unable to clinically differentiate the children with the combined deficiency from isolated disease cases. That does not, however, mean there is no CH25H phenotype.

Here we report the effects on the plasma oxysterol profile of deletion of both *CH25H* and *LIPA* genes in three children. Like children with only LAL deficiency (Wolman disease), the oxysterol profiles of the three children with *CH25H - LIPA* homozygous deletions show elevation in 3β,5α,6β-triol, 7-OC and 7β-HC and their onwards metabolites towards bile acids. As expected, 25-HC was absent from plasma of these children. However, following allogenic hematopoietic stem cell transplant (HSCT) 25-HC was detected in plasma, and as time after transplant progressed the levels of 3β,5α,6β-triol, 7-OC and 7β-HC and of their metabolites fell towards normality. A point of interest is that 7α,25-dihydroxysterols were present in plasma of the children where *CH25H* is deleted before and after HSCT, indicating that 7α,25-dihydroxycholesterol (7α,25-diHC) can be formed in the absence of CH25H, presumably by cytochrome P450 (CYP) 3A4 (31, 32). This has relevance to the presumed biological activity of 7α,25-diHC based on the extrapolation of data from experiments where *Ch25h* is deleted (24, 33–35).

## Material and Methods

### Patient Samples

Patients were all treated at Royal Manchester Children Hospital and had confirmation of diagnosis of Wolman disease by demonstrating a deficiency of LAL activity in leukocytes.

Genome-Wide SNP Microarray analysis using the Affymetrix Genome-Wide SNP6.0 microarray (Santa Clara, CA, USA) was undertaken for affected individuals on DNA extracted from lymphocytes. Copy number data was generated using the SNP 6.0 CN/LOH Algorithm within the Affymetrix Genotyping console v4.2 and visualised using Chromosome Analysis suite v2.1. Autozygosity mapping was conducted using autoSNPa software (36). The region flanking the deleted region was amplified by PCR using primers, followed by Sanger sequencing to confirm the breakpoints.

The three patients received both sebelipase alfa (enzyme replacement therapy, ERT) and dietary substrate reduction (DSR) prior to HSCT. Data was gathered retrospectively from notes and electronic records and to the best of our knowledge none of the patients were related. Patient details are listed in Table 1. Parents were heterozygotes for the deletion of both genes.

**Table 1.** Data for patients presenting with the *LIPA – CH25H* homozygous deletion.

| Anonymised Code | Patient 08-002 | Patient 08-006 | Patient 08-008 |
| --- | --- | --- | --- |
| Sex | Female | Male | Male |
| Age at HSCT (years) | <5 | <5 | <5 |
| Pathogenic variant | Homozygous deletions of *CH25H* and *LIPA* | Homozygous deletions of *CH25H* and *LIPA* | Homozygous deletions of *CH25H* and *LIPA* |
| Peripheral blood chimerism | 25% stable (4.5 years post-HSCT) | 30% stable (3 years post-HSCT) | 100% stable (4 years post-HSCT) |
| ERT dose post-HSCT | Reduced significantly post-transplantation | Reduced significantly post-transplantation | Reduced slightly post-transplantation |
| GI tract symptoms post-HSCT | Significant improvement | Significant improvement |  |
| BCG status | BCG site abscess at presentation. After HSCT developed suppurative BCG lymphadenitis | Post-HSCT BCG site erythema temporally associated with other vaccinations | BCG site abscess at presentation |

ERT dose was based upon clinical need. During the pre-HSCT period doses ranged from 3 mg/kg to 7.5 mg/kg once weekly, delivered as an intravenous infusion, usually via a central venous catheter. Post-HSCT, the frequency and dose of ERT was reduced. HSCT, including donor source and conditioning treatment, was allocated as the best option available for the patient at the time of HSCT. Conditioning involved immunosuppression and myeloablation and was modified according to the patient’s individual clinical condition and the donor source.

All participants or their parents/guardians provided informed consent and the study was performed with institutional review board approval (REC08/H1010/63) and adhered to the principles of the Declaration of Helsinki.

### Materials

Authentic standards were from Avanti Polar Lipids (Alabaster, AL, USA) or a kind gift from Professor Jan Sjövall at Karolinska Institutet, Stockholm. Details of all other materials used can be found in references (11, 16, 37).

### Extraction and Derivatisation

The analytical method for oxysterol analysis is described in detail in reference (37). In brief, oxysterols were extracted from plasma (100 μL) by single phase extraction into absolute ethanol (1.05 mL) containing isotope labelled internal standards. After dilution to 70% ethanol and centrifugation, the supernatant was subjected to solid phase extraction (SPE) on a C_18_ column (200 mg, Certified Sep-Pak C_18_, Waters Corp). Oxysterols elute in 70% ethanol (SPE1-Fr1, 7 mL), and after a wash with 70% ethanol (4 mL), cholesterol and similarly hydrophobic sterols elute in absolute ethanol (2 mL) to give SPE1-Fr3.

To maximise sensitivity for subsequent mass spectrometry analysis we employed a derivatisation strategy (see Supplemental Figure S2). The oxysterol fraction (SPE1-Fr1) was divided into two equal subfractions (SPE1-Fr1A and SPE1-Fr1B) which were dried under vacuum and reconstituted in 100 μL of propan-2-ol. To fraction A, 1 mL of 50 mM KH_2_PO_4_ (pH 7) was added containing cholesterol oxidase enzyme from *Streptomyces sp* (0.264 units, Merck, UK). The reaction mixture was incubated at 37 °C for 1 hr and quenched with 2 mL of methanol. 150 μL of glacial acetic acid was then added followed by 190 mg of [^2^H_5_]Girard P (GP) hydrazine (bromide salt) reagent. The reaction was allowed to proceed at room temperature overnight in the dark. An identical procedure was followed for the derivatisation of fraction B except the cholesterol oxidase enzyme was omitted and 150 mg of [^2^H_0_]GP hydrazine (chloride salt) replaced [^2^H_5_]GP. In this way oxysterols with a 3β-hydroxy-5-ene native structure become uniquely derivatised with [^2^H_5_]GP (Fraction A), while those with a 3-oxo group in their native structure become derivatised with [^2^H_5_] (in Fraction A) and to an equal extent with [^2^H_0_]GP (in Fraction B). To remove excess derivatisation reagent a re-cycling procedure was performed on an OASIS HLB column (60 mg, Waters Corp) where the reaction mixture was added to the column in 70% organic, the eluent diluted to 35% organic and recycled on the column. The next eluent was diluted to 17.5% organic and re-cycled on the column. At this point all derivatised oxysterols are retained on the column, which was then washed with 10% methanol. Oxysterols were then eluted with 3 x 1 mL of methanol. Oxysterols elute in the first 2 mL. SPE1-Fr3 was treated in an identical fashion to SPE1-Fr1 but cholesterol and its precursors elute across the 3 mL of methanol.

### LC-MS(MS^n^)

Just before analysis, derivatised oxysterols were diluted to 60% methanol then injected onto a liquid chromatography – mass spectrometry (LC-MS) system. This LC system consisted of a reversed phase Hypersil Gold C_18_ column (50 x 2.1 mm, 1.9 μm particles) and an Ultimate 3000 UHPLC (Thermo Fisher Scientific, UK). A gradient of methanol, acetonitrile and 0.1% formic acid was employed. MS analysis was on a Orbitrap Elite or Orbitrap IDX (both Thermo Fisher Scientific). Ionisation was by positive-ion electrospray, high resolution mass spectra (120,000 at *m/z* 400) were recorded in the Orbitrap, while multi-stage fragmentation (MS^3^) spectra were recorded in the linear ion-trap. Quantification was by isotope-dilution mass spectrometry (37).

Note, in this procedure a hydrolysis step was not carried out so concentrations measured are for the non-esterified molecules.

## Results and Discussion

In all three affected but unrelated children, each born to consanguineous parents with ancestry in the Indian subcontinent, copy number analysis by SNP microarray defined a 112kb homozygous deletion completely encompassing *LIPA* and the adjacent gene, *CH25H*. Identical breakpoints (chr10:90,947,449_91,060,391del) were demonstrated by PCR and sequencing, which did not affect the flanking genes *FAS* and *IFIT2* (Figure 1). Genotyping confirmed that the parents of each of the children were heterozygous for the deletion. Autozygosity analysis of SNP array data from an affected individual in each family was performed which determined that the deletion occurred within a shared haplotype spanning 6.1Mb, supporting the presence of a founder event. The genomic databases DECIPHER, HGMD, ClinVar and gnomAD did not reveal reports of similar deletions of *LIPA* and deletions of this region are absent in a database of 500 ethnically matched individuals.

**Figure 1.**
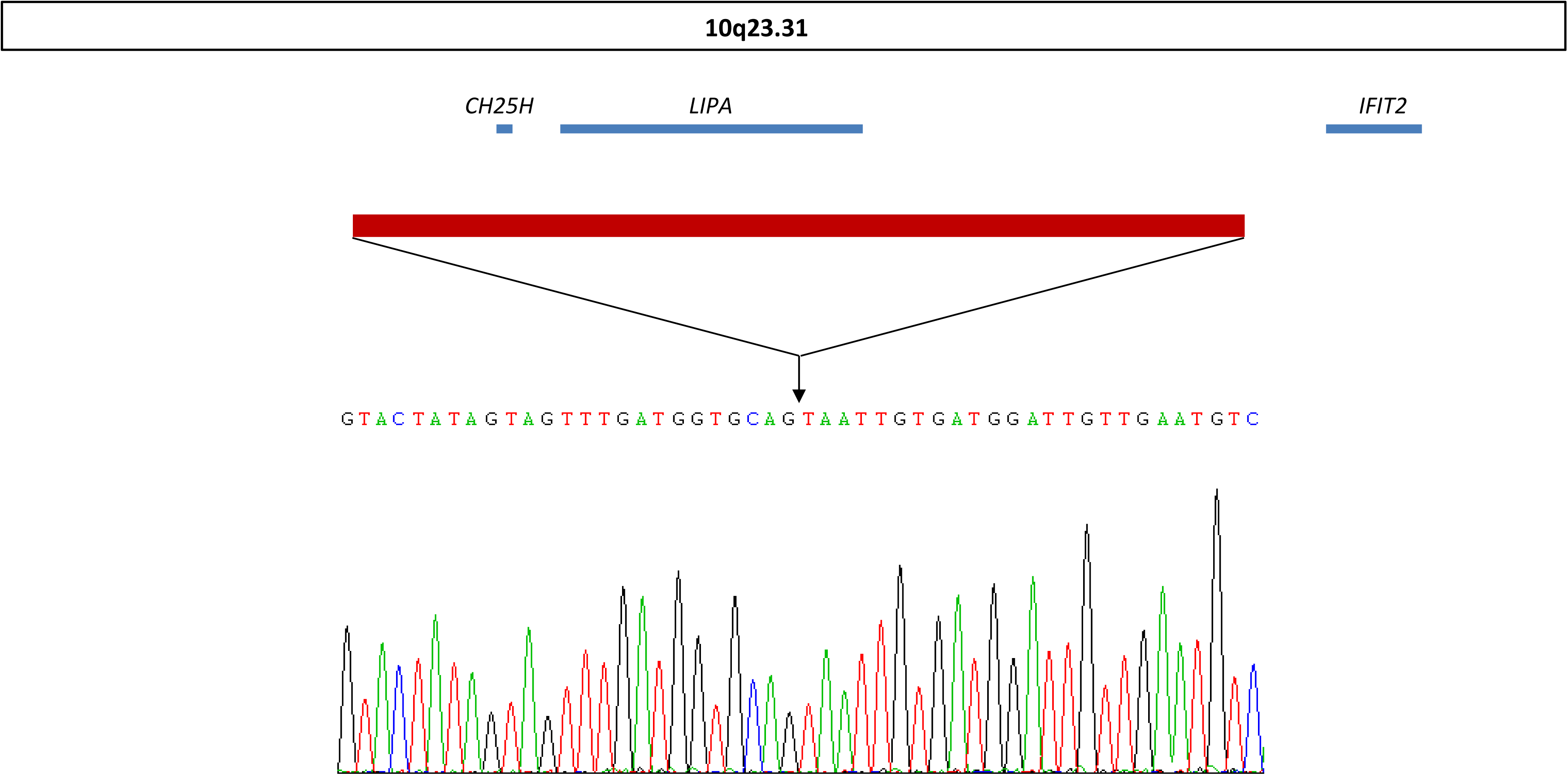
Deletion of *CH25H* and *LIPA* on chromosome 10.

We have previously utilised GP derivatisation and LC-MS with multi-stage fragmentation (MS^3^) for the analysis of oxysterols from patients with Wolman disease (11, 16). The method adds sensitivity and specificity to the analysis. Importantly, once derivatised with GP reagent oxysterols give structurally specific MS^3^ spectra, which when combined with accurate mass information (<5 ppm) and chromatographic retention time give confident identifications, even in the absence of authentic standards (37).

### Oxysterols Identified in Plasma

The oxysterols identified in plasma in the present study are listed in Supplemental Table S1. Identifications were based on reference to authentic standards, where this was not the case, it is stated in supplemental Table S1. Oxysterol in plasma represent products of cholesterol metabolism that are synthesised by different cells including white blood cells and cells in the liver, brain and lung, which enter the circulation and are transported (back) to the liver for further metabolism (38–43). Their analysis provides a snapshot of cholesterol metabolism. Oxysterols found in plasma can be divided into different groups according to the metabolic pathways in which they participate. In the healthy individual the dominant pathways are the acidic and neutral pathways of bile acid biosynthesis, while minor pathways start with 25-HC and 24S-hydroxycholesterol (24S-HC) (Supplemental Figure S3A, B, D) (44–46). However, in patients with Smith-Lemli-Opitz syndrome (SLOS), Niemann Pick diseases type C (NPC) and type B (NPB also known as acid sphingomyelinase deficiency), Wolman disease, and as seen in Supplemental Table S1 in the patients with the *CH25H* – *LIPA* homozygous deletion, metabolites from additional pathways become prominent (11, 16, 47, 48). In patients with SLOS metabolites in pathways originating from 7β-HC and 7-OC are evident (48), while in patients with Niemann Pick diseases or Wolman disease metabolites originating from 3β,5α,6β-triol are additionally observed (11, 16, 49, 50) (see Supplemental Figure S3C).

### 25-Hydroxylase Pathway

The plasma concentration of 25-HC is usually low, about 1 ng/mL for the non-esterified molecule (37, 51), but can be elevated following bacterial or viral infection (17, 18, 52). 25-HC is biosynthesised from cholesterol in activated macrophages in a reaction catalysed by the enzyme CH25H (Supplemental Figure S3A, metabolite colour teal). Recent evidence suggests that 25-HC is predominantly generated in macrophages from cholesterol taken up by receptor mediated endocytosis, or perhaps by scavenger receptors, rather than from newly synthesised cholesterol (Supplemental Figure S1) (53). When plasma was analysed from the patients with the *CH25H* – *LIPA* homozygous deletion, 25-HC was undetected (Figure 2A-C, Supplemental Table S1). However, following HSCT the level of 25-HC in each of the patients rose to a value (>5 ng/mL) greater than in their parents (1 ng/mL), but as time progressed this value fell to a value similar to that of their parents (Figure 3). Some of this fall in 25-HC concentration can be ascribed to mixed chimerism in two of the three patients (Table 1). It is notable that for each of the patients, the concentration of 25-HC maximises at the first time point post HSCT, then falls away over time.

**Figure 2.**
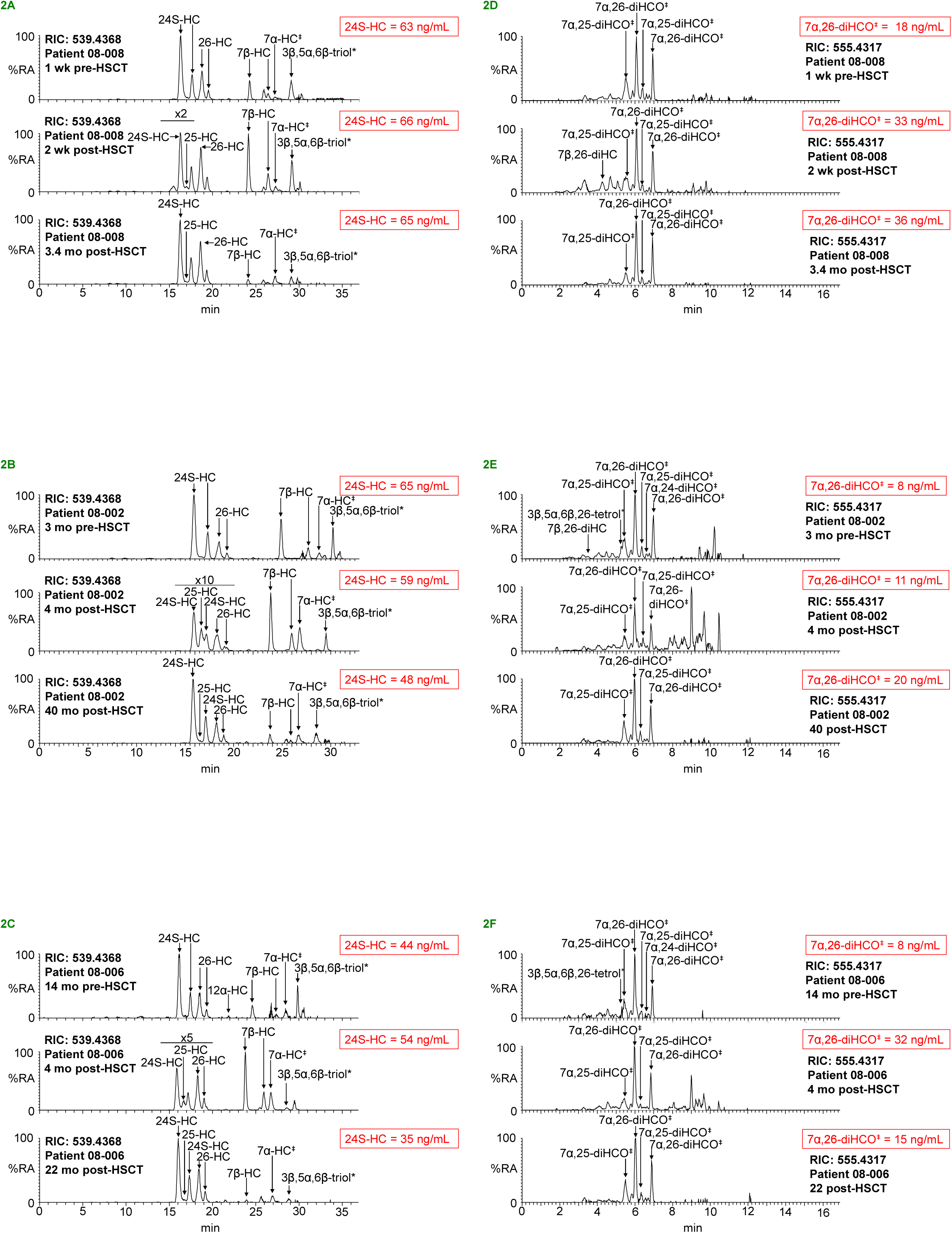
25-HC is absent in plasma from patients with the *CH25H – LIPA* homozygous deletion prior to HSCT but is present following transplant. 7α,25-Dihydroxysterols are present in these patients before and after HSCT. Reconstructed ion chromatograms (RICs) (539.4368 ± 5 ppm) for the [M]^+^ ion of monohydroxycholesterols for the three patients, before (top), immediately after (centre) and at the last time point (bottom). 3β,5α,6β-triol* is observed as the [M-H_2_O]^+^ ion. The peaks labelled 7α-HC^‡^ correspond to the 7α-HC and 7α-HCO combined. (A) Patient 08-008, (B) 08-002 and (C) 08-006.

**Figure 3.**
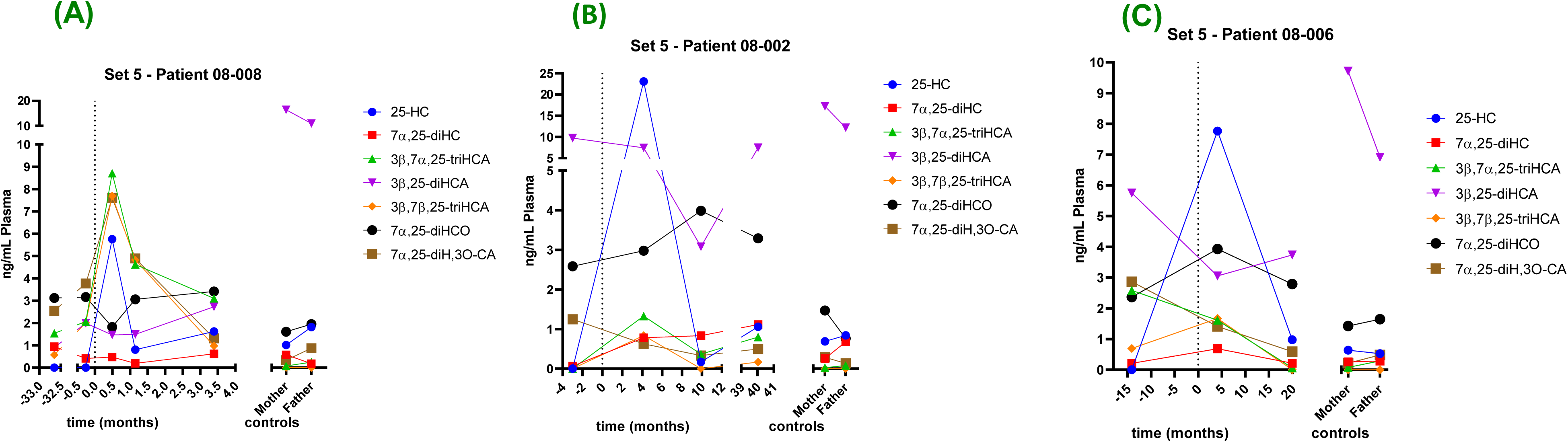
Variation of the concentrations of 25-HC, 7α,25-diHC and their metabolites before and after HSCT. (A) Patient 08-008, (B) patient 08-002 and (C) patient 08-006. Note that the axis have been split to aid visualisation. The time of HSCT (time zero) is indicated by a dashed line.

RIC (555.4317 ± 5 ppm) for the [M]^+^ ion of dihydroxysterols for the three patients before (top), immediately after (centre) and at the last time point (bottom). (D) Patient 08-008, (E) 08-002 and (F) 08-006. 3β,5α,6β,26-tetrol* is observed as the [M-H_2_O]^+^ ion. The peaks labelled 7α,25-diHCO^‡^ correspond to the 7α,25-diHC and 7α,25-diHCO combined. 7α,24-diHCO^‡^ and 7α,26-diHCO^‡^ represent similar combinations. Note, the chromatograms shown in (A), (B) and (C) were recorded with a 37 min gradient, those shown in (D), (E) and (F) with a 17 min gradient. There is some variation in retention time, as data was recorded over a time scale of years. For comparison, concentrations of selected metabolites are shown in red. Chromatogram in (A), (B) and (C) are aligned to 24S-HC. MRM-like chromatograms are shown in Supplemental Figure S5 and MS^3^ spectra in Supplemental Figure S6. Note, as a consequence of the derivatisation reaction oxysterols give cis and trans isomers about the CN double bond (see Supplemental Figure S2C) resulting in most cases in two chromatographic peaks. Abbreviation of different oxysterols are defined in Supplemental Table S1.

7α,25-diHC is a minor oxysterol in plasma, the concentration of the non-esterified molecule is usually around 0.5 ng/mL (37). 7α,25-diHC has attracted considerable interest in recent years as a mediator in the adaptive immune system acting as a ligand to GPR183 (Epstein-Barr virus induced gene 2, EBI2) (24, 33, 34). 7α,25-diHC is a substrate for hydroxysteroid dehydrogenase 3B7 (HSD3B7) being converted to inactive 7α,25-dihydroxycholest-4-en-3-one (7α,25-diHCO, Supplemental Figure S3A), which is more abundant in plasma (1 – 2 ng/mL) (37). 7α,25-diHC is usually considered to be formed from 25-HC by the action of CYP7B1 (54–56). However, there is not a complete absence of 7α,25-diHC in people with CYP7B1 deficiency suggesting an alternative route to its formation. An alternative sterol 25-hydroxylase to CH25H is CYP3A4 which can 25-hydroxylate both cholesterol and 7α-hydroxycholesterol (7α-HC) (31, 32). In the current study the patients with the *CH25H – LIPA* homozygous deletion still show the presence of 7α,25-diHC and 7α,25-diHCO in their plasma even before HSCT (Figure 2D-F and Supplemental Table S1) at levels similar to those of their parents (Figure 3). 7α,25-diHC and 7α,25-diHCO can be further metabolised to the corresponding 3β,7α,25-trihydroxycholest-5-en-26-oic (3β,7α,25-triHCA) and 7α,25-dihydroxy-3-oxocholest-4-en-26-oic acids (7α,25-diH,3O-CA, Supplemental Figure S3A) (46). At least one of these two acids was detected in plasma from each of the patient prior to transplant, indicating that CH25H is not essential for their synthesis. However, from the time-course data (Figure 3) the elevation in 3β,7α,25-triHCA soon after transplant in two of the patients suggests that it can also be formed via a pathway involving CH25H. A further 25-hydroxy metabolite assigned to the structure 3β,25-dihydroxycholest-5-en-26-oic acid (3β,25-diHCA) based on accurate mass measurement and MS^3^ spectra was found in plasma of all three patients prior to HSCT, thus, assuming the identification is correct, it must be formed independent of *CH25H*, potentially from 3β-hydroxycholest-5-en-26-oic acid in a reaction catalysed by CYP3A4.

### 24S-Hydroxylase Pathway

*Cholesterol 24S-hydroxylase* (*CYP46A1*) is almost exclusively expressed in neurons (57). It will catalyse the 24S-hydroxylation of cholesterol and 24S,24-epoxidation of desmosterol (58, 59). 24S-HC is the major cholesterol metabolite produced in brain and can pass the blood brain barrier to exit the brain and enter the circulation. The concentration of 24S-HC in blood is dependent on the relative size of the brain and liver, and is higher in children than adults (39). Interesting in each of the patients, the levels of 24S-HC in plasma are quite similar before and after HSCT but are higher than for their parents (Supplemental Figure S7). It noteworthy that 24S-HC is synthesised in brain from cholesterol which is *de novo* synthesised in brain and is almost exclusively non-esterified (60), and thus its concentrations as a CYP46A1 substrate is independent of *LIPA* expression, explaining the similar concentration of plasma 24S-HC before and after HSCT. The concentration of 24S,25-epoxycholesterol (24S,25-EC) is about a factor of 10 to 20 lower than that of 24S-HC and like 24S-HC is more abundant in the patient samples than their parents.

### Pathways Initiated by Non-Enzymatic Oxidation of cholesterol

For each of the patients, prior to HSCT most of the metabolites belonging to the 3β,5α,6β-triol, 7β-HC and 7-OC pathways (Supplemental Table S1, see also Supplemental Figure S3C for metabolic pathways) are more than an order of magnitude higher than for their parents, and we see a maintained elevated level, or further increase, in metabolite concentrations in these pathways at the first time point following HSCT. However, as time progressed the metabolite concentrations in each of these pathways fell towards normality (Figure 4).

**Figure 4.**
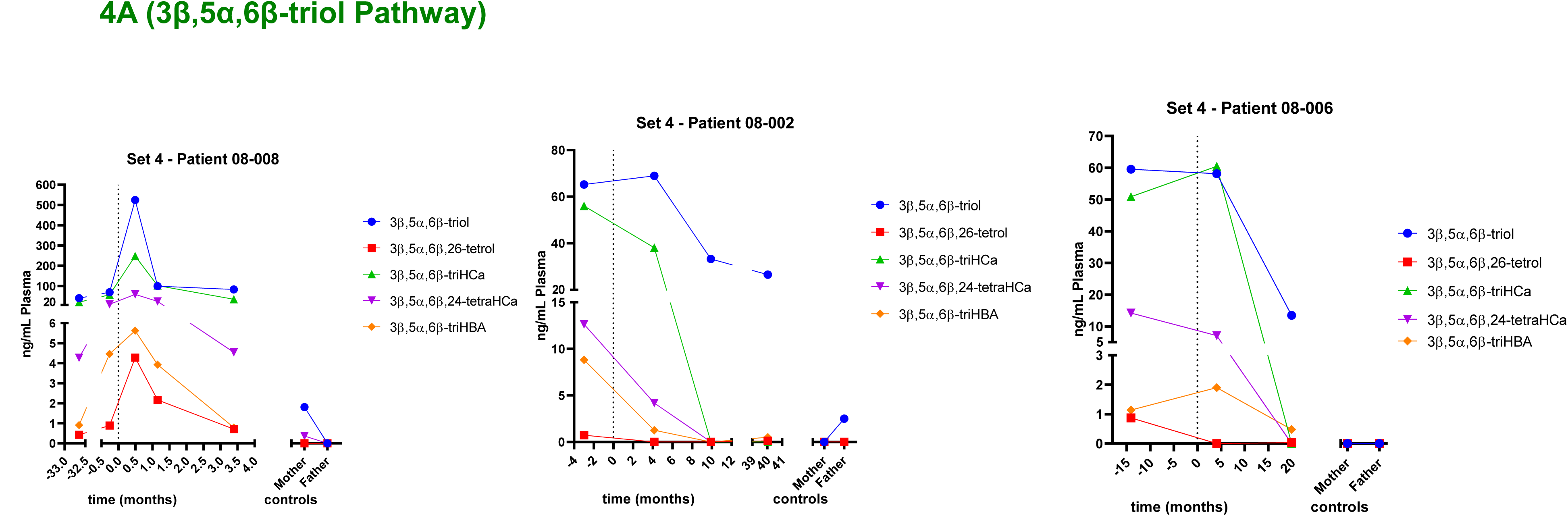
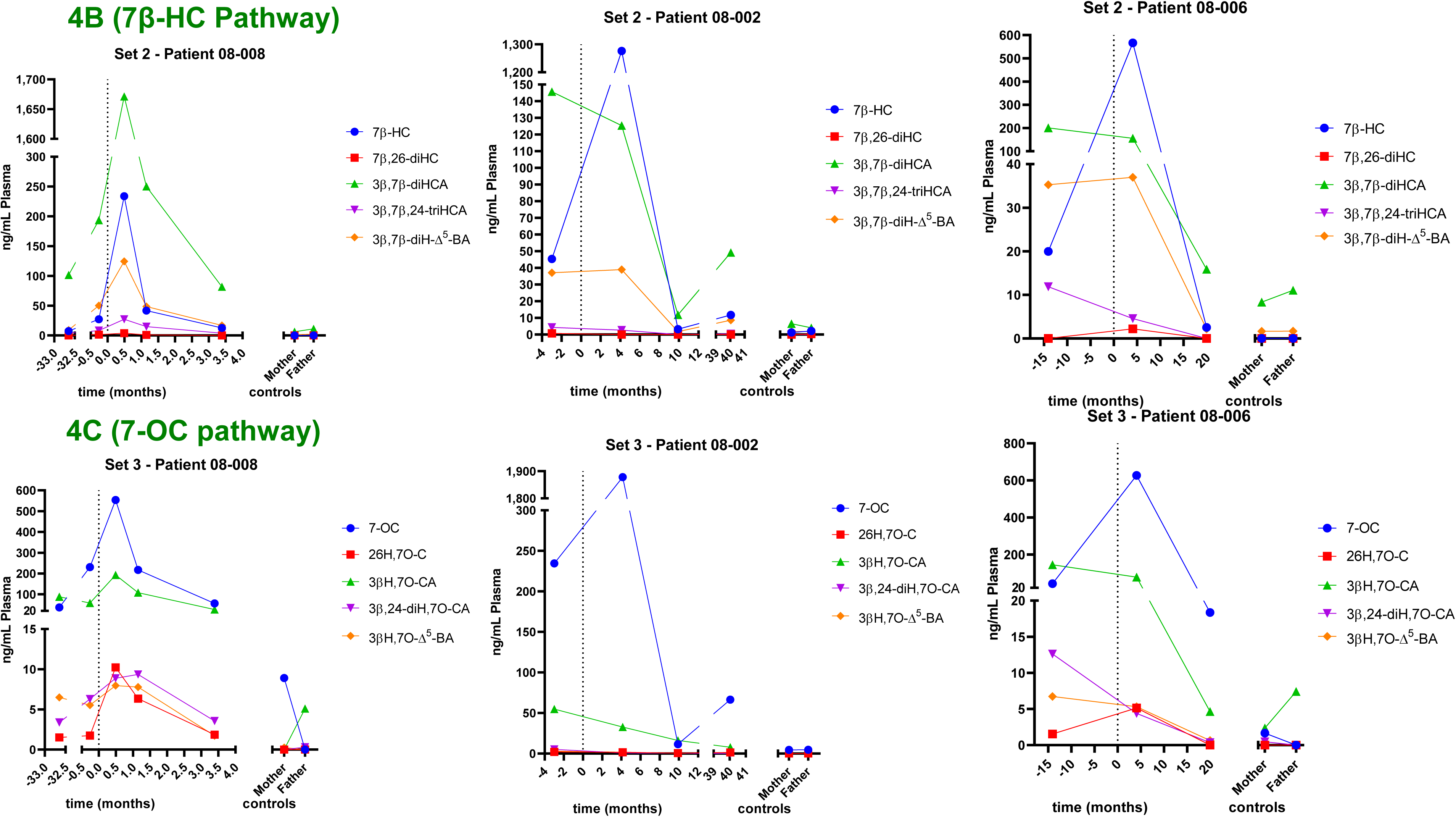
Variation of the concentrations of (A) 3β,5α,6β-triol, (B) 7β-HC and (C) 7-OC and their metabolites before and after HSCT. Data for patient 08-008 is shown in the left hand panels, for patient 08-002 in the central panels and for patient 08-006 in the right hand panels. Note that the axis have been split to aid visualisation. The time of HSCT (time zero) is indicated by a dashed line.

A complication when investigating oxysterols formed endogenously by non-enzymatic reactions, such as the early metabolites in the 3β,5α,6β-triol, 7β-HC and 7-OC pathways is that they can also be formed exogenously as artefacts of sample storage. With the exclusion of these three compounds, the dominant metabolites prior to HSCT and at the first time point(s) after HSCT are the C_27_ sterol acids 3β,5α,6β-trihydroxycholestan-26-oic acid (3β,5α,6β-triHCa), 3β,7β-dihydroxycholest-5-en-26-oic acid (3β,7β-diHCA) and 3β-hydroxy-7-oxocholest-5-en-26-oic acid (3βH,7O-CA). The LC-MS(MS^3^) data illustrating the variation of these acids with time pre-and post-HSCT is shown in Figure 5.

**Figure 5.**
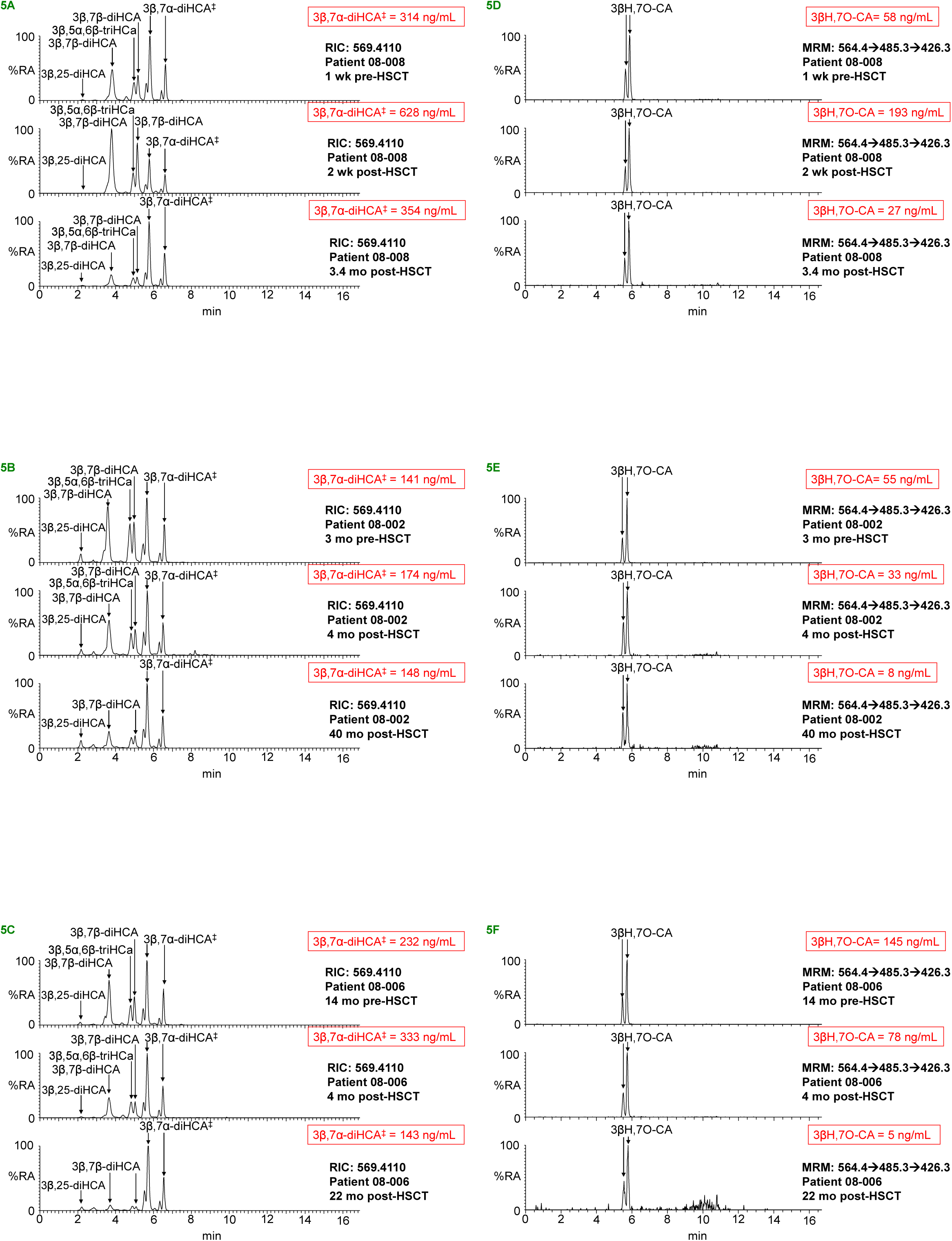
The sterol acids 3β,5α,6β-triHCa, 3β,7β-diHCA and 3βH,7O-CA are elevated in plasma from patients with the *CH25H – LIPA* homozygous deletion collected prior to, and at the first time point after HSCT. RIC *m/z* 569.4110 ± 5 ppm targeting 3β,5α,6β-triHCa and 3β,7β-diHCA for the three patients, before (top), immediately after (centre) and at the last time point (bottom). 3β,7β-diHCA is observed as the [M]^+^ ion of the [^2^H_5_]GP-derivative, while 3β,5α,6β-triHCa* is observed as the [M-H_2_O]^+^ ion of the [^2^H_5_]GP-derivative (16). (A) Patient 08-008, (B) 08-002 and (C) 08-006. MRM – like chromatograms targeting 3βH,7O-CA (564.4→485.3→426.3) (16) for the three patients, before (top), immediately after (centre) and at the last time point (bottom). (D) Patient 08-008, (E) 08-002 and (F) 08-006. The RIC *m/z* 569.4110 ± 5 ppm also shows the presence of 3β,7α-diHCA, 7αH,3O-CA (measured in combination and indicated by 3β,7α-diHCA^‡^) and 3β,25-diHCA. 3β,7α-diHCA and 7αH,3O-CA exist as 25R-and 25S-epimers, the 25S-epimers eluting just before the 25R-epimers, this is also true for 3β,7β-diHCA and 3βH,7O-CA. While R-and S-epimers are resolved for 3β,7α-diHCA, 7αH,3O-CA and 3βH,7O-CA, the S-epimer appears a leading edge on the 3β,7β-diHCA peak. Only cholestenoic acids with a 3β,7α-dihydroxy-5-ene structure are substrates for HSD3B7 and are converted in *vivo* into the 3-ones. For comparison concentrations of selected metabolites are shown in red. Shown in Supplemental Figure S8 are MS^3^ spectra of the identified oxysterols.

### Acidic and Neutral Pathways of Bile Acid Biosynthesis

Metabolites from both the neutral and acidic pathways of bile acid biosynthesis are observed in plasma. There are multiple branches to these pathways and metabolites can cross over from one pathway to another at the levels beyond 26-hydroxycholesterol (26-HC, also known by the non-systematic name 27-hydroxycholesterol (61), see Supplemental Figure S3A). As was the case with the pathways initiated by non-enzymatic oxidation, metabolites of the acidic pathway are dominated by a C_27_ sterol acid, in this case 3β-hydroxycholest-5-en-(25R)26-oic acid (3β-HCA) (Supplemental Table S1), and are elevated before HSCT and at the first time point post transplantation, and then drop to levels quite similar to those of the parents (Figure 6A). 3β,7α-Dihydroxycholest-5-en-26-oic (3β,7α-diHCA, Figure 6A), a major 7α-hydroxylated sterol found in plasma of healthy adults (37, 44), is elevated in each patient pre-HSCT and at the first time point following transplant to more than double the parent concentrations, then falls to levels similar to those of the parents over time. This is not the case with 7α-hydroxy-3-oxocholest-4-en-26-oic (7αH,3O-CA, Figure 6B) which before and after transplant is at levels similar to those of the parents. Some insight into this data comes from the work of Meaney et al which showed that in plasma 3β,7α-diHCA originates predominantly from 3β-HCA of the acidic pathway, while 7αH,3O-CA mostly originates from 7α-HC and 7α-hydroxycholest-4-en-3-one (7α-HCO) of the neutral pathway initiated in the liver (see Supplemental Figure S3A)(43).

**Figure 6.**
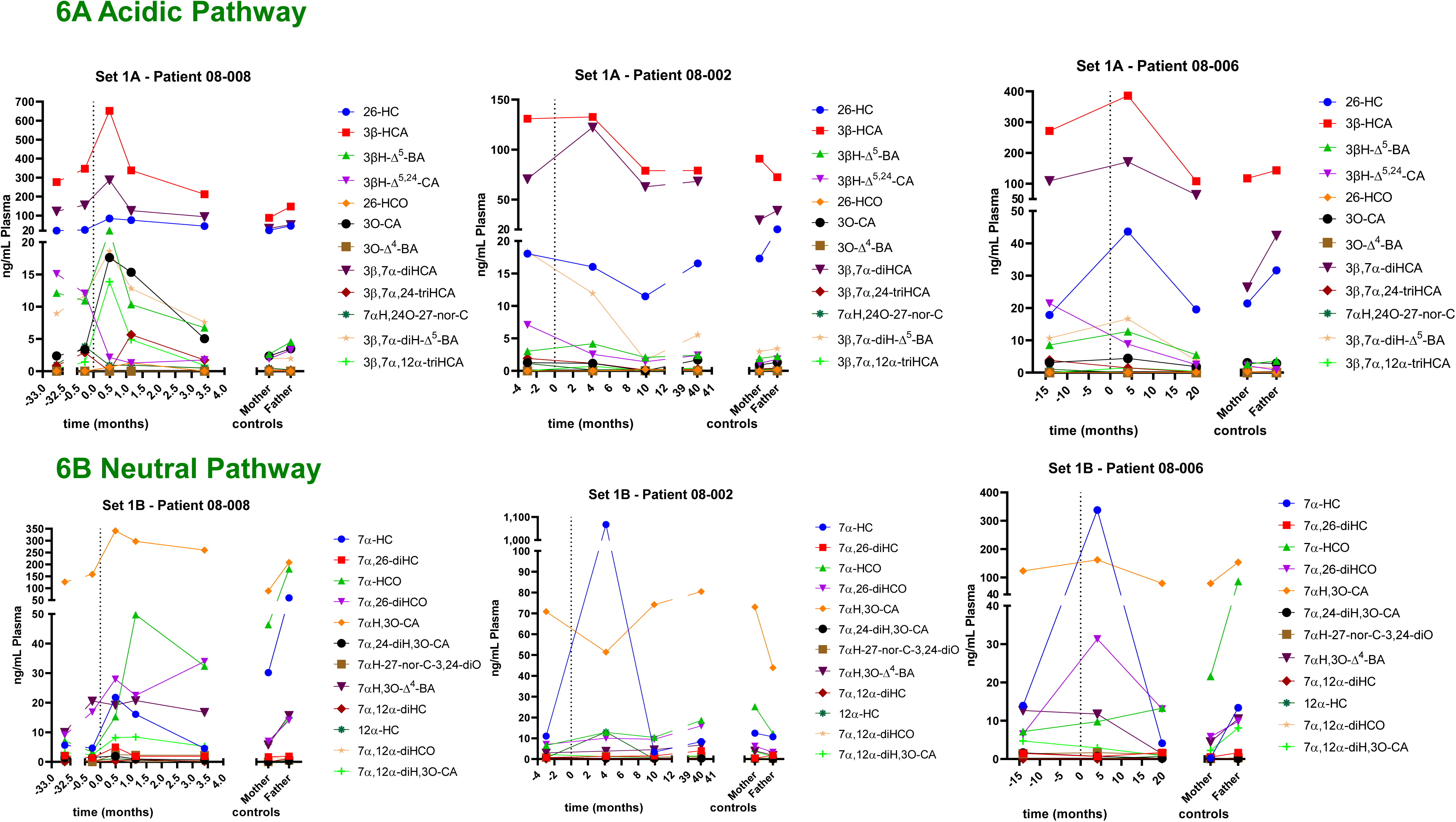
Variation of the concentrations of (A) 3β-HCA and metabolites in the acidic pathway and of (B) 7α-HC, 7α-HCO, 7αH,3O-CA and metabolites originating from the neutral pathway, before and after HSCT. Data for patient 08-008 is shown in the left-hand panel, for patient 08-002 in the centre panel and for patient 08-006 in the right hand panel. The axis have been split to aid visualisation. Note, 3β,7α-diHCA, 7αH,3O-CA, 3β,7α,12α-triHCA, 7α,12α-diH,3O-CA are present in plasma as both 25R and 25S-epimers (37), where the 25R-epimer is dominant, but here for simplicity the two epimers are measured together. The time of HSCT (time zero) is indicated by a dashed line.

In combination, the sterol acid data indicates that pre-HSCT and at the first time point post-HSCT there is increased biosynthesis of the cholestenoic acids which are synthesised, probably extrahepatically, following an initial non-enzymatic oxidation of cholesterol i.e., 3β,5α,6β-triHCa from 3β,5α,6β-triol (Figure 4A); 3β,7β-diHCA from 7β-HC (Figure 4B); and 3βH,7O-CA from 7-OC (Figure 4C), or enzymatically via extrahepatic oxidation of cholesterol utilising CYP27A1 to generate 3β-HCA and on to 3β,7α-diHCA and 3β,7α,12α-trihydroxycholest-5-en-26-oic acid (3β,7α,12α-triHCA) (Figures 6A). Alternatively, the increased plasma content of 3β-HCA and 3β,7α-diHCA could be explained by a defective uptake of these acids by the liver from the circulation. However, the fact that 7αH,3O-CA originating from 7α-HCO in the hepatic neutral pathway is not consistently elevated pre-HSCT and at the first time point post-HSCT, compared to concentrations in parents, indicates that there is no defect in the net uptake of metabolites at these time points. In fact, the data for brain-derived 24S-HC (Supplemental Figure S7) where the plasma concentration before and after HSCT does not substantially change also argues against a defect in liver uptake of sterols in the patients.

In comparison to the above sterol acids the time course data for 25-HC follows a different pattern, this is not surprising as the *CH25H* gene is deleted before transplant yet expressed following HSCT. Prior to HSCT, 25-HC is absent from plasma, then at the first time point post-HSCT reaches a maximum and then falls to values similar to those of the parents (Figure 3). In contrast, 7α,25-diHCO the dominant 7α,25-dihydroxysterol follows a pattern where its levels are quite similar before and after HSCT and to the concentrations found in plasma of the parents.

### Cholesterol and its Precursors

Finally, we investigated how the plasma concentrations of non-esterified cholesterol and its immediate precursors, desmosterol (24-DHC) and 7-dehydrocholesterol (7-DHC, which found in plasma with its isomerisation product 8-dehydrocholesterol, 8-DHC), varied in the three patients before and after HSCT, and compared to their parents. In each of the patients the cholesterol concentration reaches a maximum soon after HSCT and drops to a level similar to that of the parents over time (Supplemental Figure S9). Following HSCT, when donor leukocytes become engrafted into the host bone marrow, donor derived leukocytes populate the peripheral circulation, express *LIPA* and LAL, which can then be taken up by host cells through receptor mediated endocytosis and delivered to the lysosome (8). The initial increase in non-esterified cholesterol following HSCT can thus be explained by the increased availability to cells of LAL, catalysing cholesterol release from its esters, and cholesterol export from cells by ABC-transporters onto circulating lipoproteins. In contrast, the concentration of cholesterol precursors, particularly desmosterol, fall soon after transplant (Supplemental Figure S9). This is explained by LAL released non-esterified cholesterol supressing its own synthesis via inhibition of the SREBP pathway (Supplemental Figure S1) (4). A decrease in cholesterol synthesis can explain the subsequent fall in plasma cholesterol levels. It is noticeable that the cholesterol levels in patient 08-002 first rose slightly following HSCT then fell, but later increased again. The latter increase in cholesterol coincides with an increase in cholesterol precursors which could be a consequence of a fall to 25% chimerism and the resulting reduced availability of non-esterified cholesterol in the endoplasmic reticulum to restrict SREBP-2 processing and cholesterol synthesis, with the resultant increase in its synthesis and that of its precursors.

### BCG Site Abscesses

Bacillus Calmette–Guérin (BCG) vaccine primarily used against tuberculosis (TB) is given to infants soon after birth. The vaccine is a live attenuated strain of *Mycobacterium bovis* which causes TB in cattle. Interestingly, two (08-002 and 08-008) of the three patients studied here had BCG site abscess at the time of presentation and the third patient (08-006) had post-HSCT BCG injection site erythema temporally associated with other vaccinations. In contrast, no BCG abscesses were reported in a group of eight children with only *LIPA* deficiency (62). As discussed above, the oxysterol profiling of plasma from the patients with the homozygous deletion of *CH25H* and *LIPA* showed an absence prior to HSCT of 25-HC which is normally present in plasma at a low level but is enhanced in response to bacterial infection (17, 18). However, 25-HC was present in plasma post-HSCT and also in the heterozygote parents. 25-HC itself is an anti-inflammatory oxysterol down-regulating inflammasome activity and IL-1β production, via a mechanism thought to involve 25-HC restraining cholesterol synthesis required for inflammasome activation (Supplemental Figure S1) (25). It is possible that an absence of 25-HC prior to HSCT results in removal of a metabolic brake on the inflammatory process resulting in the presentation of the BCG site abscess (25). The patient who developed post-HSCT injection site erythema had mixed chimerism at around 30% which could contribute to a reduced metabolic brake applied by 25-HC to the inflammatory process and to post BCG erythema. In fact, there is evidence that LAL inhibition can accentuate inflammasome activity (53), suggesting that a combination of deficient enzyme activity of both LAL and CH25H in combination heightens inflammation in response to BCG. A second explanation for BCG site abscess in the three patients is an absence of the anti-bacterial protection provided by 25-HC (63–65). 25-HC has been shown to reduce “accessible” cholesterol from the plasma membrane of cells, this can reduce the cell-to cell spread of bacteria and also attenuate the effects of bacterial toxins providing an innate immunity mechanism to counter bacteria. In support of this explanation, genetic deficiencies in either IFN-γ or its receptor, *IFNGR*, have been shown to increase host susceptibility to BCG (66, 67). The loss of the innate immunity provided by 25-HC in the three patients may contribute to the presentation of the BCG abscess. Interestingly, recent studies of humans infected by TB found elevated plasma levels of cholest-4-en-3-one produced by oxidation of cholesterol by the *M. tuberculosis* enzyme 3β-hydroxysteroid dehydrogenase (3β-hsd), an activity the human 3β-HSD enzyme does not possess (68). In the three patients with the homozygous deletion of *CH25H* and *LIPA*, cholest-4-en-3-one was elevated at the time point before and/or following HSCT, falling to parental levels with time (Supplemental Figure S9). An additional factor which may or may not be relevant to the presentation of BCG abscesses in the patients with the *CH25H – LIPA* homozygous deletion is that deletion of *Ch25h* in mouse causes an increase in serum immunoglobin A, ascribed to a removal of the suppressive effect of 25-HC on class switching in B cells (17). Whether a similar mechanism applies to human is yet to be established. Further studies are ongoing into the effects of CH25H and/or LIPA deficiency on mycobacterial susceptibility.

### Autoxidation

What might be the explanation for the elevation of the cholesterol autoxidation products 3β,5α,6β-triol, 7-OC, 7β-HC, their sterol acids, 3β,5α,6β-triHCa, 3β,7β-diHCA, 3βH,7O-CA, and the cholestenoic acids of the acidic pathway, 3β-HCA, 3β,7α-diHCA, 3β,7α,12α-triHCA, at the first time point following HSCT? A point to note, is that the cells transplanted are hematopoietic, i.e., cells responsible for production of blood cells in the bone marrow, and resultant donor-derived macrophages will express not only *CH25H* but also, like recipient macrophages, *CYP27A1* (38, 40). A plausible scenario is that there is an increase in cholesterol autoxidation products in circulating donor-derived white cells expressing LAL as a consequence of elevated substrate availability following LAL-mediated release of non-esterified cholesterol. These cholesterol autoxidation products themselves are substrates for CYP27A1 expressed in macrophages and will lead to the production of the sterol acids 3β,5α,6β-triHCa, 3β,7β-diHCA, 3βH,7O-CA. The enhanced availability of cholesterol in white blood cells can also explain the increased abundance of extrahepatic 3β-HCA and 3β,7α-diHCA. It is noticeable that it is extrahepatic acids that are elevated at the first time point after HSCT which coincide with maximal plasma cholesterol concentrations. The cholesterol metabolites of the hepatic neutral pathway of bile acid biosynthesis, other than 7α-HC that can be formed by autoxidation, do not rise to a maximum in parallel to plasma cholesterol, this suggest that transfer of LAL to hepatic cells is a less efficient process than expression of LAL *per se* in white blood cells.

### Wolman Disease with intact CH25H

We have previously analysed plasma from patients with Wolman disease with functional *CH25H.* Although present at low levels, 25-HC was detected in plasma from these patients, and concentrations of 3β,5α,6β-triol, 7β-HC and 7-OC were found to be elevated as were concentrations of the corresponding C_27_ acids i.e. 3β,5α,6β-triHCA, 3β,7β-diHCA and 3βH,7O-CA (11). In contrast to the patients with the *LIPA* - *CH25H* homozygous deletion, patients with classical Wolman disease did not show elevated levels of 3β-HCA and levels of 3β,7α-diHCA were essentially normal. Unfortunately, we did not perform a time course study on these Wolman patients and so cannot dissect the exact effects of *CH25H* deletion from those of HSCT. However, the *CH25H-LIPA* homozygous deletion does appear to specifically enhance the formation of 3β-HCA. 3β-HCA is formed extrahepatically by macrophages from cholesterol by the enzyme CYP27A1 (38, 40, 69, 70), and elevation in its formation coinciding with *CH25H* deletion would fit the hypothesis that 25-HC, the metabolic product of CH25H, has a function to inhibit the synthesis and uptake of cholesterol by macrophages (25). In the absence of *CH25H* and 25-HC, excess cellular cholesterol becomes converted by macrophages to 3β-HCA and the extrahepatic acidic pathway of bile acid biosynthesis stimulated.

### Cautions and Caveats

There are a number of caveats to the present study. Firstly, only three patients have been analysed and the time course data is opportunistic rather than planned. Secondly, HSCT was performed according to patient need and the availability of donor cells, so the biochemical phenotype differs between patients at the time of transplant. Thirdly the three patients were of different ages at the time of transplant, the degree of chimerism differed as did the dose of enzyme replacement therapy pre and post-HSCT. Despite these imperfections there is good evidence for time-course changes in metabolite levels in response to HSCT allowing the development of a plausible hypothesis to explain these changes. Finally, it should be noted that this is the first report of the plasma metabolite profile of patients with the *CH25H-LIPA* homozygous deletion.

## Conclusions

In summary, in the current study we show that 7α,25-dihydroxysterols but not 25-HC can be formed in the absence of *CH25H*. We also present a plausible explanation for the development of BCG-site abscesses in patients with a homozygous deletion of *CH25H* and *LIPA* and show that HSCT with donor cells without such deletions result in attenuation of cholesterol autoxidation, decreasing the plasma levels of both primary autoxidation products and their enzymatically derived products with time. Finally, we provide a rational for the enhanced formation of sterol acids at the first time point following HSCT based on enhanced extra-hepatic availability of non-esterified cholesterol.

## Data Availability

All data produced in the present study are available upon reasonable request to the authors

## Acknowledgements

Work in Swansea was supported by UKRI (grant numbers BB/I001735/1, BB/N015932/1, BB/S019588/1, MR/X012387/1, MR/Y008057/1 and BB/L001942/1), the European Union through European Structural Funds (ESF), as part of the Welsh Government funded Academic Expertise for Business project (to WJG and YW). Members of the European Network for Oxysterol Research (ENOR, https://www.oxysterols.net/) are thanked for informative discussions. GMB and WGN are supported by the NIHR-MRC (MR/Y008340/1) and the NIHR Manchester Biomedical Research Centre (NIHR 203308).

## Supplemental Table and Figures

**Supplemental Table S1.** Oxysterols identified in plasma of patients with the *CH25H-LIPA* homozygous deletion. Oxysterols are colour coded according to the pathway in which they appear. Patients (with anonymised code) are colour coded with their parents according to pre-and post-HSCT.

**Figure S1.** Schematic illustration of the cellular processes affected by LAL and CH25H enzymes. Blue arrows indicate a process, red arrows a reaction, black arrows transport, purple arrows with a diamond arrowhead activation of a nuclear receptor and green arrows with an oval head enzyme catalysis. A crossed arrow indicates inhibition. Suggested minor reaction is indicated by broken arrows.

**Figure S2.** Derivatisation of oxysterols with GP reagents. (A) Oxysterols with a 3β-hydroxy-5-ene group are converted to their 3-oxo-4-ene analogues by bacterial cholesterol oxidase then reacted with [^2^H_5_]GP. (B) In the absence of cholesterol oxidase oxysterols containing an oxo group will react with [^2^H_0_]GP. In the case of e.g. 7α,25-diHC and 7α,25-diHCO both will be derivatised with [^2^H_5_]GP to give the same product, but only 7α,25-diHCO will be derivatised with [^2^H_0_]GP. 7α,25-diHC can easily be differentiated from 7α,25-diHCO by subtraction of (B) from (A). (C) GP derivatisation gives cis and trans isomers about the CN double bond. Shown in (D) are MRM-like fragmentation pathways highlighting mono-hydroxycholesterols and dihydroxysterols.

**Figure S3.** Oxysterols found in plasma and the pathways of their formation. (A) Metabolites of the neutral and acidic pathways of bile acid biosynthesis are in blue and red, respectively. Metabolites can cross over between pathways. Metabolites in lavender fall mostly into the acidic pathway. Metabolites of the 25-hydroxylase pathway are in teal. Metabolites not analysed are shown on a white background. Enzymes are shown on a green background. Presumed enzyme activity is indicated by an underline of the enzyme abbreviation. Postulated pathway is indicated by broken arrows. (B) An expanded view of the pathway to include 3β,7α,12α-triHCA. (C) Metabolites of the 7β-HC, 7-OC and 3β,5α,6β-triol pathways. (D) Simplified view of cholesterol biosynthesis and start of the 24S-hydroxylase pathway. The pathways are shown with chemical structures in Supplemental Figure S4. For simplicity the enzyme activities of SLC27A2 (VLCS) and SLC27A5 (BACS) which catalyse the removal of the CoA group from cholestenoyl/cholestanoyl and cholenoyl/cholanoyl CoA-thioesters have been omitted.

**Supplemental Figure S4.** Chemical structures showing oxysterols in the (A) acidic pathway branch 1, (B) acidic pathway branch 2 and neutral pathway branch 1, (C) neutral pathway branch 2, and cross over with the acidic pathway. (D) 7β-HC and 7-OC pathways, (E) 24-hydroxylase pathway (CYP46A1), (F) 25-hydroxylase pathway. Metabolites can cross over between pathways. For simplicity the enzymes SLC27A5 (BACS) or SLC27A2 (VLCS) which catalyse the removal of the CoA group from cholestenoyl/cholestanoyl and cholenoyl/cholanoyl CoA-thioesters have been omitted. Although the enzyme HSD3B1 normally converts C_19_ and C_21_ steroids with a 3β-hydroxy-5-ene structure to 3-oxo-4- ene products it also has minor activity towards C_27_ sterols e.g. 26-HC and 3β-HCA (71).

**Supplemental Figure S5.** 25-HC is absent from plasma from patients with the *CH25H – LIPA* homozygous deletion prior to HSCT but is present following transplant. 7α,25-Dihydroxysterols are present in these patients before and after HSCT. Multiple reaction monitoring (MRM) – like chromatograms targeting monohydroxycholesterols (539.4→455.4→437.4) for the three patients, before (top), at the first time point after (centre) and at the last time point (bottom). (A) Patient 08- 008, (B) 08-002 and (C) 08-006. While the peak eluting between the two 24S-HC epimers corresponds to 25-HC in the samples collected after transplant (centre and bottom), the minor peak eluting in samples collected before transplant (top) does not give a MS^3^ spectrum compatible with 25-HC. MRM–like chromatograms targeting dihydroxysterols (555.4→471.4→453.3) for the three patients before (top), at the first time point after (centre) and at the last time point (bottom). (D) Patient 08- 008, (E) 08-002 and (F) 08-006. Structures of MRM ions are shown in Supplemental Figure S2C. Note, the chromatograms shown in (A), (B) and (C) were recorded with a 37 min gradient, those shown in (D), (E) and (F) with a 17 min gradient. There is some variation in retention time, as data was recorded over a number of years. Chromatogram in (A), (B) and (C) are aligned to 24S-HC. Shown in Figure 2 are the corresponding RICs generated from high resolution accurate mass data. MS^3^ spectra of the identified oxysterols are shown in Supplemental Figure S6.

**Supplemental Figure S6**. Typical MS^3^ ([M]^+^→[M-Py]^+^→) spectra of monohydroxycholesterols and dihydroxysterols. Spectra were recorded from patient 08-008 at the first time point following HSCT. Spectra are for the [^2^H_5_]GP-derivatised molecules (fraction A in the sample preparation protocol). (A) 24S-HC, (B) 25-HC, (C) 26-HC, (D) 7β-HC, (E) 7α-HC^‡^ (7α-HC + 7α-HCO), (F) 3β,5α,6β-triol* [M-H_2_O]^+^, (G) 7α,25-diHCO^‡^ (7α,25-diHC + 7α,25-diHCO), (H) 7α,26-diHCO^‡^ (7α,26-diHC + 7α,26-diHCO). 3β,5α,6β-triol* is the dehydration product of 3β,5α,6β-triol. 7α-HC is differentiated from 7α-HCO by also generating appropriate RIC for the [^2^H_0_]GP-derivatised molecules (fraction B in the sample preparation protocol). (I) Schematic depicting fragmentation nomenclature.

**Supplemental Figure S7.** Variation of the concentrations of 24S-HC, and 24S,25-EC before and after HSCT. (A) Patient 08-008, (B) patient 08-002 and (C) patient 08-006. Note that the axis have been split to aid visualisation.

**Supplemental Figure S8**. Typical MS^3^ spectra of sterol acids. Spectra were recorded from patient 08- 008 at the first time point following HSCT. (A) 3β,7β-diHCA, (B) 3β,5α,6β-triHCa*, (C) 3β,7α-diHCA^‡^, (D) 3βH,7O-CA. (E) Spectrum of 3β,25-diHCA recorded from patient 08-002 at the first time point following HSCT. 3β,5α,6β-triHCa* corresponds to the [M-H_2_O]^+^ ion.

**Supplemental Figure S9**. Variation of the concentrations of cholesterol and its precursors desmosterol and 7-DHC (7-DHC isomerises to 8-DHC) and the bacterial product cholestenone (see metabolites in green in Supplemental Figure S3D) before and after HSCT. (A) Patient 08-008, (B) patient 08-002 and (C) patient 08-006. Note that the axis have been split to aid visualisation.

## References

1. Brown, M. S., and J. L. Goldstein. 1979. Receptor-mediated endocytosis: insights from the lipoprotein receptor system. Proc Natl Acad Sci U S A 76: 3330–3337.

2. Infante, R. E., M. L. Wang, A. Radhakrishnan, H. J. Kwon, M. S. Brown, and J. L. Goldstein. 2008. NPC2 facilitates bidirectional transfer of cholesterol between NPC1 and lipid bilayers, a step in cholesterol egress from lysosomes. Proceedings of the National Academy of Sciences 105: 15287–15292.

3. Trinh, M. N., M. S. Brown, J. L. Goldstein, J. Han, G. Vale, J. G. McDonald, J. Seemann, J. T. Mendell, and F. Lu. 2020. Last step in the path of LDL cholesterol from lysosome to plasma membrane to ER is governed by phosphatidylserine. Proc Natl Acad Sci U S A 117: 18521–18529.

4. Goldstein, J. L., R. A. DeBose-Boyd, and M. S. Brown. 2006. Protein sensors for membrane sterols. Cell 124: 35–46.

5. Lund, E. G., T. A. Kerr, J. Sakai, W. P. Li, and D. W. Russell. 1998. cDNA cloning of mouse and human cholesterol 25-hydroxylases, polytopic membrane proteins that synthesize a potent oxysterol regulator of lipid metabolism. J Biol Chem 273: 34316–34327.

6. Radhakrishnan, A., Y. Ikeda, H. J. Kwon, M. S. Brown, and J. L. Goldstein. 2007. Sterol-regulated transport of SREBPs from endoplasmic reticulum to Golgi: oxysterols block transport by binding to Insig. Proc Natl Acad Sci U S A 104: 6511–6518.

7. Zhang, H. 2018. Lysosomal acid lipase and lipid metabolism: new mechanisms, new questions, and new therapies. Current opinion in lipidology 29: 218–223.

8. Potter, J. E., G. Petts, A. Ghosh, F. J. White, J. L. Kinsella, S. Hughes, J. Roberts, A. Hodgkinson, K. Brammeier, H. Church, C. Merrigan, J. Hughes, P. Evans, H. Campbell, D. Bonney, W. G. Newman, B. W. Bigger, A. Broomfield, S. A. Jones, and R. F. Wynn. 2021. Enzyme replacement therapy and hematopoietic stem cell transplant: a new paradigm of treatment in Wolman disease. Orphanet Journal of Rare Diseases 16: 235.

9. Hoffman, E. P., M. L. Barr, M. A. Giovanni, and M. F. Murray. 2015. Lysosomal Acid Lipase Deficiency. *In* GeneReviews. M. P. Adam, H. H. Ardinger, and R. A. Pagon, editors. University of Washington, Seattle;.

10. Boenzi, S., F. Deodato, R. Taurisano, B. M. Goffredo, C. Rizzo, and C. Dionisi-Vici. 2016. Evaluation of plasma cholestane-3beta,5alpha,6beta-triol and 7-ketocholesterol in inherited disorders related to cholesterol metabolism. J Lipid Res 57: 361–367.

11. Griffiths, W. J., E. Yutuc, J. Abdel-Khalik, P. J. Crick, T. Hearn, A. Dickson, B. W. Bigger, T. Hoi-Yee Wu, A. Goenka, A. Ghosh, S. A. Jones, D. F. Covey, D. S. Ory, and Y. Wang. 2019. Metabolism of Non-Enzymatically Derived Oxysterols: Clues from sterol metabolic disorders. Free Radic Biol Med 144: 124–133.

12. Cooper, J. A., H. J. Church, and H. Y. Wu. 2020. Cholestane-3β, 5α, 6β-triol: Further insights into the performance of this oxysterol in diagnosis of Niemann-Pick disease type C. Molecular Genetics and Metabolism 130: 77–86.

13. de Castro Lopez, M. J., F. J. White, V. Holmes, J. Roberts, T. H. Y. Wu, J. A. Cooper, H. J. Church, G. Petts, R. F. Wynn, S. A. Jones, and A. Ghosh. 2025. Does Early Diagnosis and Treatment Alter the Clinical Course of Wolman Disease? Divergent Trajectories in Two Siblings and a Consideration for Newborn Screening. Int J Neonatal Screen 11.

14. Jiang, X., R. Sidhu, F. D. Porter, N. M. Yanjanin, A. O. Speak, D. T. te Vruchte, F. M. Platt, H. Fujiwara, D. E. Scherrer, J. Zhang, D. J. Dietzen, J. E. Schaffer, and D. S. Ory. 2011. A sensitive and specific LC-MS/MS method for rapid diagnosis of Niemann-Pick C1 disease from human plasma. J Lipid Res 52: 1435–1445.

15. Patrick, A. D., and B. D. Lake. 1969. Deficiency of an acid lipase in Wolman’s disease. Nature 222: 1067–1068.

16. Griffiths, W. J., I. Gilmore, E. Yutuc, J. Abdel-Khalik, P. J. Crick, T. Hearn, A. Dickson, B. W. Bigger, T. H. Wu, A. Goenka, A. Ghosh, S. A. Jones, and Y. Wang. 2018. Identification of unusual oxysterols and bile acids with 7-oxo or 3beta,5alpha,6beta-trihydroxy functions in human plasma by charge-tagging mass spectrometry with multistage fragmentation. J Lipid Res 59: 1058–1070.

17. Bauman, D. R., A. D. Bitmansour, J. G. McDonald, B. M. Thompson, G. Liang, and D. W. Russell. 2009. 25-Hydroxycholesterol secreted by macrophages in response to Toll-like receptor activation suppresses immunoglobulin A production. Proc Natl Acad Sci U S A 106: 16764–16769.

18. Diczfalusy, U., K. E. Olofsson, A. M. Carlsson, M. Gong, D. T. Golenbock, O. Rooyackers, U. Flaring, and H. Bjorkbacka. 2009. Marked upregulation of cholesterol 25-hydroxylase expression by lipopolysaccharide. J Lipid Res 50: 2258–2264.

19. Park, K., and A. L. Scott. 2010. Cholesterol 25-hydroxylase production by dendritic cells and macrophages is regulated by type I interferons. J Leukoc Biol 88: 1081–1087.

20. Blanc, M., W. Y. Hsieh, K. A. Robertson, K. A. Kropp, T. Forster, G. Shui, P. Lacaze, S. Watterson, S. J. Griffiths, N. J. Spann, A. Meljon, S. Talbot, K. Krishnan, D. F. Covey, M. R. Wenk, M. Craigon, Z. Ruzsics, J. Haas, A. Angulo, W. J. Griffiths, C. K. Glass, Y. Wang, and P. Ghazal. 2013. The transcription factor STAT-1 couples macrophage synthesis of 25-hydroxycholesterol to the interferon antiviral response. Immunity 38: 106–118.

21. Liu, S. Y., R. Aliyari, K. Chikere, G. Li, M. D. Marsden, J. K. Smith, O. Pernet, H. Guo, R. Nusbaum, J. A. Zack, A. N. Freiberg, L. Su, B. Lee, and G. Cheng. 2013. Interferon-inducible cholesterol-25- hydroxylase broadly inhibits viral entry by production of 25-hydroxycholesterol. Immunity 38: 92–105.

22. Zang, R., J. B. Case, E. Yutuc, X. Ma, S. Shen, M. F. Gomez Castro, Z. Liu, Q. Zeng, H. Zhao, J. Son, P. W. Rothlauf, A. J. B. Kreutzberger, G. Hou, H. Zhang, S. Bose, X. Wang, M. D. Vahey, K. Mani, W. J. Griffiths, T. Kirchhausen, D. H. Fremont, H. Guo, A. Diwan, Y. Wang, M. S. Diamond, S. P. J. Whelan, and S. Ding. 2020. Cholesterol 25-hydroxylase suppresses SARS-CoV-2 replication by blocking membrane fusion. Proc Natl Acad Sci U S A 117: 32105–32113.

23. Reboldi, A., E. V. Dang, J. G. McDonald, G. Liang, D. W. Russell, and J. G. Cyster. 2014. Inflammation. 25-Hydroxycholesterol suppresses interleukin-1-driven inflammation downstream of type I interferon. Science 345: 679–684.

24. Cyster, J. G., E. V. Dang, A. Reboldi, and T. Yi. 2014. 25-Hydroxycholesterols in innate and adaptive immunity. Nat Rev Immunol 14: 731–743.

25. Dang, E. V., J. G. McDonald, D. W. Russell, and J. G. Cyster. 2017. Oxysterol Restraint of Cholesterol Synthesis Prevents AIM2 Inflammasome Activation. Cell 171: 1057–1071 e1011.

26. Jang, J., S. Park, H. Jin Hur, H. J. Cho, I. Hwang, Y. Pyo Kang, I. Im, H. Lee, E. Lee, W. Yang, H. C. Kang, S. Won Kwon, J. W. Yu, and D. W. Kim. 2016. 25-hydroxycholesterol contributes to cerebral inflammation of X-linked adrenoleukodystrophy through activation of the NLRP3 inflammasome. Nat Commun 7: 13129.

27. Janowski, B. A., M. J. Grogan, S. A. Jones, G. B. Wisely, S. A. Kliewer, E. J. Corey, and D. J. Mangelsdorf. 1999. Structural requirements of ligands for the oxysterol liver X receptors LXRalpha and LXRbeta. Proc Natl Acad Sci U S A 96: 266–271.

28. Lehmann, J. M., S. A. Kliewer, L. B. Moore, T. A. Smith-Oliver, B. B. Oliver, J. L. Su, S. S. Sundseth, D. A. Winegar, D. E. Blanchard, T. A. Spencer, and T. M. Willson. 1997. Activation of the nuclear receptor LXR by oxysterols defines a new hormone response pathway. J Biol Chem 272: 3137–3140.

29. Wang, B., and P. Tontonoz. 2018. Liver X receptors in lipid signalling and membrane homeostasis. Nat Rev Endocrinol 14: 452–463.

30. Cheng, D., C. C. Y. Chang, X.-m. Qu, and T.-Y. Chang. 1995. Activation of Acyl-Coenzyme A:Cholesterol Acyltransferase by Cholesterol or by Oxysterol in a Cell-free System (&#x2217;). Journal of Biological Chemistry 270: 685–695.

31. Honda, A., T. Miyazaki, T. Ikegami, J. Iwamoto, T. Maeda, T. Hirayama, Y. Saito, T. Teramoto, and Y. Matsuzaki. 2011. Cholesterol 25-hydroxylation activity of CYP3A. J Lipid Res 52: 1509–1516.

32. Griffiths, W. J., P. J. Crick, A. Meljon, S. Theofilopoulos, J. Abdel-Khalik, E. Yutuc, J. E. Parker, D. E. Kelly, S. L. Kelly, E. Arenas, and Y. Wang. 2019. Additional pathways of sterol metabolism: Evidence from analysis of Cyp27a1-/- mouse brain and plasma. Biochim Biophys Acta Mol Cell Biol Lipids 1864: 191–211.

33. Hannedouche, S., J. Zhang, T. Yi, W. Shen, D. Nguyen, J. P. Pereira, D. Guerini, B. U. Baumgarten, S. Roggo, B. Wen, R. Knochenmuss, S. Noel, F. Gessier, L. M. Kelly, M. Vanek, S. Laurent, I. Preuss, C. Miault, I. Christen, R. Karuna, W. Li, D. I. Koo, T. Suply, C. Schmedt, E. C. Peters, R. Falchetto, A. Katopodis, C. Spanka, M. O. Roy, M. Detheux, Y. A. Chen, P. G. Schultz, C. Y. Cho, K. Seuwen, J. G. Cyster, and A. W. Sailer. 2011. Oxysterols direct immune cell migration via EBI2. Nature 475: 524–527.

34. Liu, C., X. V. Yang, J. Wu, C. Kuei, N. S. Mani, L. Zhang, J. Yu, S. W. Sutton, N. Qin, H. Banie, L. Karlsson, S. Sun, and T. W. Lovenberg. 2011. Oxysterols direct B-cell migration through EBI2. Nature 475: 519–523.

35. Wanke, F., S. Moos, A. L. Croxford, A. P. Heinen, S. Graf, B. Kalt, D. Tischner, J. Zhang, I. Christen, J. Bruttger, N. Yogev, Y. Tang, M. Zayoud, N. Israel, K. Karram, S. Reissig, S. M. Lacher, C. Reichhold, I. A. Mufazalov, A. Ben-Nun, T. Kuhlmann, N. Wettschureck, A. W. Sailer, K. Rajewsky, S. Casola, A. Waisman, and F. C. Kurschus. 2017. EBI2 Is Highly Expressed in Multiple Sclerosis Lesions and Promotes Early CNS Migration of Encephalitogenic CD4 T Cells. Cell Rep 18: 1270–1284.

36. Carr, I. M., K. J. Flintoff, G. R. Taylor, A. F. Markham, and D. T. Bonthron. 2006. Interactive visual analysis of SNP data for rapid autozygosity mapping in consanguineous families. Hum Mutat 27: 1041–1046.

37. Yutuc, E., A. L. Dickson, M. Pacciarini, L. Griffiths, P. R. S. Baker, L. Connell, A. Öhman, L. Forsgren, M. Trupp, S. Vilarinho, Y. Khalil, P. T. Clayton, S. Sari, B. Dalgic, P. Höflinger, L. Schöls, W. J. Griffiths, and Y. Wang. 2021. Deep mining of oxysterols and cholestenoic acids in human plasma and cerebrospinal fluid: Quantification using isotope dilution mass spectrometry. Anal Chim Acta 1154: 338259.

38. Björkhem, I., O. Andersson, U. Diczfalusy, B. Sevastik, R. J. Xiu, C. Duan, and E. Lund. 1994. Atherosclerosis and sterol 27-hydroxylase: evidence for a role of this enzyme in elimination of cholesterol from human macrophages. Proc Natl Acad Sci U S A 91: 8592–8596.

39. Lutjohann, D., O. Breuer, G. Ahlborg, I. Nennesmo, A. Siden, U. Diczfalusy, and I. Bjorkhem. 1996. Cholesterol homeostasis in human brain: evidence for an age-dependent flux of 24S- hydroxycholesterol from the brain into the circulation. Proc Natl Acad Sci U S A 93: 9799–9804.

40. Babiker, A., O. Andersson, D. Lindblom, J. van der Linden, B. Wiklund, D. Lutjohann, U. Diczfalusy, and I. Bjorkhem. 1999. Elimination of cholesterol as cholestenoic acid in human lung by sterol 27-hydroxylase: evidence that most of this steroid in the circulation is of pulmonary origin. J Lipid Res 40: 1417–1425.

41. Bjorkhem, I., U. Andersson, E. Ellis, G. Alvelius, L. Ellegard, U. Diczfalusy, J. Sjovall, and C. Einarsson. 2001. From brain to bile. Evidence that conjugation and omega-hydroxylation are important for elimination of 24S-hydroxycholesterol (cerebrosterol) in humans. J Biol Chem 276: 37004–37010.

42. Meaney, S., M. Hassan, A. Sakinis, D. Lutjohann, K. von Bergmann, A. Wennmalm, U. Diczfalusy, and I. Bjorkhem. 2001. Evidence that the major oxysterols in human circulation originate from distinct pools of cholesterol: a stable isotope study. J Lipid Res 42: 70–78.

43. Meaney, S., A. Babiker, D. Lutjohann, U. Diczfalusy, M. Axelson, and I. Bjorkhem. 2003. On the origin of the cholestenoic acids in human circulation. Steroids 68: 595–601.

44. Axelson, M., and J. Sjovall. 1990. Potential bile acid precursors in plasma--possible indicators of biosynthetic pathways to cholic and chenodeoxycholic acids in man. J Steroid Biochem 36: 631–640.

45. Russell, D. W. 2003. The enzymes, regulation, and genetics of bile acid synthesis. Annu Rev Biochem 72: 137–174.

46. Abdel-Khalik, J., P. J. Crick, E. Yutuc, A. E. DeBarber, P. B. Duell, R. D. Steiner, I. Laina, Y. Wang, and W. J. Griffiths. 2018. Identification of 7alpha,24-dihydroxy-3-oxocholest-4-en-26-oic and 7alpha,25-dihydroxy-3-oxocholest-4-en-26-oic acids in human cerebrospinal fluid and plasma. Biochimie 153: 86–98.

47. Wang, Y., E. Yutuc, and W. J. Griffiths. 2021. Cholesterol metabolism pathways - are the intermediates more important than the products? FEBS J.

48. Abdel-Khalik, J., T. Hearn, A. L. Dickson, P. J. Crick, E. Yutuc, K. Austin-Muttitt, B. W. Bigger, A. A. Morris, C. H. Shackleton, P. T. Clayton, T. Iida, R. Sircar, R. Rohatgi, H. U. Marschall, J. Sjovall, I. Bjorkhem, J. G. L. Mullins, W. J. Griffiths, and Y. Wang. 2020. Bile acid biosynthesis in Smith-Lemli-Opitz syndrome bypassing cholesterol: Potential importance of pathway intermediates. J Steroid Biochem Mol Biol 206: 105794.

49. Mazzacuva, F., P. Mills, K. Mills, S. Camuzeaux, P. Gissen, E. R. Nicoli, C. Wassif, D. Te Vruchte, F. D. Porter, M. Maekawa, N. Mano, T. Iida, F. Platt, and P. T. Clayton. 2016. Identification of novel bile acids as biomarkers for the early diagnosis of Niemann-Pick C disease. FEBS Lett 590: 1651–1662.

50. Jiang, X., R. Sidhu, L. Mydock-McGrane, F. F. Hsu, D. F. Covey, D. E. Scherrer, B. Earley, S. E. Gale, N. Y. Farhat, F. D. Porter, D. J. Dietzen, J. J. Orsini, E. Berry-Kravis, X. Zhang, J. Reunert, T. Marquardt, H. Runz, R. Giugliani, J. E. Schaffer, and D. S. Ory. 2016. Development of a bile acid-based newborn screen for Niemann-Pick disease type C. Sci Transl Med 8: 337ra363.

51. Dzeletovic, S., O. Breuer, E. Lund, and U. Diczfalusy. 1995. Determination of cholesterol oxidation products in human plasma by isotope dilution-mass spectrometry. Anal Biochem 225: 73–80.

52. Zu, S., Y. Q. Deng, C. Zhou, J. Li, L. Li, Q. Chen, X. F. Li, H. Zhao, S. Gold, J. He, X. Li, C. Zhang, H. Yang, G. Cheng, and C. F. Qin. 2020. 25-Hydroxycholesterol is a potent SARS-CoV-2 inhibitor. Cell Res 30: 1043–1045.

53. Viaud, M., S. Ivanov, N. Vujic, M. Duta-Mare, L. E. Aira, T. Barouillet, E. Garcia, F. Orange, I. Dugail, I. Hainault, C. Stehlik, S. Marchetti, L. Boyer, R. Guinamard, F. Foufelle, A. Bochem, K. G. Hovingh, E. B. Thorp, E. L. Gautier, D. Kratky, P. Dasilva-Jardine, and L. Yvan-Charvet. 2018. Lysosomal Cholesterol Hydrolysis Couples Efferocytosis to Anti-Inflammatory Oxysterol Production. Circ Res 122: 1369–1384.

54. Schule, R., T. Siddique, H. X. Deng, Y. Yang, S. Donkervoort, M. Hansson, R. E. Madrid, N. Siddique, L. Schols, and I. Bjorkhem. 2010. Marked accumulation of 27-hydroxycholesterol in SPG5 patients with hereditary spastic paresis. J Lipid Res 51: 819–823.

55. Theofilopoulos, S., W. J. Griffiths, P. J. Crick, S. Yang, A. Meljon, M. Ogundare, S. S. Kitambi, A. Lockhart, K. Tuschl, P. T. Clayton, A. A. Morris, A. Martinez, M. A. Reddy, A. Martinuzzi, M. T. Bassi, A. Honda, T. Mizuochi, A. Kimura, H. Nittono, G. De Michele, R. Carbone, C. Criscuolo, J. L. Yau, J. R. Seckl, R. Schule, L. Schols, A. W. Sailer, J. Kuhle, M. J. Fraidakis, J. A. Gustafsson, K. R. Steffensen, I. Bjorkhem, P. Ernfors, J. Sjovall, E. Arenas, and Y. Wang. 2014. Cholestenoic acids regulate motor neuron survival via liver X receptors. J Clin Invest 124: 4829–4842.

56. Meljon, A., P. J. Crick, E. Yutuc, J. L. Yau, J. R. Seckl, S. Theofilopoulos, E. Arenas, Y. Wang, and W. J. Griffiths. 2019. Mining for Oxysterols in Cyp7b1(-/-) Mouse Brain and Plasma: Relevance to Spastic Paraplegia Type 5. Biomolecules 9.

57. Lund, E. G., J. M. Guileyardo, and D. W. Russell. 1999. cDNA cloning of cholesterol 24- hydroxylase, a mediator of cholesterol homeostasis in the brain. Proc Natl Acad Sci U S A 96: 7238–7243.

58. Russell, D. W., R. W. Halford, D. M. Ramirez, R. Shah, and T. Kotti. 2009. Cholesterol 24- hydroxylase: an enzyme of cholesterol turnover in the brain. Annu Rev Biochem 78: 1017–1040.

59. Goyal, S., Y. Xiao, N. A. Porter, L. Xu, and F. P. Guengerich. 2014. Oxidation of 7- dehydrocholesterol and desmosterol by human cytochrome P450 46A1. J Lipid Res 55: 1933–1943.

60. Dietschy, J. M., and S. D. Turley. 2004. Thematic review series: brain Lipids. Cholesterol metabolism in the central nervous system during early development and in the mature animal. J Lipid Res 45: 1375–1397.

61. Fakheri, R. J., and N. B. Javitt. 2012. 27-Hydroxycholesterol, does it exist? On the nomenclature and stereochemistry of 26-hydroxylated sterols. Steroids 77: 575–577.

62. Goenka, A., A. Ghosh, S. Dixon, W. J. Griffiths, S. M. Hughes, W. G. Newman, J. Urquhart, Y. Wang, R. F. Wynn, T. Hussell, S. A. Jones, and P. D. Arkwright. 2017. Susceptibility to BCG abscess associated with deletion of two cholesterol metabolism genes: lysosomal acid lipase and cholesterol 25-hydroxylase. *In* UKPIN Conference, Brighton, UK.

63. Ormsby, T. J. R., S. E. Owens, A. D. Horlock, D. Davies, W. J. Griffiths, Y. Wang, J. G. Cronin, J. J. Bromfield, and I. M. Sheldon. 2021. Oxysterols protect bovine endometrial cells against pore-forming toxins from pathogenic bacteria. Faseb j 35: e21889.

64. Zhou, Q. D., X. Chi, M. S. Lee, W. Y. Hsieh, J. J. Mkrtchyan, A. C. Feng, C. He, A. G. York, V. L. Bui, E. B. Kronenberger, A. Ferrari, X. Xiao, A. E. Daly, E. J. Tarling, R. Damoiseaux, P. O. Scumpia, S. T. Smale, K. J. Williams, P. Tontonoz, and S. J. Bensinger. 2020. Interferon-mediated reprogramming of membrane cholesterol to evade bacterial toxins. Nat Immunol 21: 746–755.

65. Abrams, M. E., K. A. Johnson, S. S. Perelman, L. S. Zhang, S. Endapally, K. B. Mar, B. M. Thompson, J. G. McDonald, J. W. Schoggins, A. Radhakrishnan, and N. M. Alto. 2020. Oxysterols provide innate immunity to bacterial infection by mobilizing cell surface accessible cholesterol. Nat Microbiol 5: 929–942.

66. Schroder, K., P. J. Hertzog, T. Ravasi, and D. A. Hume. 2004. Interferon-gamma: an overview of signals, mechanisms and functions. J Leukoc Biol 75: 163–189.

67. Jouanguy, E., S. Lamhamedi-Cherradi, D. Lammas, S. E. Dorman, M. C. Fondanèche, S. Dupuis, R. Döffinger, F. Altare, J. Girdlestone, J. F. Emile, H. Ducoulombier, D. Edgar, J. Clarke, V. A. Oxelius, M. Brai, V. Novelli, K. Heyne, A. Fischer, S. M. Holland, D. S. Kumararatne, R. D. Schreiber, and J. L. Casanova. 1999. A human IFNGR1 small deletion hotspot associated with dominant susceptibility to mycobacterial infection. Nat Genet 21: 370–378.

68. Chandra, P., H. Coullon, M. Agarwal, C. W. Goss, and J. A. Philips. 2022. Macrophage global metabolomics identifies cholestenone as host/pathogen cometabolite present in human Mycobacterium tuberculosis infection. J Clin Invest 132.

69. Cali, J. J., and D. W. Russell. 1991. Characterization of human sterol 27-hydroxylase. A mitochondrial cytochrome P-450 that catalyzes multiple oxidation reaction in bile acid biosynthesis. J Biol Chem 266: 7774–7778.

70. Quinn, C. M., W. Jessup, J. Wong, L. Kritharides, and A. J. Brown. 2005. Expression and regulation of sterol 27-hydroxylase (CYP27A1) in human macrophages: a role for RXR and PPARgamma ligands. Biochem J 385: 823–830.

71. Dickson, A., E. Yutuc, C. A. Thornton, J. E. Dunford, U. Oppermann, Y. Wang, and W. J. Griffiths. 2022. HSD3B1 is an Oxysterol 3β-Hydroxysteroid Dehydrogenase in Human Placenta. bioRxiv: 2022.2004.2001.486576.

