## Supplementary figures and images for "Deletion of *CH25H* and *LIPA* Genes in Human Abolishes Biosynthesis of 25-Hydroxycholesterol but not of 7α,25-Dihydroxysterols and Enhances Non-enzymatic Cholesterol Oxidation: Metabolic Changes are Partially Reversed by Hematopoietic Stem Cell Transplant^‡^"

### Figure S1

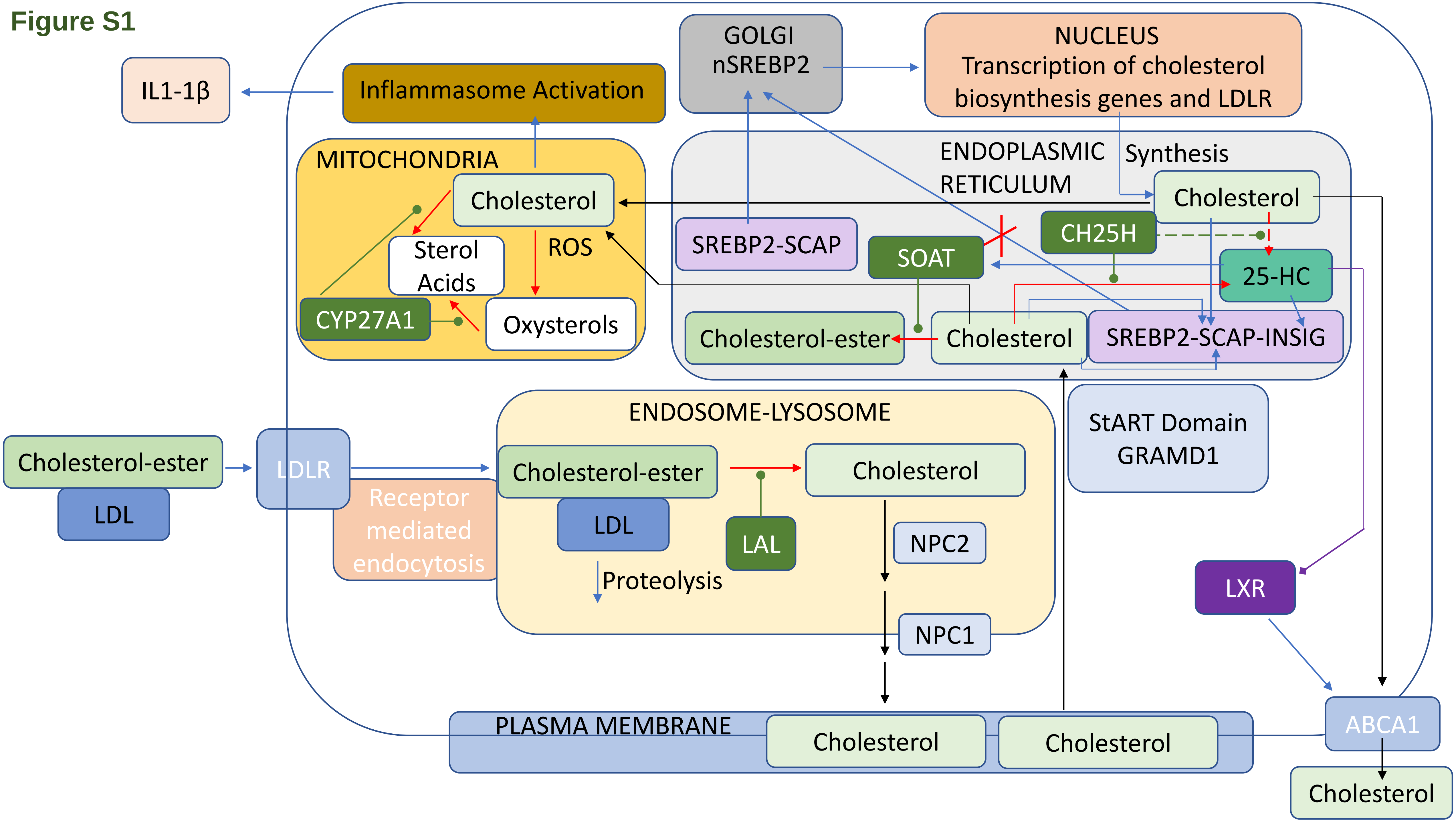

### Figure S3

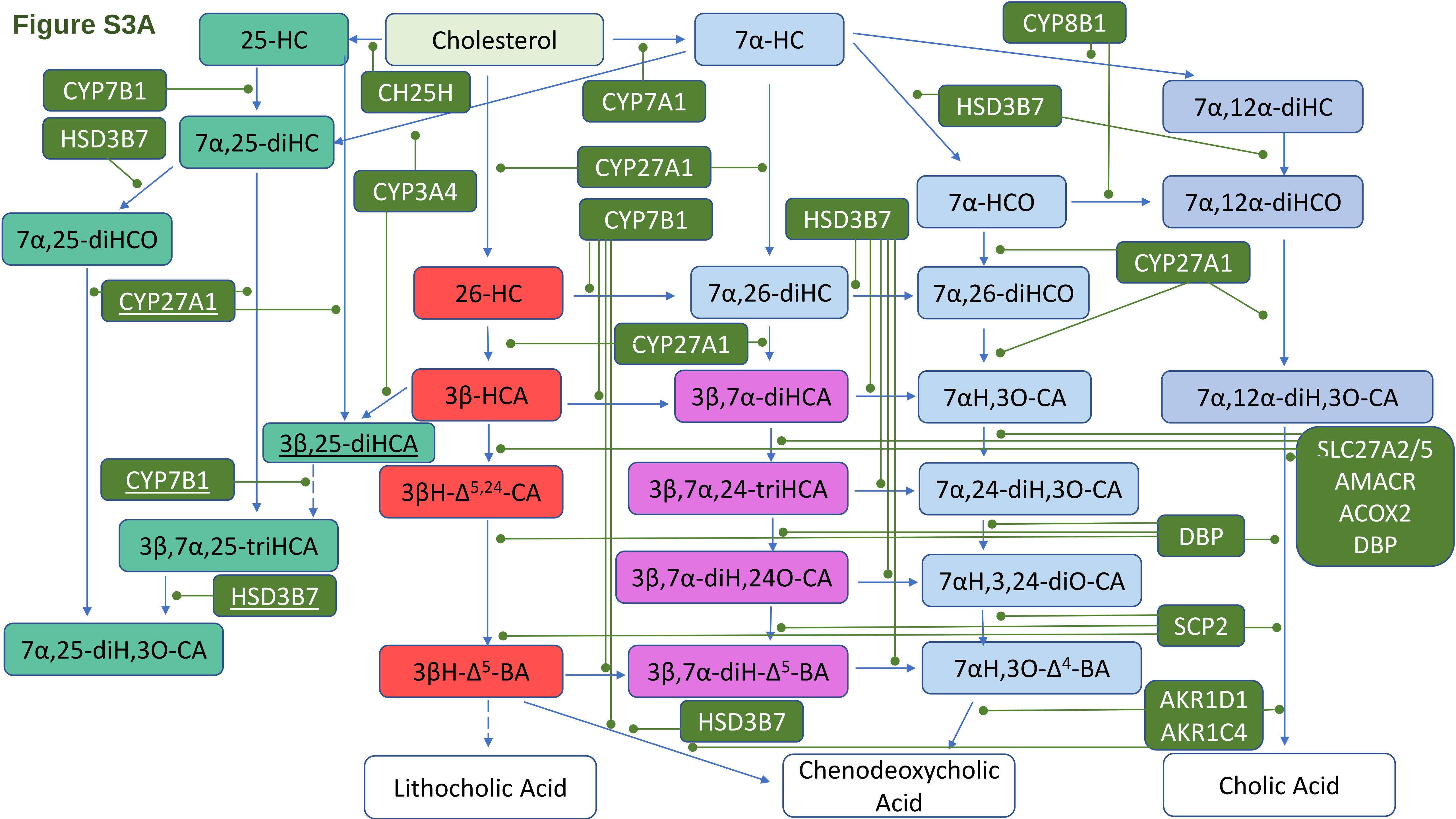

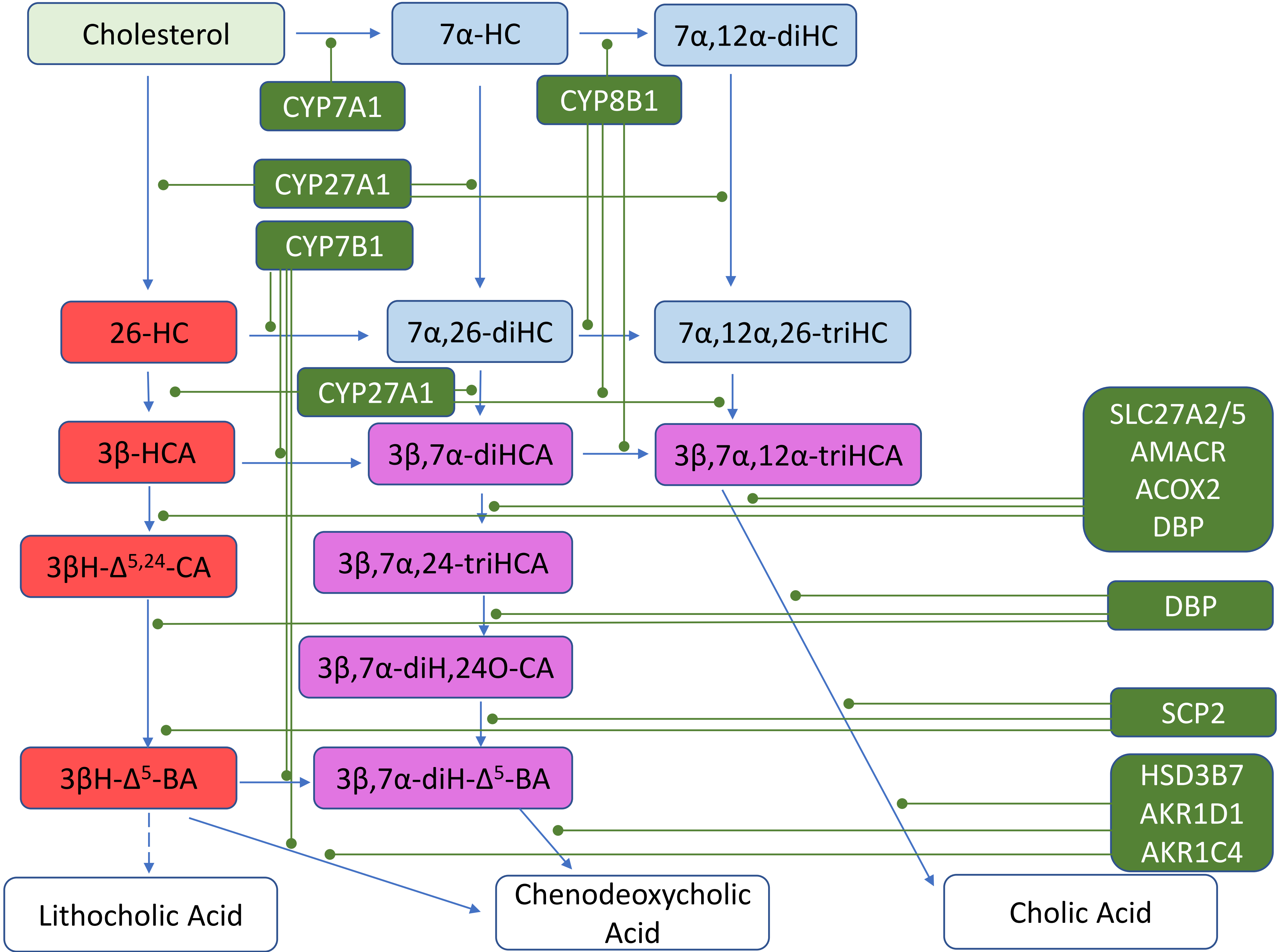

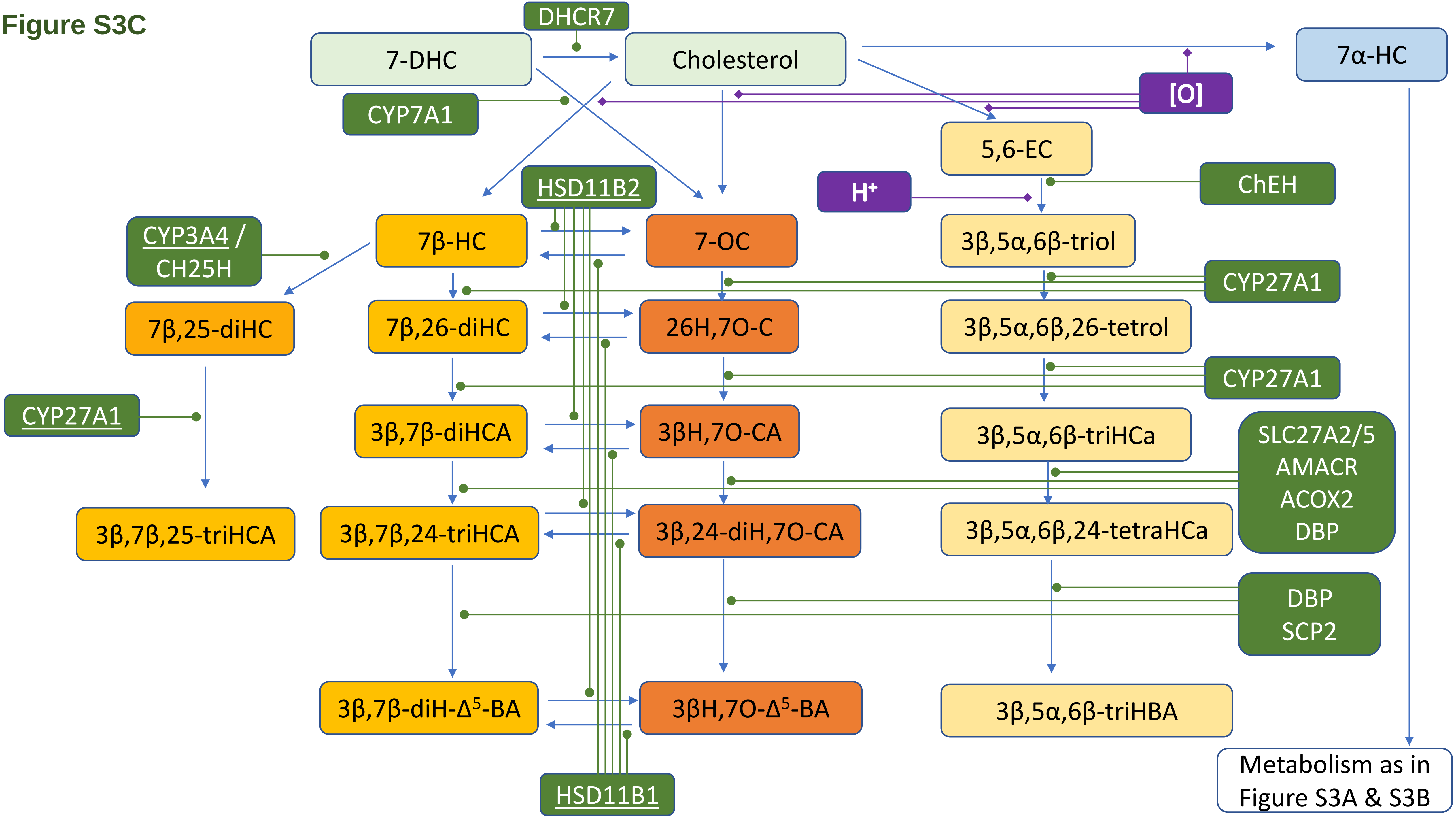

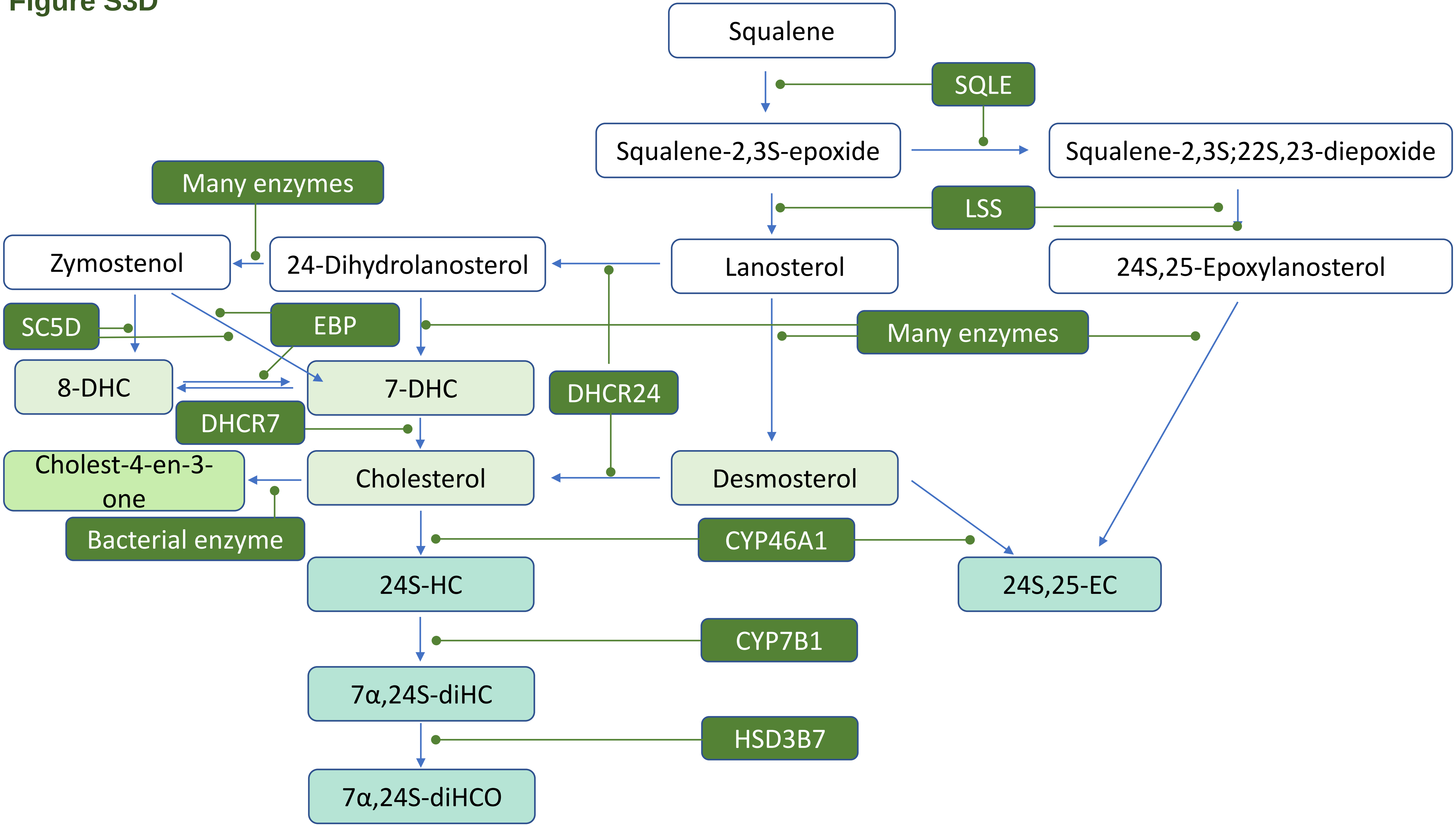

### Figure S4

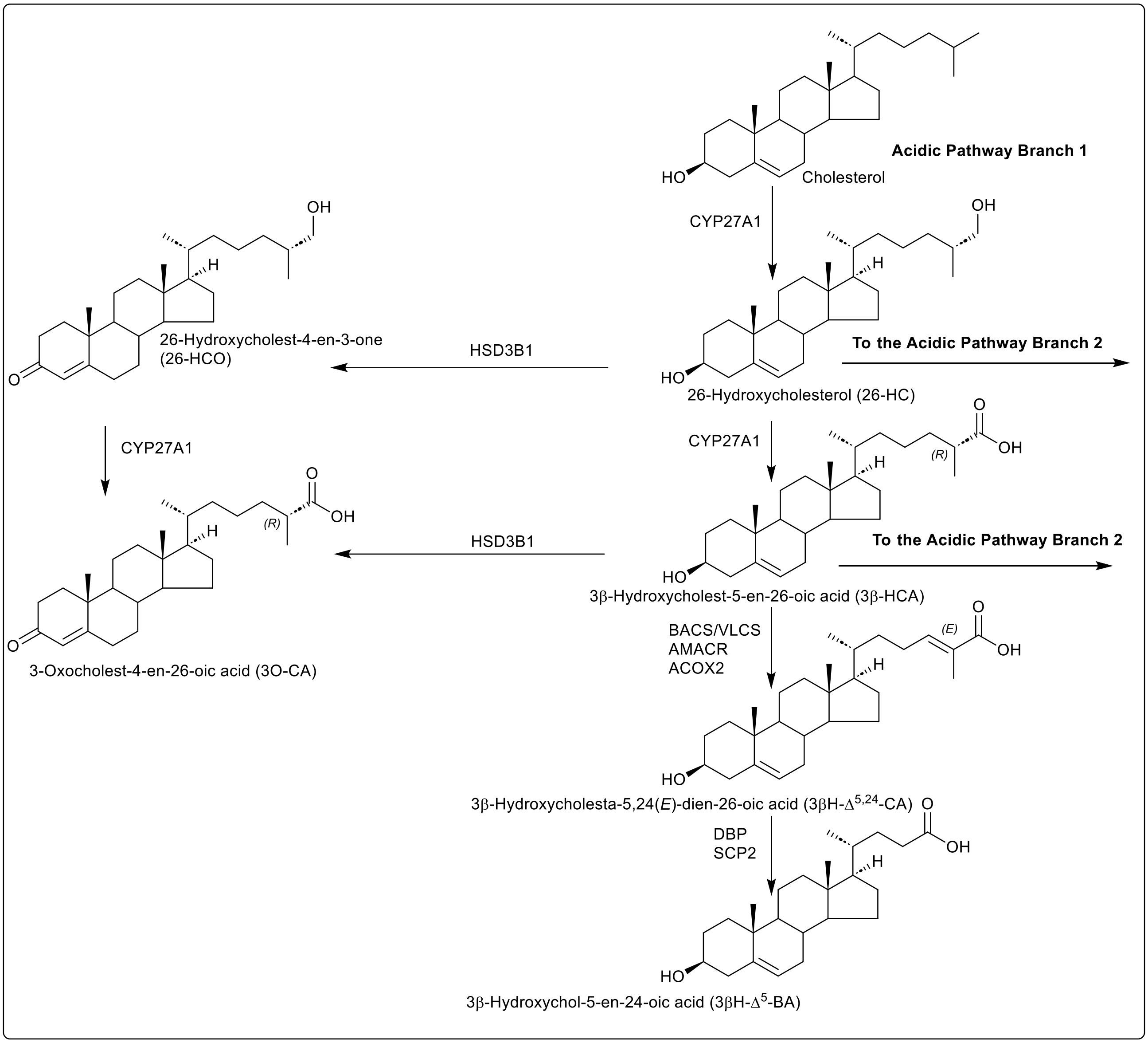

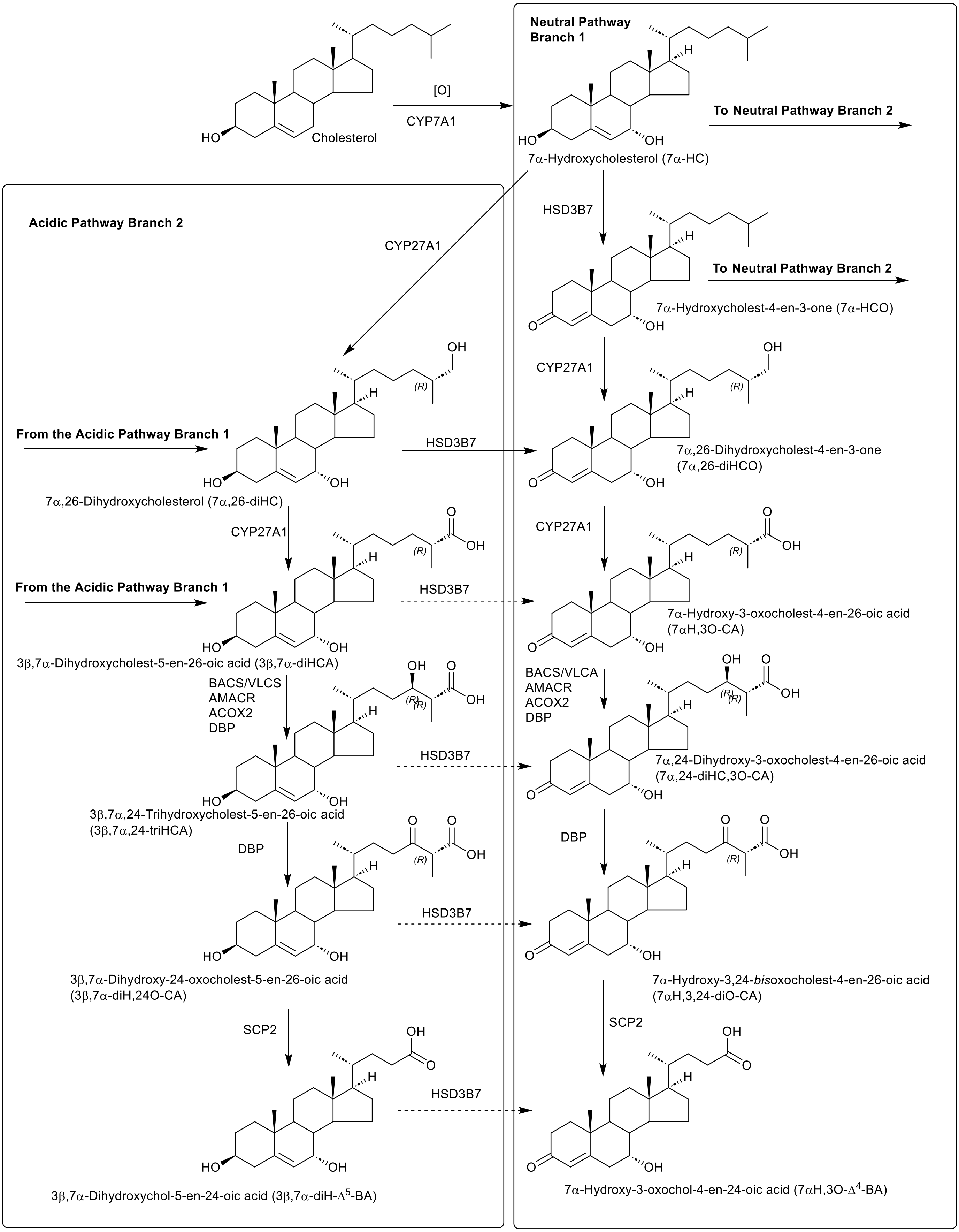

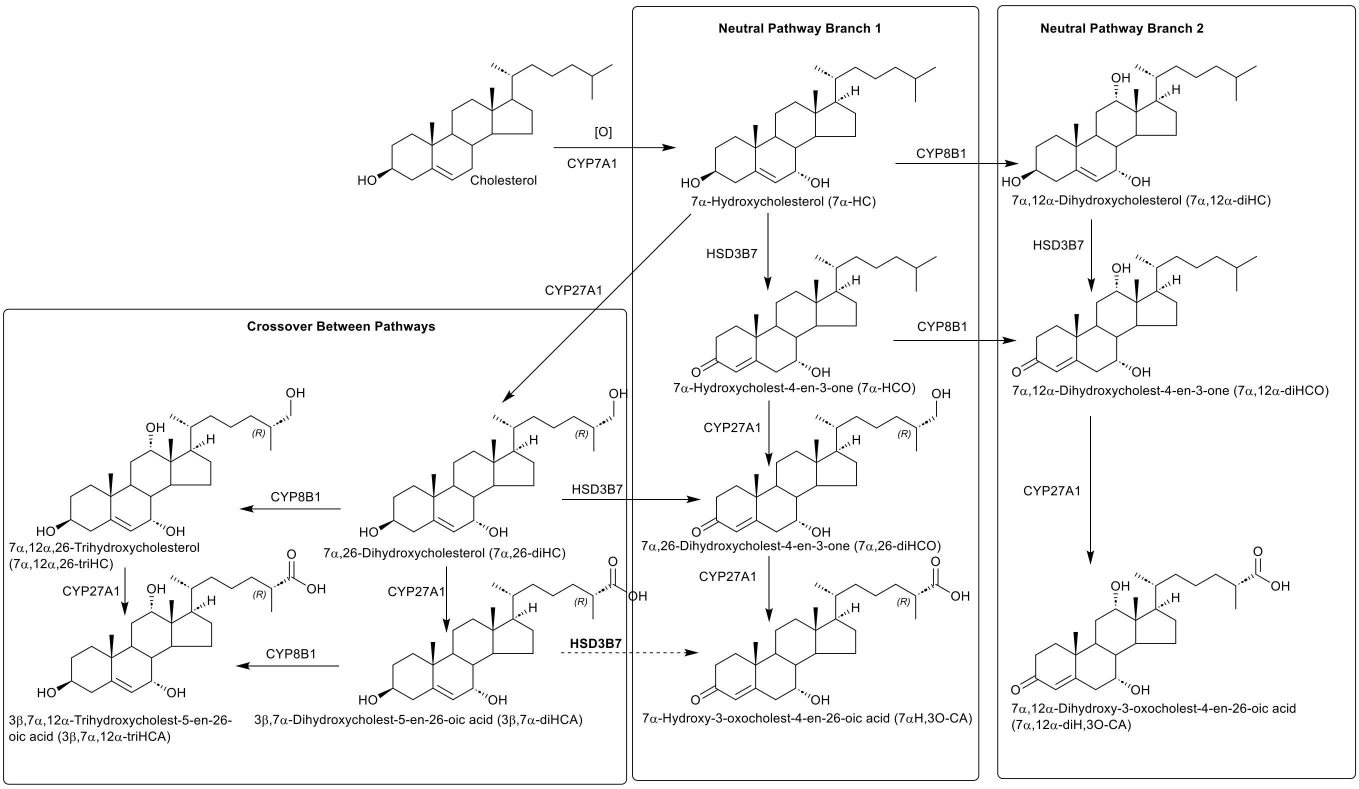

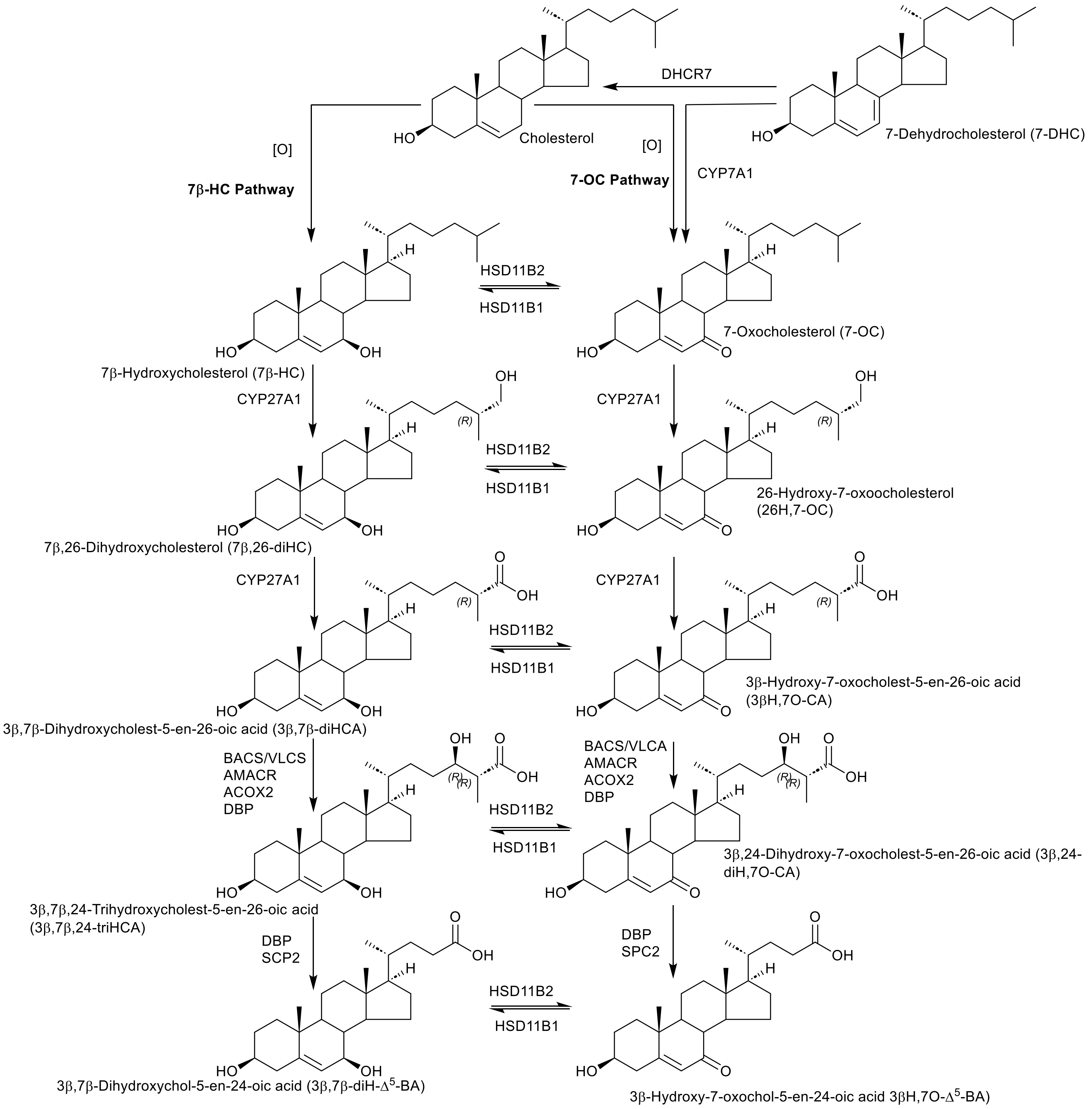

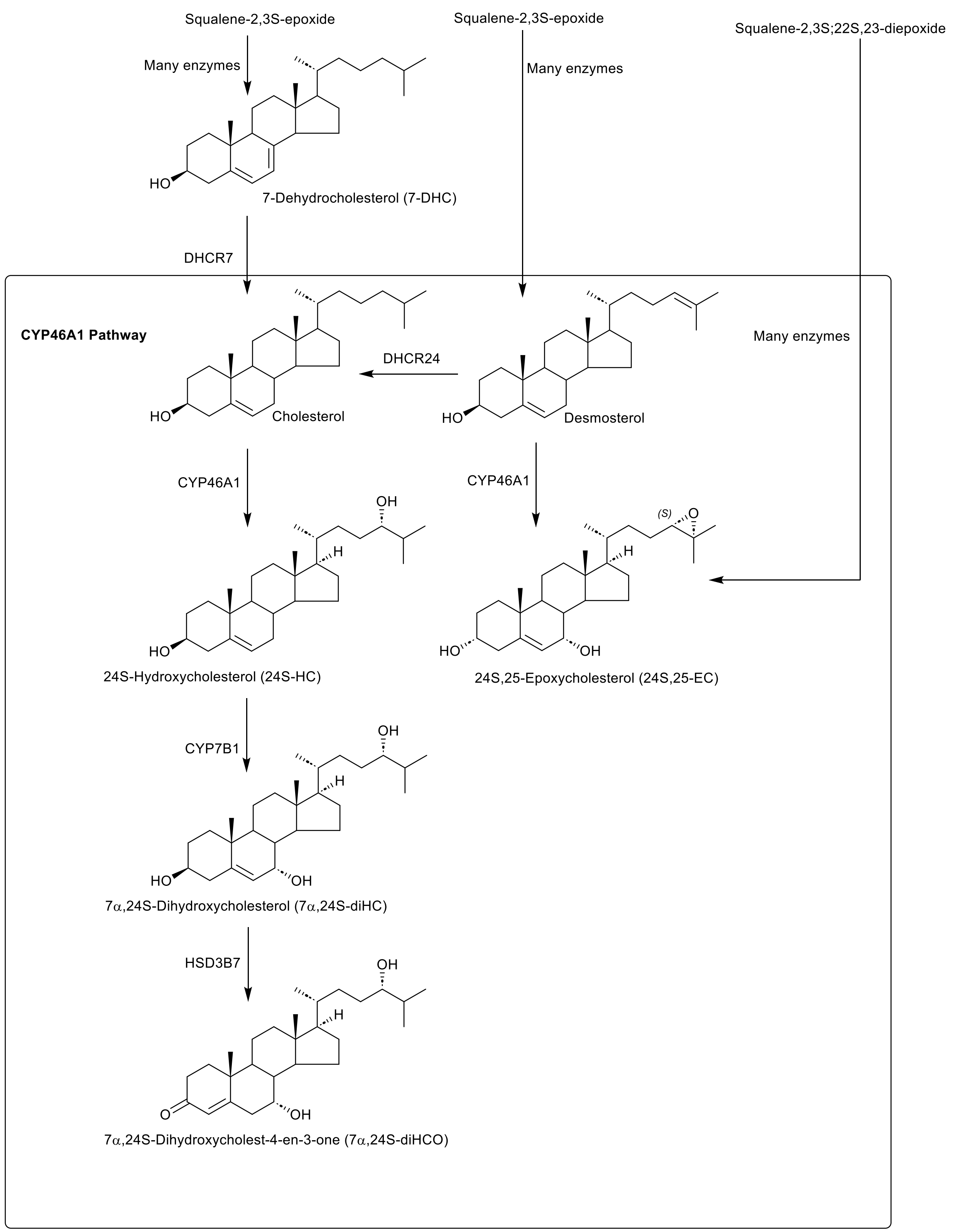

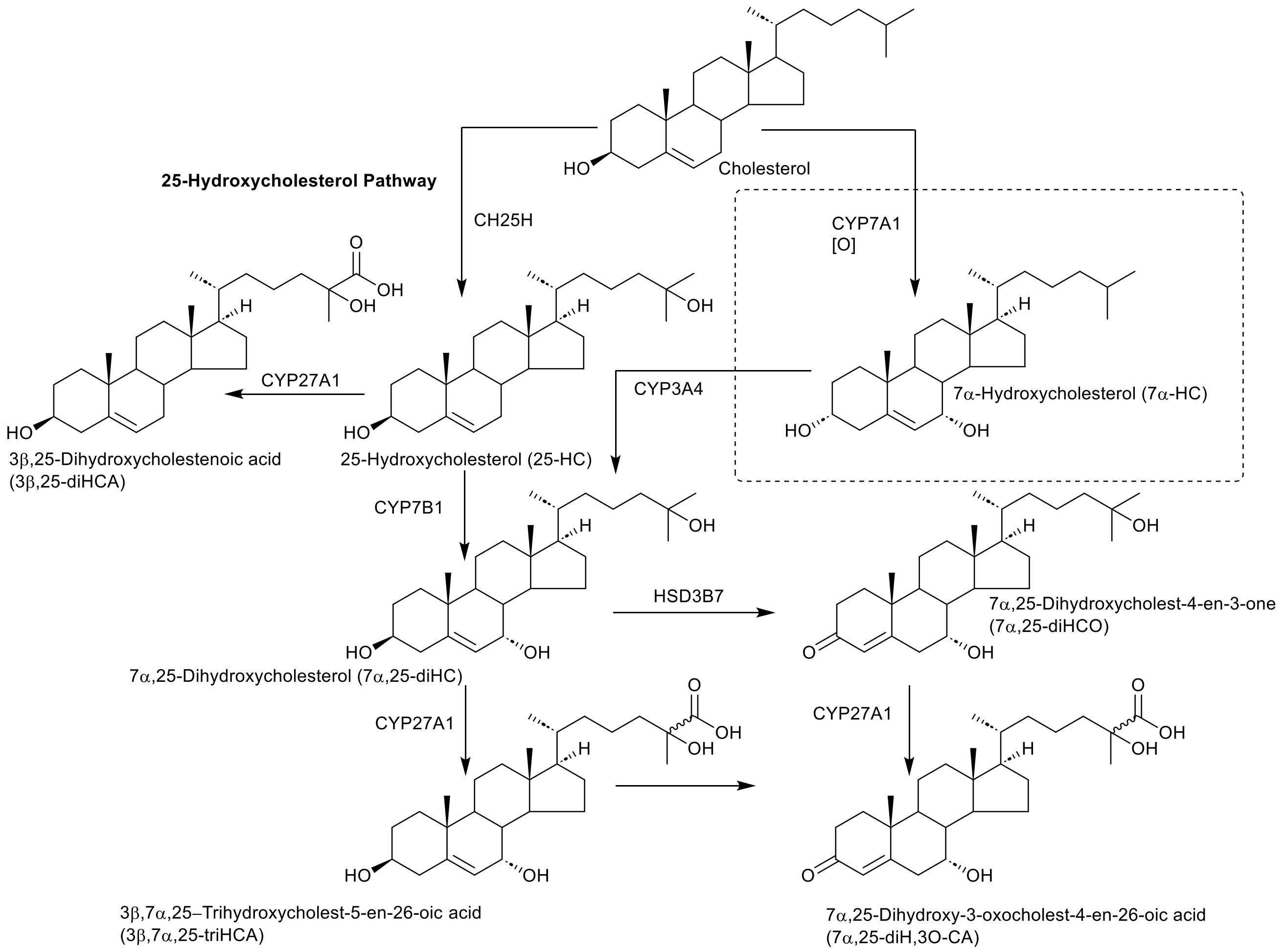

### Figure S5

#### Figure S5A

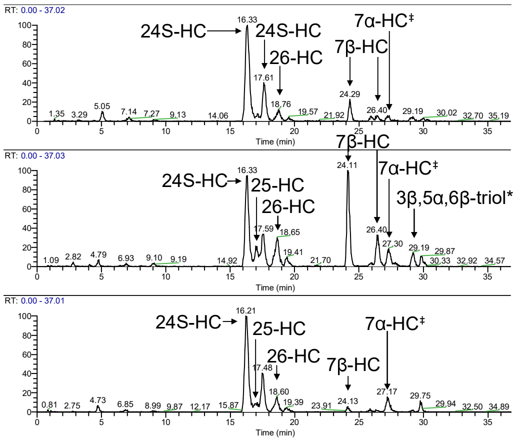


#### Figure S5D

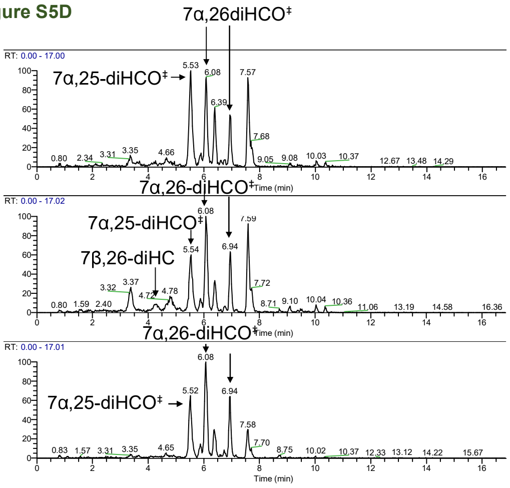


#### Figure S5B

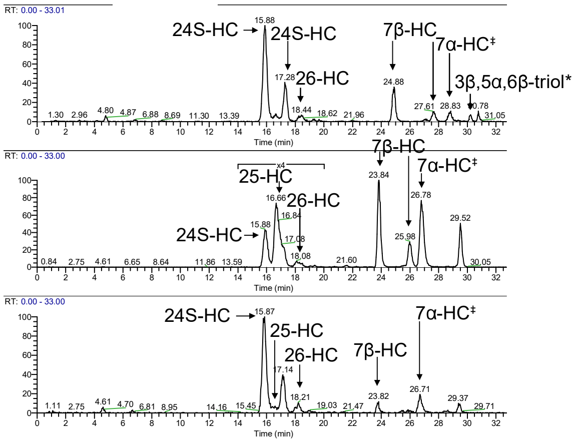


#### Figure S5E

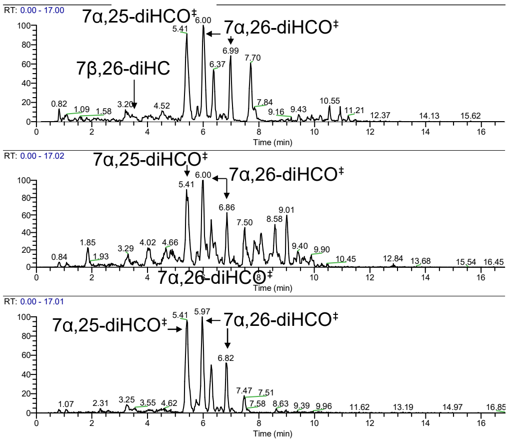


#### Figure S5C

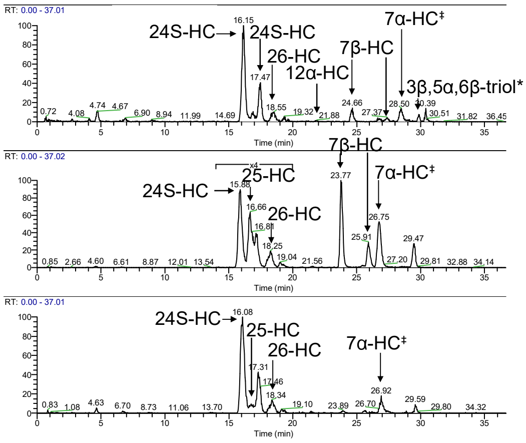


#### Figure S5F

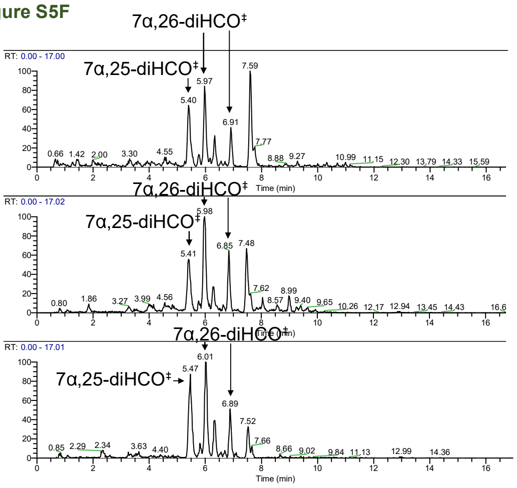

### Figure S6

Figure S6A

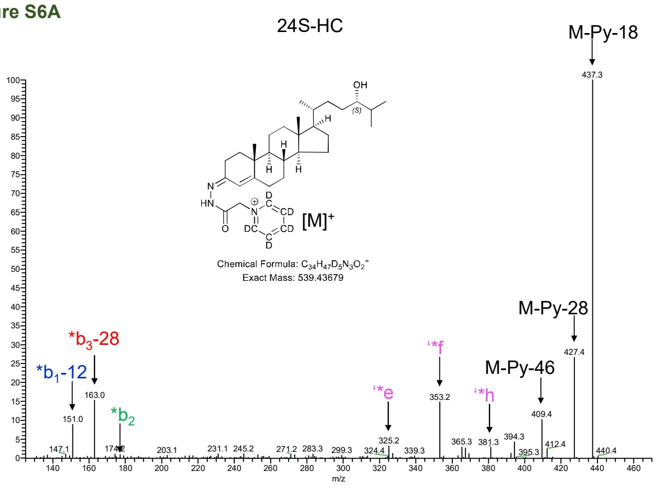


Figure S6D

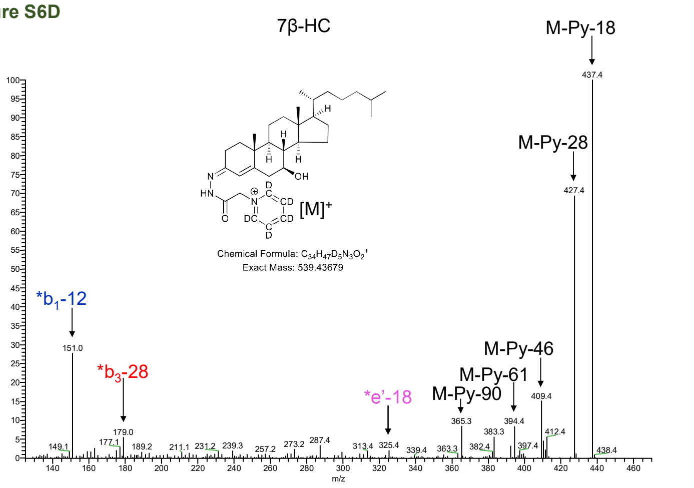


Figure S6B

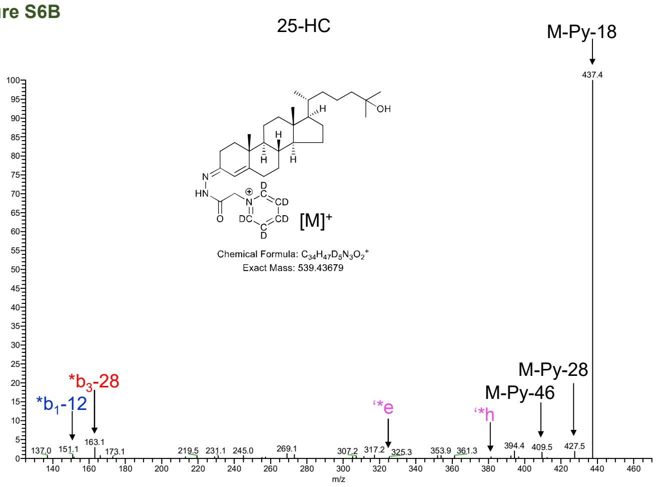


Figure S6E

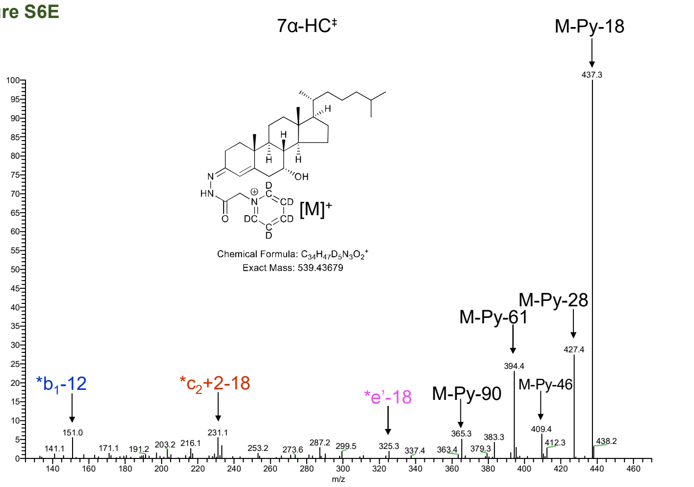


Figure S6C

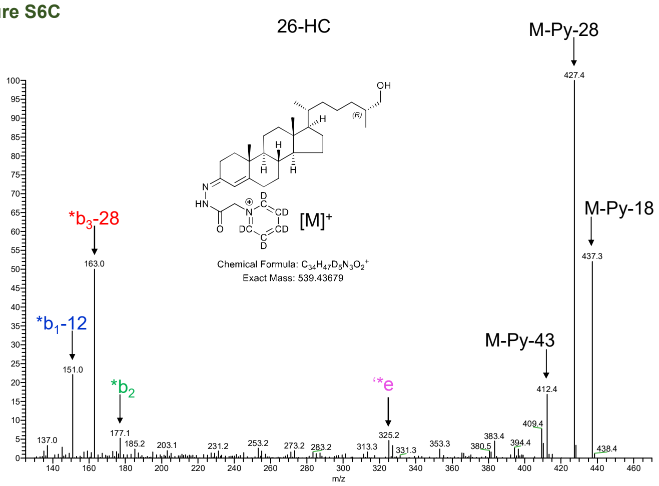


Figure S6F

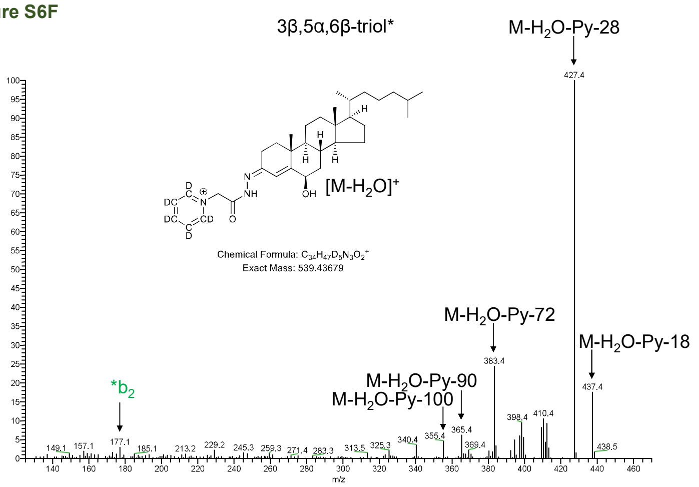

**Figure S6G**

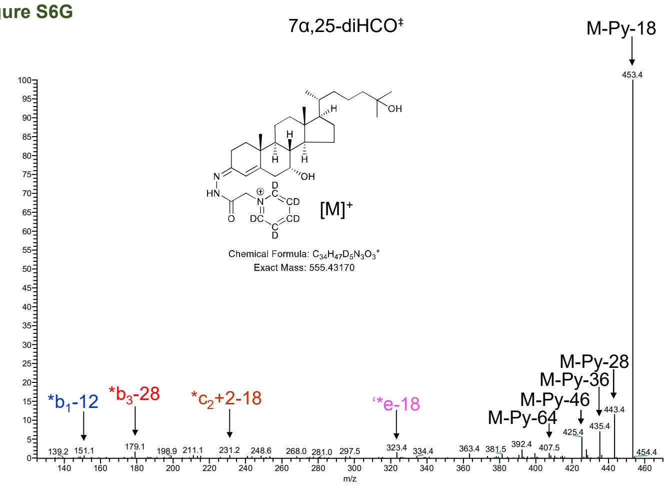


**Figure S6H**

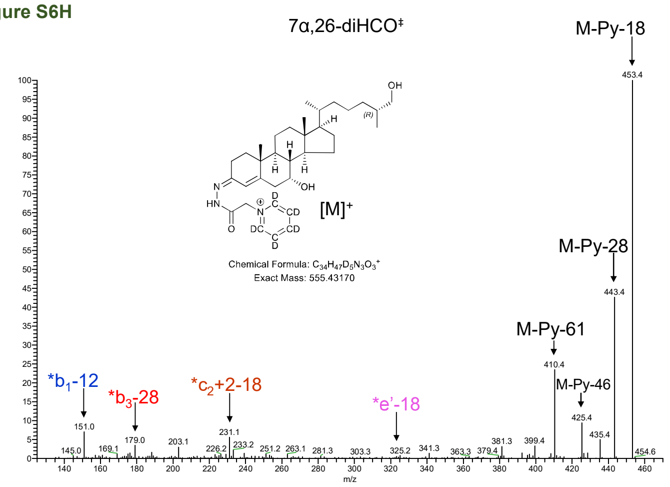


**Figure S6I**

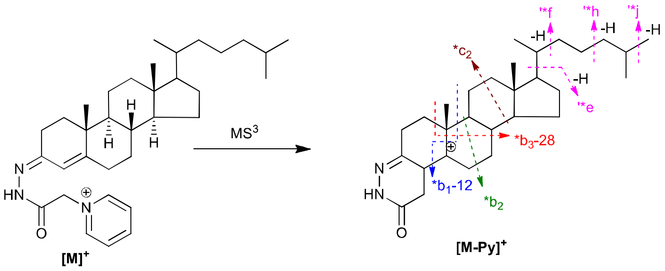

### Figure S7

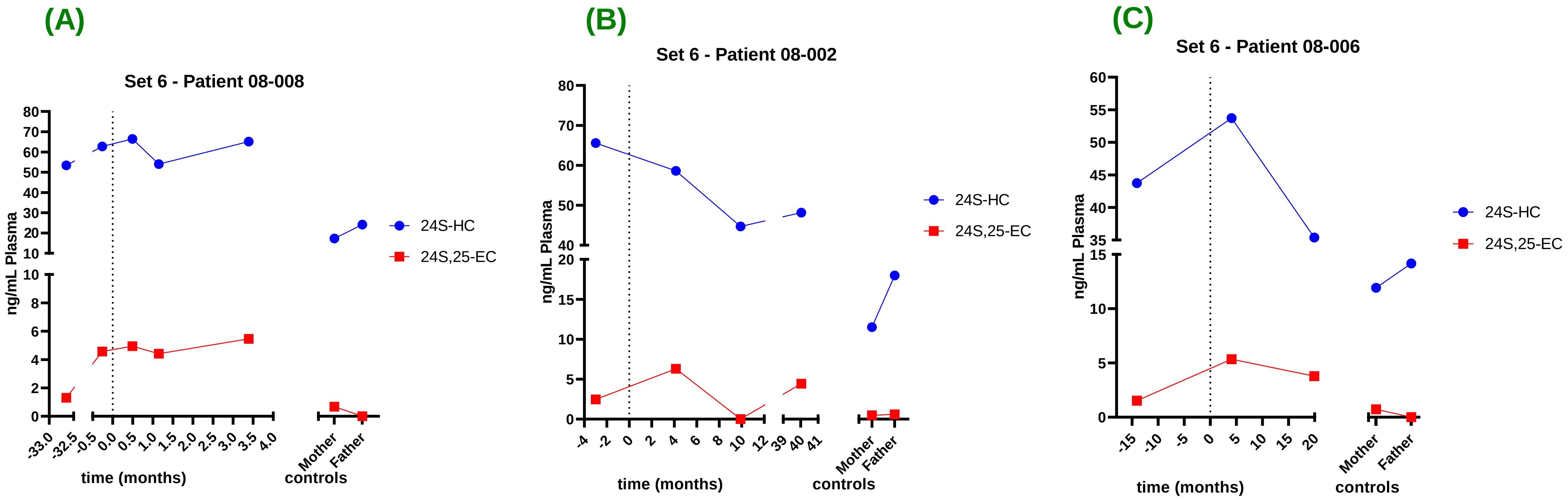

### Figure S8

**Figure S8A**

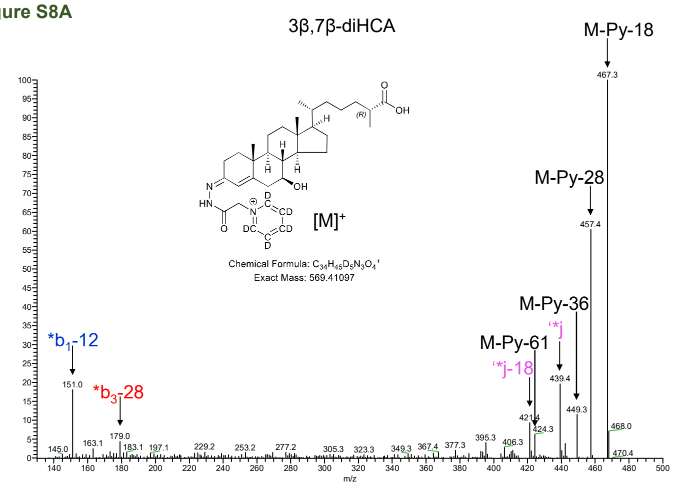


**Figure S8D**

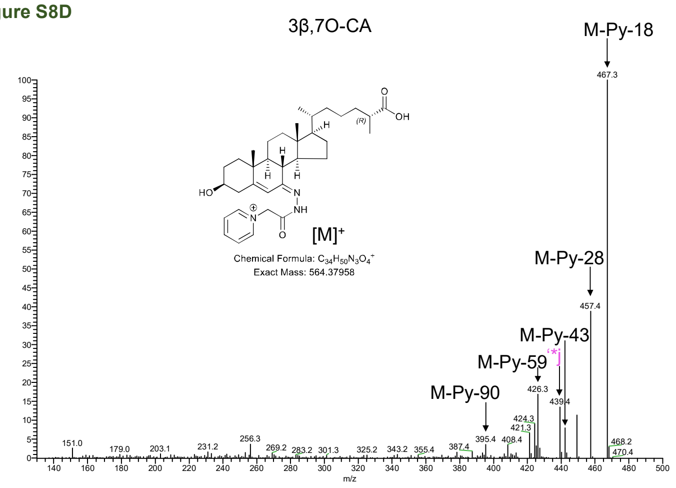


**Figure S8B**

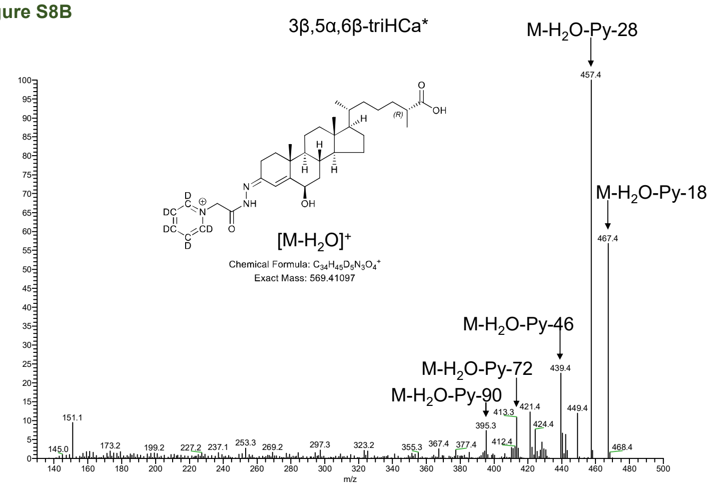


**Figure S8E**

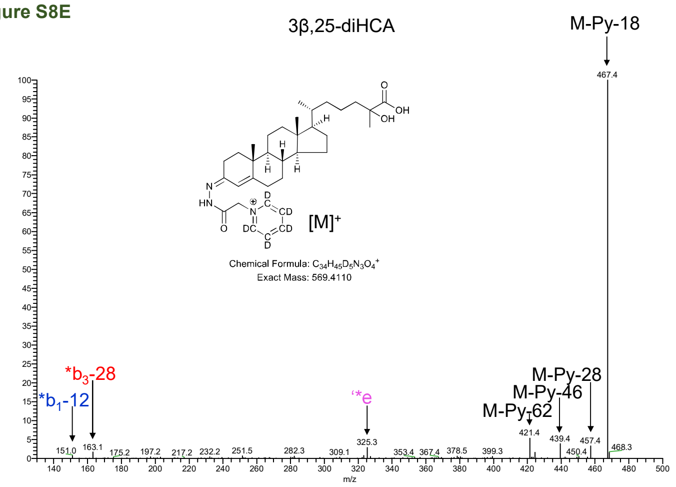


**Figure S8C**

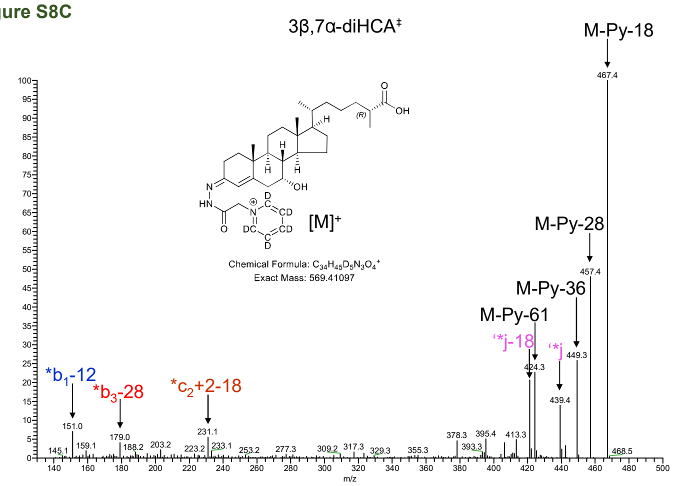

### Figure S9

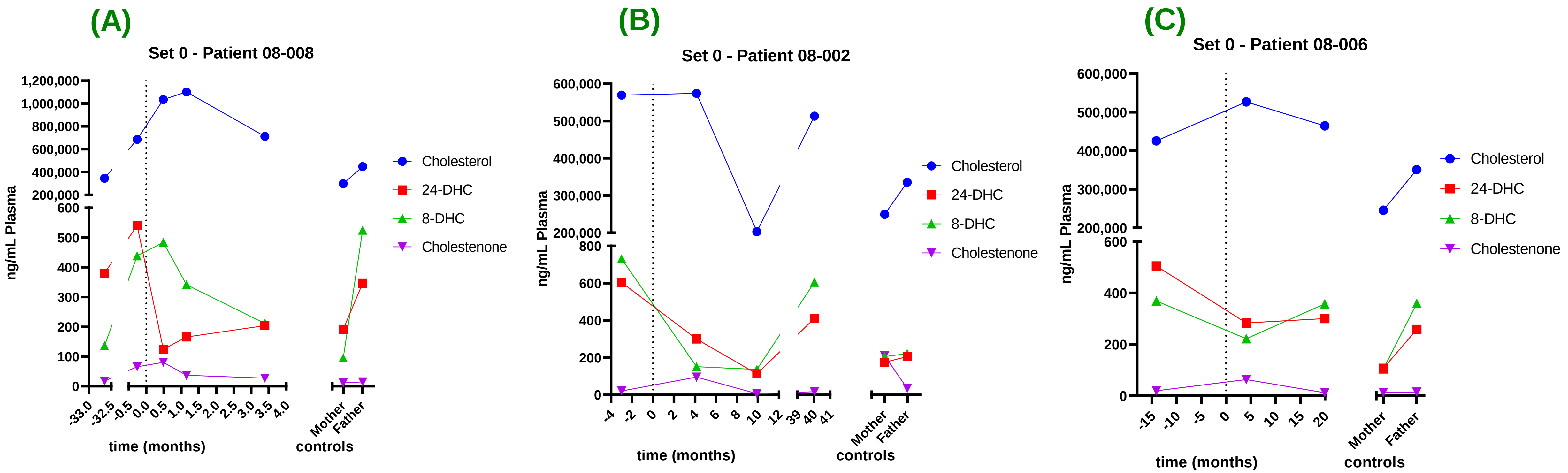
