## Supplementary material for "Deletion of *CH25H* and *LIPA* Genes in Human Abolishes Biosynthesis of 25-Hydroxycholesterol but not of 7α,25-Dihydroxysterols and Enhances Non-enzymatic Cholesterol Oxidation: Metabolic Changes are Partially Reversed by Hematopoietic Stem Cell Transplant^‡^": Figure S2

### Figure S2A/B

**FRACTION A**

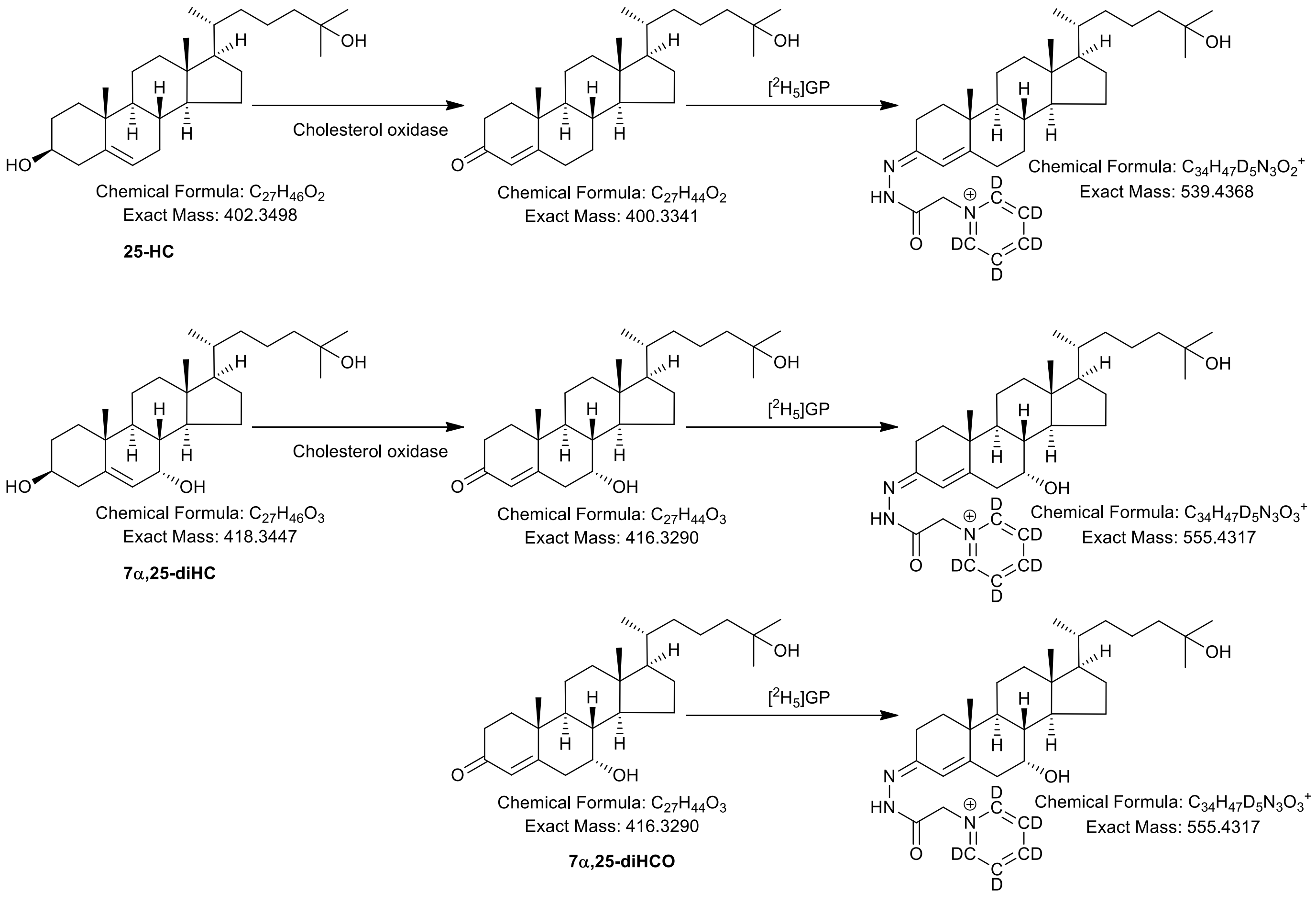


**FRACTION B**

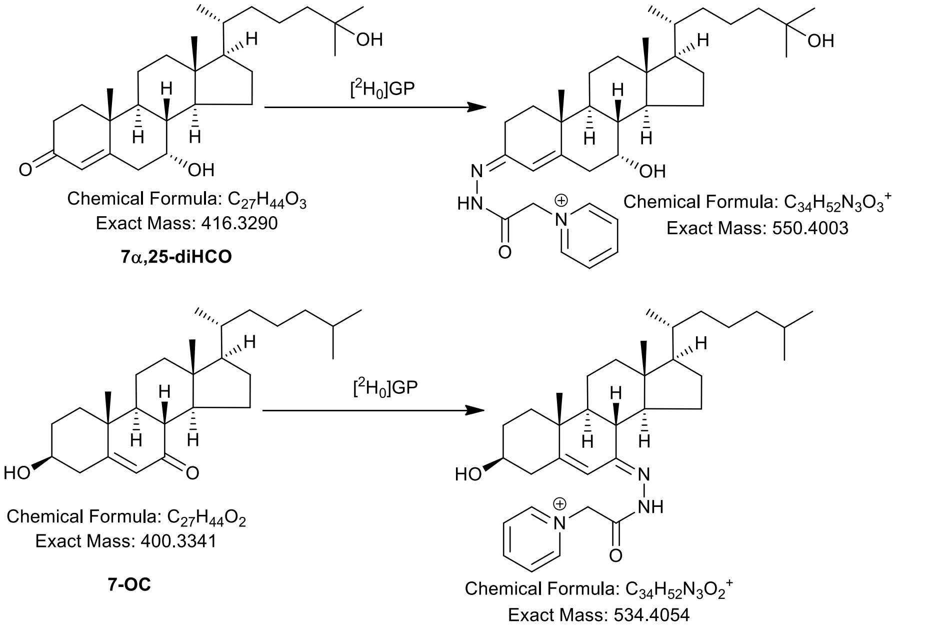

### Figure S2C

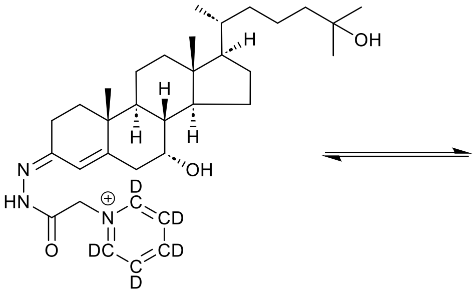

Chemical Formula: $C_{34}H_{47}D_5N_3O_3^+$
Exact Mass: 555.4317

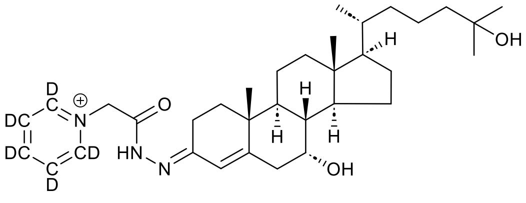

Chemical Formula: $C_{34}H_{47}D_5N_3O_3^+$
Exact Mass: 555.4317

### Figure S2D

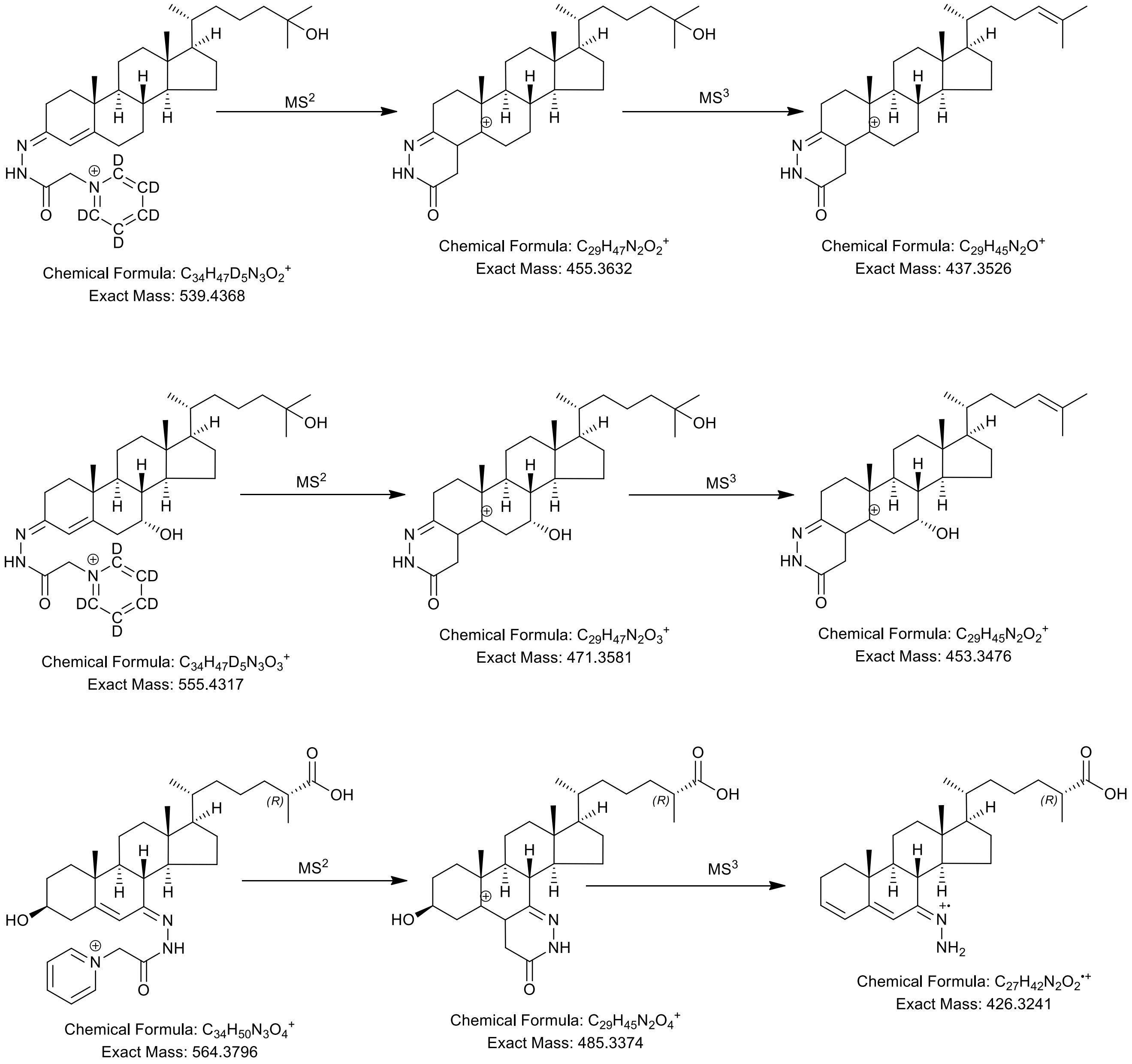
MS$^2$
MS$^3$
Chemical Formula: $C_{34}H_{47}D_5N_3O_2^+$
Exact Mass: 539.4368
Chemical Formula: $C_{29}H_{47}N_2O_2^+$
Exact Mass: 455.3632
Chemical Formula: $C_{29}H_{45}N_2O^+$
Exact Mass: 437.3526
MS$^2$
MS$^3$
Chemical Formula: $C_{34}H_{47}D_5N_3O_3^+$
Exact Mass: 555.4317
Chemical Formula: $C_{29}H_{47}N_2O_3^+$
Exact Mass: 471.3581
Chemical Formula: $C_{29}H_{45}N_2O_2^+$
Exact Mass: 453.3476
MS$^2$
MS$^3$
Chemical Formula: $C_{34}H_{50}N_3O_4^+$
Exact Mass: 564.3796
Chemical Formula: $C_{29}H_{45}N_2O_4^+$
Exact Mass: 485.3374
Chemical Formula: $C_{27}H_{42}N_2O_2^{\bullet+}$
Exact Mass: 426.3241
