## Supplementary material for "Deletion of *CH25H* and *LIPA* Genes in Human Abolishes Biosynthesis of 25-Hydroxycholesterol but not of 7α,25-Dihydroxysterols and Enhances Non-enzymatic Cholesterol Oxidation: Metabolic Changes are Partially Reversed by Hematopoietic Stem Cell Transplant^‡^": Table S1

**Table 1. Oxysterols identified in plasma of patients with the *CH25H-LIPA* homozygous deletion. Oxysterols are colour coded according to the pathway in which they appear. Patients (with anonymised code) are colour coded with their parents according to pre- and post-HSCT.**

| GROUP | Systematic Name (Common name) | Simple Abbreviation | Systematic abbreviation |
|---|---|---|---|
| 0 | Cholest-5-en-3β-ol (Cholesterol) | **C** | C$^{5}$-3β-ol |
| 0 | Cholest-4-en-3-one (Cholestenone) | **CO** | C$^{4}$-3-one |
| 0 | Cholesta-5,24-dien-3β-ol (Desmosterol) | **24-DHC** | C$^{5,24}$-3β-ol |
| 0 | Cholesta-5,8(9)-dien-3β-ol (8-Dehydrocholesterol) | **8-DHC** | C$^{5,8}$-3β-ol |
| 1A | Cholest-5-en-3β,(25R)26-diol (26-Hydroxycholesterol) [11] | **26-HC** | C$^{5}$-3β,(25R)26-diol |
| 1A | (25R)26-Hydroxycholest-4-en-3-one | **26-HCO** | C$^{4}$-(25R)26-ol-3-one |
| 1A | 3β-Hydroxycholesta-5-en-(25R)26-oic acid (Cholestenoic acid) | **3β-HCA** | CA$^{5}$-3β-ol |
| 1A | 3-Oxocholesta-4-en-(25R)26-oic acid ((25R)-Δ$^{4}$-Dafachronic acid | **3O-CA** | CA$^{4}$-3-one |
| 1A | 3β-Hydroxychol-5-en-24-oic acid | **3βH-Δ$^{5}$-BA** | BA$^{5}$-3β-ol |
| 1A | 3β-Hydroxycholesta-5,24-dien-26-oic acid | **3βH-Δ$^{5,24}$-CA** [1] | CA$^{5,24}$-3β-ol |
| 1B/1B | Cholest-5-en-3β,7α-diol (7α-Hydroxycholesterol) | **7α-HC** | C$^{5}$-3β,7α-diol |
| 1B/1B | 7α-Hydroxycholest-4-en-3-one | **7α-HCO** | C$^{4}$-7α-ol-3-one |
| 1B/1B | Cholest-5-en-3β,7α,26-triol (7α,26-Dihydroxycholesterol) [11] | **7α,26-diHC** [2] | C$^{5}$-3β,7α,26-triol [2] |
| 1B/1B | 7α,26-Dihydrocycholest-4-en-3-one | **7α,26-diHCO** [2] | C$^{4}$-7α,26-diol-3-one [2] |
| 1A/1B | 3β,7α-Dihydroxycholest-5-en-26-oic acid | **3β,7α-diHCA** [2] | CA$^{5}$-3β,7α-diol [2] |
| 1A/1B | 7α-Hydroxy-3-oxocholest-4-en-26-oic acid | **7αH,3O-CA** [2] | CA$^{4}$-7α-ol-3-one [2] |
| 1A/1B | 3β,7α,24-Trihydroxycholest-5-en-26-oic acid | **3β,7α,24-triHCA** | CA$^{5}$-3β,7α,24-triol |
| 1A/1B | 7α,24-Dihydroxy-3-oxocholest-4-en-26-oic acid | **7α,24-diH,3O-CA** | CA$^{4}$-7α,24-diol-3-one |
| 1A/1B | 3β,7α-Dihydroxy-27-nor-cholest-5-en-24-one | **7αH,24O-27-nor-C** [1,9] | 27-nor-C$^{5}$-3β,7α-diol-24-one [1,9] |
| 1A/1B | 7α-Hydroxy-27-nor-cholest-4-en-3,24-dione | **7αH-27-nor-C-3,24-diO** [1,9] | 27-nor-C$^{4}$-7α-ol-3,24-dione [1,9] |
| 1A/1B | 3β,7α-Dihydroxychol-5-en-24-oic acid | **3β,7α-diH-Δ$^{5}$-BA** | BA$^{5}$-3β,7α-diol |
| 1A/1B | 7α-Hydroxy-3-oxochol-4-en-24-oic acid | **7αH,3O-Δ$^{4}$-BA** | BA$^{4}$-7α-ol-3-one |
| 1B/1B | Cholest-5-en-3β,12α-diol (12α-Hydroxycholesterol) | **12α-HC** [1] | C$^{5}$-3β,12α-diol [1] |
| 1B/1B | Cholest-5-en-3β,7α,12α-triol (7α,12α-Dihydroxycholesterol) | **7α,12α-diHC** | C$^{5}$-3β,7α,12α-triol |
| 1B/1B | 7α,12α-Dihydrocycholest-4-en-3-one | **7α,12α-diHCO** | C$^{4}$-7α,12α-diol-3-one |
| 1A/1B | 3β,7α,12α-Trihydroxycholest-5-en-26-oic acid | **3β,7α,12α-triHCA** [1] | CA$^{5}$-3β,7α,12α-triol [1] |
| 1A/1B | 7α,12α-Dihydroxy-3-oxocholest-4-en-26-oic acid | **7α,12α-diH,3O-CA** [1] | CA$^{4}$-7α,12α,-diol-3-one [1] |
| 2 | Cholest-5-en-3β,7β-diol (7β-Hydroxycholesterol) | **7β-HC** | C$^{5}$-3β,7β-diol |
| 2 | Cholest-5-en-3β,7β,26-triol (7α,26-Dihydroxycholesterol) [11] | **7β,26-diHC** | C$^{5}$-3β,7β,26-triol |
| 2 | 3β,7β-Dihydroxycholest-5-en-26-oic acid | **3β,7β-diHCA**[2] | CA$^{5}$-3β,7β-diol[2] |
| 2 | 3β,7β,24-Trihydroxycholest-5-en-26-oic acid | **3β,7β,24-triHCA** [1] | CA$^{5}$-3β,7β,24-triol [1] |
| 2 | 3β,7β-Dihydroxychol-5-en-24-oic acid | **3β,7β-diH-Δ$^{5}$-BA** | BA$^{5}$-3β,7β-diol |
| 3 | 3β-Hydroxycholest-5-en-7-one (7-Oxocholesterol) | **7-OC** | C$^{5}$-3β-ol-7-one |
| 3 | 3β,26-Dihydroxycholest-5-en-7-one (26-hydroxy-7-oxocholesterol) [11] | **26H,7O-C** | C$^{5}$-3β,26-diol-7-one |
| 3 | 3β-Hydroxy-7-oxocholest-5-en-26-oic acid | **3βH,7O-CA** | CA$^{5}$-3β-ol-7-one |
| 3 | 3β,24-Dihydroxy-7-oxocholest-5-en-26-oic acid | **3β,24-diH,7O-CA** [1] | CA$^{5}$-3β,24-diol-7-one [1] |
| 3 | 3β-Hydroxy-7-oxochol-5-en-24-oic acid | **3βH,7O-Δ$^{5}$-BA** [1] | BA$^{5}$-3β-ol-7-one [1] |
| 4 | Cholestane-3β,5α,6β-triol | **3β,5α,6β-triol** [10] | C-3β,5α,6β-triol [10] |
| 4 | Cholestane-3β,5α,6β,26-tetrol | **3β,5α,6β,26-tetrol** [1,10] | C-3β,5α,6β,26-tetrol [1,10] |
| 4 | 3β,5α,6β-Trihydroxycholestan-26-oic acid | **3β,5α,6β-triHCa** [1,10] | CA-3β,5α,6β-triol [1,10] |
| 4 | 3β,5α,6β,24-Tetrahydroxycholestan-26-oic acid | **3β,5α,6β,24-tetraHCa** [1] | CA-3β,5α,6β,24-tetrol [1,10] |
| 4 | 3β,5α,6β-Trihydroxycholan-24-oic acid | **3β,5α,6β-triHBA** [1] | BA-3β,5α,6β-triol [1,10] |
| 5 | Cholest-5-en-3β,25-diol (25-Hydroxycholesterol) | **25-HC** | C$^{5}$-3β,25-diol |
| 5 | Cholest-5-en-3β,7α,25-triol (7α,25-Dihydroxycholesterol) | **7α,25-diHC** | C$^{5}$-3β,7α,25-triol |
| 5 | 7α,25-Dihydrocycholest-4-en-3-one | **7α,25-diHCO** | C$^{4}$-7α,25-diol-3-one |
| 5 | 3β,7α,25-Trihydroxycholest-5-en-26-oic acid | **3β,7α,25-triHCA** | CA$^{5}$-3β,7α,25-triol |
| 5 | 7α,25-Dihydroxy-3-oxocholest-4-en-26-oic acid | **7α,25-diH,3O-CA** | CA$^{4}$-7α,25-diol-3-one |
| 5 | 3β,25-Dihydroxycholest-5-en-26-oic acid | **3β,25-diHCA)** [1] | CA$^{5}$-3β,25-diol [1] |
| | 3β,7β,25-Trihydroxycholest-5-en-26-oic acid | **3β,7β,25-triHCA** [1] | CA$^{5}$-3β,7β,25-triol [1] |
| 6 | Cholest-5-en-3β,24S-diol (24S-Hydroxycholesterol) | **24S-diHC** | C$^{5}$-3β,24S-diol |
| 6 | 3β-Hydroxycholest-5-en-24S,25-epoxide (24S,25-Epoxycholesterol) | **24S,25-EC** [3] | C$^{5}$-3β-ol-24,25-epoxide [3] |
| | Cholest-5-en-3β,22R-diol (22R-Hydroxycholesterol) | **22R-HC** | C$^{5}$-3β,22R-diol |
| | Cholest-5-en-3β,24R-diol (24R-Hydroxycholesterol) | **24R-HC** | C$^{5}$-3β,24R-diol |
| 6 | Cholest-5-en-3β,7α,24-triol (7α,24-Dihydroxycholesterol) | **7α,24-diHC** | C$^{5}$-3β,7α,24-triol |
| 6 | 7α,24-Dihydrocycholest-4-en-3-one | **7α,24-diHCO** | C$^{4}$-7α,24-diol-3-one |
| | 3β,22-Dihydroxycholest-5-en-24-one | **22H,24O-C** [1,4] | C$^{5}$-3β,22-diol-24-one [1,4] |
| | 3β,22-Dihydroxycholest-5-en-26-oic acid | **3β,22-diHCA** [1,5] | CA$^{5}$-3β,22-diol [1,5] |
| | 3β,x-Dihydroxycholest-5-en-26-oic acid | **3β,x-diHCA** [1,6] | CA$^{5}$-3β,x-diol [1,6] |
| | 3β,22,25-Trihydroxycholest-5-en-24-one | **22,25-diH,24O-C** [1,7] | C$^{5}$-3β,22,25-triol [1,7] |
| | 3β,x-Dihydroxycholest-5-en-26-oic acid | **3β,x-diHCA** [1,8] | CA$^{5}$-3β,x-diol [1,8] |
| | 3β,7α,x-Trihydroxycholest-5-en-26-oic acid | **3β,7α,x-triHCA** [1] | CA$^{5}$-3β,7α,x-triol [1] |
| | 7α,x-Dihydroxy-3-oxocholest-4-en-26-oic acid | **7α,x-diH,3O-CA** [1] | CA$^{4}$-7α,x-diol-3-one [1] |

| Simple Abbreviation | Patient 08-008 Pre-HSCT 32.65 mo Affected (LALD) Affected(CH25H) ng/mL | Patient 08-008 Pre-HSCT 0.26 mo Affected (LALD) Affected(CH25H) ng/mL | Patient 08-008 Post-HSCT 0.49 mo Affected (LALD) Affected(CH25H) ng/mL | Patient 08-008 Post-HSCT 1.15 mo Affected (LALD) Affected(CH25H) ng/mL | Patient 08-008 Post-HSCT 3.39 mo Affected (LALD) Affected(CH25H) ng/mL | Mom 08-008 Mother HET HET ng/mL | Dad 08-008 Father HET HET ng/mL |
|---|---|---|---|---|---|---|---|
| C | 344039.73 | 685065.81 | 1034384.74 | 1101041.69 | 711728.66 | 297955.59 | 447612.06 |
| CO | 17.76 | 65.43 | 80.43 | 37.19 | 27.41 | 11.60 | 14.67 |
| 24-DHC | 380.27 | 540.26 | 124.72 | 165.77 | 203.59 | 191.80 | 346.63 |
| 8-DHC | 136.16 | 438.04 | 483.96 | 341.46 | 210.85 | 95.03 | 524.77 |
| 26-HC | 20.47 | 24.44 | 84.76 | 76.14 | 44.39 | 22.34 | 45.98 |
| 26-HCO | 0.00 | 0.00 | 0.65 | 1.28 | 0.00 | 0.19 | 0.00 |
| 3β-HCA | 276.44 | 346.61 | 651.20 | 338.14 | 212.26 | 87.97 | 147.38 |
| 3O-CA | 2.37 | 3.37 | 17.61 | 15.33 | 5.04 | 2.34 | 3.49 |
| 3βH-Δ$^{5}$-BA | 12.13 | 10.93 | 20.11 | 10.34 | 6.74 | 2.62 | 4.54 |
| 3βH-Δ$^{5,24}$-CA [1] | 15.09 | 12.06 | 2.14 | 1.26 | 1.74 | 1.93 | 3.27 |
| 7α-HC | 5.66 | 4.68 | 21.80 | 16.07 | 4.56 | 30.18 | 59.26 |
| 7α-HCO | 7.09 | 3.34 | 15.23 | 49.76 | 32.35 | 46.38 | 180.77 |
| 7α,26-diHC [2] | 2.07 | 1.35 | 4.98 | 1.89 | 1.87 | 1.70 | 1.93 |
| 7α,26-diHCO [2] | 9.11 | 16.84 | 27.96 | 22.48 | 33.94 | 7.00 | 14.06 |
| 3β,7α-diHCA [2] | 122.39 | 155.26 | 286.96 | 126.07 | 94.11 | 32.06 | 51.78 |
| 7αH,3O-CA [2] | 126.45 | 158.63 | 341.34 | 297.03 | 260.16 | 88.73 | 208.08 |
| 3β,7α,24-triHCA | 0.84 | 2.90 | 0.00 | 5.65 | 1.73 | 0.21 | 0.13 |
| 7α,24-diH,3O-CA | 1.35 | 1.56 | 1.96 | 0.96 | 0.68 | 0.00 | 0.39 |
| 7αH,24O-27-nor-C [1,9] | 1.01 | 3.84 | 0.91 | 0.93 | 0.49 | 0.48 | 0.21 |
| 7αH-27-nor-C-3,24-diO [1,9] | 2.01 | 0.00 | 2.45 | 2.28 | 2.29 | 0.00 | 0.51 |
| 3β,7α-diH-Δ$^{5}$-BA | 8.93 | 11.77 | 18.59 | 12.82 | 7.60 | 1.97 | 1.95 |
| 7αH,3O-Δ$^{4}$-BA | 10.02 | 20.60 | 19.20 | 20.76 | 16.77 | 5.83 | 15.72 |
| 12α-HC [1] | 0.00 | 0.46 | 1.06 | 0.77 | 0.00 | 0.00 | 0.00 |
| 7α,12α-diHC | 0.00 | 0.00 | 0.31 | 0.82 | 0.16 | 0.00 | 1.10 |
| 7α,12α-diHCO | 0.00 | 0.00 | 0.59 | 0.00 | 0.41 | 1.92 | 0.69 |
| 3β,7α,12α-triHCA [1] | 0.40 | 1.43 | 13.85 | 4.89 | 0.89 | 0.21 | 0.00 |
| 7α,12α-diH,3O-CA [1] | 3.09 | 2.41 | 8.21 | 8.40 | 5.23 | 5.98 | 14.84 |
| 7β-HC | 7.34 | 27.33 | 233.78 | 41.80 | 12.64 | 0.00 | 0.00 |
| 7β,26-diHC | 0.00 | 1.39 | 3.92 | 1.06 | 0.00 | 0.00 | 0.00 |
| 3β,7β-diHCA[2] | 101.47 | 193.32 | 1671.24 | 250.08 | 81.72 | 6.22 | 11.23 |
| 3β,7β,24-triHCA [1] | 3.61 | 8.36 | 27.18 | 14.92 | 3.88 | 0.00 | 0.00 |
| 3β,7β-diH-Δ$^{5}$-BA | 9.04 | 50.05 | 124.35 | 48.58 | 16.75 | 1.45 | 2.11 |
| 7-OC | 36.87 | 230.55 | 553.64 | 217.02 | 55.88 | 8.91 | 0.00 |
| 26H,7O-C | 1.51 | 1.75 | 10.24 | 6.35 | 1.86 | 0.00 | 0.00 |
| 3βH,7O-CA | 88.05 | 58.07 | 193.05 | 107.77 | 26.57 | 0.00 | 5.10 |
| 3β,24-diH,7O-CA [1] | 3.39 | 6.28 | 8.88 | 9.35 | 3.57 | 0.00 | 0.31 |
| 3βH,7O-Δ$^{5}$-BA [1] | 6.49 | 5.54 | 7.96 | 7.79 | 1.73 | 0.25 | 0.00 |
| 3β,5α,6β-triol [10] | 39.65 | 69.33 | 524.53 | 99.57 | 82.81 | 1.81 | 0.00 |
| 3β,5α,6β,26-tetrol [1,10] | 0.43 | 0.89 | 4.28 | 2.17 | 0.71 | 0.00 | 0.00 |
| 3β,5α,6β-triHCa [1,10] | 20.15 | 56.88 | 247.70 | 102.18 | 34.50 | 0.00 | 0.00 |
| 3β,5α,6β,24-tetraHCa [1] | 4.27 | 9.38 | 58.72 | 23.94 | 4.54 | 0.37 | 0.00 |
| 3β,5α,6β-triHBA [1] | 0.91 | 4.47 | 5.62 | 3.92 | 0.79 | 0.00 | 0.00 |
| 25-HC | 0.00 | 0.00 | 5.76 | 0.81 | 1.62 | 1.02 | 1.81 |
| 7α,25-diHC | 0.95 | 0.41 | 0.48 | 0.19 | 0.62 | 0.58 | 0.18 |
| 7α,25-diHCO | 3.12 | 3.17 | 1.82 | 3.06 | 3.41 | 1.61 | 1.95 |
| 3β,7α,25-triHCA | 1.54 | 2.06 | 8.71 | 4.63 | 3.10 | 0.08 | 0.25 |
| 7α,25-diH,3O-CA | 2.55 | 3.77 | 7.61 | 4.89 | 1.31 | 0.33 | 0.87 |
| 3β,25-diHCA) [1] | 0.90 | 2.00 | 1.47 | 1.49 | 2.73 | 16.44 | 10.97 |
| 3β,7β,25-triHCA [1] | 0.57 | 2.01 | 7.69 | 4.82 | 0.98 | 0.00 | 0.00 |
| 24S-diHC | 53.44 | 62.72 | 66.46 | 54.03 | 65.14 | 17.26 | 24.15 |
| 24S,25-EC [3] | 1.31 | 4.57 | 4.94 | 4.41 | 5.46 | 0.68 | 0.00 |
| 22R-HC | 0.00 | 0.00 | 0.00 | 1.75 | 0.00 | 0.00 | 0.00 |
| 24R-HC | DNQ | 0.00 | DNQ | 0.00 | DNQ | DNQ | DNQ |
| 7α,24-diHC | DNQ | DNQ | DNQ | DNQ | DNQ | DNQ | 0.00 |
| 7α,24-diHCO | DNQ | DNQ | DNQ | 0.00 | 0.00 | 0.00 | 0.00 |
| 22H,24O-C [1,4] | 0.00 | 0.90 | 0.45 | 0.00 | 0.00 | 26.31 | 25.19 |
| 3β,22-diHCA [1,5] | 5.35 | 1.88 | 3.30 | 1.65 | 0.00 | 2.79 | 2.97 |
| 3β,x-diHCA [1,6] | 4.50 | 9.97 | 0.00 | 4.60 | 2.96 | 0.53 | 0.58 |
| 22,25-diH,24O-C [1,7] | 0.00 | 0.00 | 0.00 | 0.00 | 0.00 | 6.34 | 5.57 |
| 3β,x-diHCA [1,8] | 4.66 | 0.00 | 21.51 | 2.25 | 2.38 | 2.78 | 1.02 |
| 3β,7α,x-triHCA [1] | 3.83 | 8.30 | 6.92 | 8.49 | 4.53 | 0.43 | 0.94 |
| 7α,x-diH,3O-CA [1] | 9.45 | 10.06 | 20.06 | 9.12 | 9.50 | 1.62 | 2.52 |

| Simple Abbreviation | Patient 08-002 Pre-HSCT 2.99 mo Affected (LALD) Affected(CH25H) ng/mL | Patient 08-002 Post-HSCT 4.14 mo Affected (LALD) Affected(CH25H) ng/mL | Patient 08-002 Post-HSCT 9.9 mo Affected (LALD) Affected(CH25H) ng/mL | Patient 08-002 Post-HSCT 40.04 mo Affected (LALD) Affected(CH25H) ng/mL | Dad 08-002 Father HET HET ng/mL | Mom 08-002 Mother HET HET ng/mL |
|---|---|---|---|---|---|---|
| C | 569646.36 | 574385.13 | 202878.33 | 513146.72 | 335390.21 | 249350.38 |
| CO | 20.93 | 95.96 | 7.07 | 17.19 | 35.20 | 209.53 |
| 24-DHC | 604.08 | 299.95 | 113.27 | 411.08 | 205.82 | 175.22 |
| 8-DHC | 730.17 | 151.44 | 136.38 | 605.16 | 221.59 | 205.81 |
| 26-HC | 18.05 | 16.02 | 11.48 | 16.54 | 20.19 | 17.29 |
| 26-HCO | 0.00 | 0.00 | 0.22 | 0.14 | 0.18 | 0.00 |
| 3β-HCA | 130.90 | 132.64 | 78.88 | 79.00 | 72.28 | 90.88 |
| 3O-CA | 1.27 | 1.16 | 0.00 | 1.69 | 1.31 | 0.91 |
| 3βH-Δ$^{5}$-BA | 3.02 | 4.19 | 2.07 | 2.41 | 2.22 | 1.94 |
| 3βH-Δ$^{5,24}$-CA [1] | 7.09 | 2.55 | 1.40 | 2.38 | 1.77 | 1.17 |
| 7α-HC | 11.07 | 1067.33 | **3.37** | 8.41 | 10.71 | 12.46 |
| 7α-HCO | 6.53 | 12.84 | 10.43 | 18.61 | 12.31 | 25.16 |
| 7α,26-diHC [2] | 0.73 | 1.32 | 1.57 | 4.03 | 1.85 | 0.40 |
| 7α,26-diHCO [2] | 7.03 | 10.01 | 9.54 | 15.99 | 3.26 | 6.20 |
| 3β,7α-diHCA [2] | 70.51 | 122.27 | 62.81 | 68.21 | 38.95 | 29.50 |
| 7αH,3O-CA [2] | 70.87 | 51.41 | 74.21 | 80.48 | 43.88 | 73.03 |
| 3β,7α,24-triHCA | 1.91 | 1.17 | 0.12 | 0.26 | 0.42 | 0.31 |
| 7α,24-diH,3O-CA | 0.75 | 0.34 | 0.26 | 0.30 | 0.04 | 0.06 |
| 7αH,24O-27-nor-C [1,9] | 1.16 | 0.15 | 0.26 | 0.16 | 0.30 | 0.22 |
| 7αH-27-nor-C-3,24-diO [1,9] | 0.76 | 1.35 | 0.92 | 1.21 | 0.24 | 0.30 |
| 3β,7α-diH-Δ$^{5}$-BA | 18.38 | 11.93 | 1.64 | 5.54 | 3.43 | 2.98 |
| 7αH,3O-Δ$^{4}$-BA | 2.98 | 3.84 | 4.52 | 6.80 | 1.88 | 3.98 |
| 12α-HC [1] | 0.60 | 13.01 | 0.00 | 0.57 | 0.42 | 0.49 |
| 7α,12α-diHC | 0.00 | 0.00 | 0.00 | 0.00 | 0.00 | 0.16 |
| 7α,12α-diHCO | 0.00 | 0.00 | 0.00 | 0.28 | 0.00 | 0.27 |
| 3β,7α,12α-triHCA [1] | 0.00 | 0.71 | 0.00 | 0.47 | 0.40 | 0.00 |
| 7α,12α-diH,3O-CA [1] | 2.23 | 1.20 | 0.82 | 1.59 | 1.08 | 3.66 |
| 7β-HC | 45.39 | 1276.38 | 3.26 | 11.79 | 2.15 | 1.26 |
| 7β,26-diHC | 0.69 | 0.00 | 0.00 | 0.00 | 0.25 | 0.00 |
| 3β,7β-diHCA[2] | 145.73 | 125.38 | 11.85 | 49.25 | 4.09 | 6.52 |
| 3β,7β,24-triHCA [1] | 4.30 | 2.66 | 0.00 | 0.45 | 0.00 | 0.00 |
| 3β,7β-diH-Δ$^{5}$-BA | 37.07 | 39.00 | 1.75 | 8.63 | 1.94 | 1.76 |
| 7-OC | 234.58 | 1878.18 | 11.62 | 66.52 | 4.62 | 4.57 |
| 26H,7O-C | 1.98 | 1.49 | 0.65 | 1.36 | 0.00 | 0.00 |
| 3βH,7O-CA | 54.85 | 32.77 | 16.15 | 7.81 | 0.00 | 0.00 |
| 3β,24-diH,7O-CA [1] | 5.21 | 0.67 | 0.59 | 0.00 | 0.00 | 0.00 |
| 3βH,7O-Δ$^{5}$-BA [1] | 3.05 | 1.76 | 0.46 | 0.79 | 0.07 | 0.12 |
| 3β,5α,6β-triol [10] | 65.23 | 68.95 | 33.24 | 26.53 | 2.50 | 0.00 |
| 3β,5α,6β,26-tetrol [1,10] | 0.72 | 0.00 | 0.00 | 0.12 | 0.00 | 0.00 |
| 3β,5α,6β-triHCa [1,10] | 56.01 | 38.11 | 0.00 | 0.00 | 0.00 | 0.00 |
| 3β,5α,6β,24-tetraHCa [1] | 12.63 | 4.18 | 0.00 | 0.00 | 0.00 | 0.00 |
| 3β,5α,6β-triHBA [1] | 8.82 | 1.26 | 0.00 | 0.50 | 0.00 | 0.00 |
| 25-HC | 0.00 | 23.08 | 0.17 | 1.06 | 0.84 | 0.69 |
| 7α,25-diHC | 0.06 | 0.78 | 0.84 | 1.11 | 0.68 | 0.26 |
| 7α,25-diHCO | 2.59 | 2.98 | 3.99 | 3.29 | 0.76 | 1.47 |
| 3β,7α,25-triHCA | 0.00 | 1.33 | 0.37 | 0.80 | 0.08 | 0.03 |
| 7α,25-diH,3O-CA | 1.25 | 0.63 | 0.33 | 0.50 | 0.13 | 0.29 |
| 3β,25-diHCA) [1] | 9.75 | 7.43 | 3.08 | 7.49 | 12.20 | 17.25 |
| 3β,7β,25-triHCA [1] | 0.00 | 0.84 | 0.00 | 0.17 | 0.00 | 0.00 |
| 24S-diHC | 65.58 | 58.64 | 44.72 | 48.16 | 17.98 | 11.52 |
| 24S,25-EC [3] | 2.48 | 6.30 | 0.00 | 4.43 | 0.61 | 0.49 |
| 22R-HC | 0.57 | 1.36 | 0.00 | 0.00 | 0.00 | 0.00 |
| 24R-HC | DNQ | DNQ | DNQ | DNQ | DNQ | DNQ |
| 7α,24-diHC | DNQ | DNQ | 0.00 | DNQ | DNQ | 0.00 |
| 7α,24-diHCO | DNQ | 0.00 | 0.00 | DNQ | 0.00 | 0.00 |
| 22H,24O-C [1,4] | 9.51 | 1.58 | 3.91 | 8.84 | 20.16 | 21.84 |
| 3β,22-diHCA [1,5] | 2.19 | 4.91 | 3.29 | 4.96 | 2.64 | 4.49 |
| 3β,x-diHCA [1,6] | 0.00 | 1.53 | 0.55 | 0.76 | 0.00 | 0.00 |
| 22,25-diH,24O-C [1,7] | 0.00 | 0.00 | 3.21 | 12.48 | 8.96 | 15.91 |
| 3β,x-diHCA [1,8] | 0.00 | 3.81 | 2.70 | 2.14 | 0.00 | 0.00 |
| 3β,7α,x-triHCA [1] | 3.36 | 0.33 | 0.70 | 1.00 | 0.61 | 0.18 |
| 7α,x-diH,3O-CA [1] | 1.94 | 2.97 | 1.48 | 2.24 | 0.77 | 1.15 |

| Simple Abbreviation | Patient 08-006 Pre-HCST 14.10 mo Affected (LALD) Affected(CH25H) ng/mL | Patient 08-006 Post-HSCT 4.08 mo Affected (LALD) Affected(CH25H) ng/mL | Patient 08-006 Post-HSCT 21.99 mo Affected (LALD) Affected(CH25H) ng/mL | Mom 08-006 Mother HET HET ng/mL | Dad 08-006 Father HET HET ng/mL |
|---|---|---|---|---|---|
| C | 425742.65 | 526636.88 | 464411.83 | 245666.59 | 350756.19 |
| CO | 19.82 | 63.65 | 11.93 | 13.13 | 15.29 |
| 24-DHC | 504.88 | 283.20 | 300.84 | 107.22 | 258.28 |
| 8-DHC | 369.05 | 222.15 | 357.91 | 103.98 | 359.51 |
| 26-HC | 17.83 | 43.64 | 19.56 | 21.38 | 31.62 |
| 26-HCO | 0.00 | 0.00 | 0.19 | 0.38 | 0.00 |
| 3β-HCA | 271.39 | 385.80 | 108.05 | 117.38 | 143.04 |
| 3O-CA | 3.14 | 4.41 | 1.85 | 3.04 | 2.85 |
| 3βH-Δ$^{5}$-BA | 8.55 | 12.73 | 5.52 | 2.61 | 3.67 |
| 3βH-Δ$^{5,24}$-CA [1] | 21.41 | 8.84 | 2.47 | 2.15 | 0.80 |
| 7α-HC | 13.96 | 337.98 | 4.11 | 0.24 | 13.40 |
| 7α-HCO | 7.15 | 9.74 | 13.29 | 21.58 | 85.78 |
| 7α,26-diHC [2] | 1.47 | 0.67 | 1.69 | 0.52 | 1.65 |
| 7α,26-diHCO [2] | 6.49 | 31.32 | 13.05 | 5.81 | 9.86 |
| 3β,7α-diHCA [2] | 109.01 | 170.68 | 63.92 | 26.42 | 42.40 |
| 7αH,3O-CA [2] | 122.85 | 162.20 | 79.17 | 78.56 | 153.70 |
| 3β,7α,24-triHCA | 3.82 | 1.47 | 0.19 | 0.00 | 0.14 |
| 7α,24-diH,3O-CA | 1.63 | 0.72 | 0.19 | 0.09 | 0.11 |
| 7αH,24O-27-nor-C [1,9] | 1.05 | 0.00 | 0.00 | 0.15 | 0.24 |
| 7αH-27-nor-C-3,24-diO [1,9] | 1.40 | 1.67 | 1.26 | 0.09 | 0.32 |
| 3β,7α-diH-Δ$^{5}$-BA | 10.65 | 16.66 | 3.70 | 1.64 | 1.63 |
| 7αH,3O-Δ$^{4}$-BA | 12.66 | 11.73 | 1.33 | 4.50 | 10.49 |
| 12α-HC [1] | 0.09 | 0.00 | 0.72 | 0.07 | 0.00 |
| 7α,12α-diHC | 0.00 | 0.00 | 0.07 | 0.02 | 0.00 |
| 7α,12α-diHCO | 0.00 | 0.00 | 0.07 | 0.11 | 0.00 |
| 3β,7α,12α-triHCA [1] | 0.00 | 1.49 | 0.43 | 0.01 | 0.12 |
| 7α,12α-diH,3O-CA [1] | 4.69 | 2.89 | 0.86 | 2.25 | 8.10 |
| 7β-HC | 19.95 | 566.67 | 2.54 | 0.00 | 0.00 |
| 7β,26-diHC | 0.00 | 2.22 | 0.00 | 0.00 | 0.00 |
| 3β,7β-diHCA[2] | 200.69 | 155.45 | 15.88 | 8.35 | 11.06 |
| 3β,7β,24-triHCA [1] | 11.88 | 4.58 | 0.00 | 0.00 | 0.00 |
| 3β,7β-diH-Δ$^{5}$-BA | 35.27 | 36.97 | 2.17 | 1.65 | 1.68 |
| 7-OC | 42.41 | 627.38 | 18.36 | 1.67 | 0.00 |
| 26H,7O-C | 1.55 | 5.15 | 0.00 | 0.00 | 0.00 |
| 3βH,7O-CA | 144.83 | 77.74 | 4.64 | 2.36 | 7.43 |
| 3β,24-diH,7O-CA [1] | 12.62 | 4.39 | 0.34 | 0.46 | 0.00 |
| 3βH,7O-Δ$^{5}$-BA [1] | 6.73 | 5.32 | 0.64 | 0.00 | 0.00 |
| 3β,5α,6β-triol [10] | 59.52 | 58.13 | 13.46 | 0.00 | 0.00 |
| 3β,5α,6β,26-tetrol [1,10] | 0.88 | 0.00 | 0.03 | 0.00 | 0.00 |
| 3β,5α,6β-triHCa [1,10] | 50.86 | 60.46 | 0.00 | 0.00 | 0.00 |
| 3β,5α,6β,24-tetraHCa [1] | 14.18 | 7.10 | 0.00 | 0.00 | 0.00 |
| 3β,5α,6β-triHBA [1] | 1.14 | 1.90 | 0.48 | 0.00 | 0.00 |
| 25-HC | 0.00 | 7.77 | 0.98 | 0.64 | 0.53 |
| 7α,25-diHC | 0.21 | 0.69 | 0.21 | 0.26 | 0.29 |
| 7α,25-diHCO | 2.37 | 3.93 | 2.79 | 1.43 | 1.65 |
| 3β,7α,25-triHCA | 2.58 | 1.61 | 0.06 | 0.08 | 0.29 |
| 7α,25-diH,3O-CA | 2.87 | 1.41 | 0.59 | 0.22 | 0.48 |
| 3β,25-diHCA) [1] | 5.75 | 3.05 | 3.74 | 9.72 | 6.91 |
| 3β,7β,25-triHCA [1] | 0.70 | 1.67 | 0.00 | 0.00 | 0.00 |
| 24S-diHC | 43.74 | 53.72 | 35.39 | 11.93 | 14.17 |
| 24S,25-EC [3] | 1.53 | 5.35 | 3.78 | 0.73 | 0.00 |
| 22R-HC | 0.00 | 0.00 | 0.00 | 0.00 | 0.00 |
| 24R-HC | DNQ | DNQ | DNQ | DNQ | DNQ |
| 7α,24-diHC | DNQ | DNQ | DNQ | 0.00 | 0.00 |
| 7α,24-diHCO | DNQ | DNQ | 0.00 | 0.00 | 0.00 |
| 22H,24O-C [1,4] | 15.05 | 3.61 | 5.28 | 16.09 | 12.65 |
| 3β,22-diHCA [1,5] | 2.88 | 1.36 | 4.01 | 3.48 | 3.12 |
| 3β,x-diHCA [1,6] | 6.50 | 8.08 | 0.71 | 0.74 | 0.67 |
| 22,25-diH,24O-C [1,7] | 0.00 | 0.00 | 4.59 | 3.04 | 3.82 |
| 3β,x-diHCA [1,8] | 3.64 | 2.23 | 2.37 | 0.88 | 0.44 |
| 3β,7α,x-triHCA [1] | 2.61 | 3.60 | 0.86 | 0.06 | 0.00 |
| 7α,x-diH,3O-CA [1] | 13.48 | 6.19 | 1.87 | 1.11 | 2.72 |

| Simple Abbreviation | 200820_NIST_QC [12] ng/mL | NISTSRM1950 [13] ng/mL | NISTSRM1950 [14] ng/mL | NISTSRM1950 [15] ng/mL |
|---|---|---|---|---|
| C | NM | 402515.01 | 403510.83 | 410815.71 |
| CO | NM | NM | NM | NM |
| 24-DHC | NM | 235.40 | NM | NM |
| 8-DHC | NM | 428.55 | NM | NM |
| 26-HC | 22.56 | 24.11 | 22.87 | 22.44 |
| 26-HCO | 0.28 | NM | NM | NM |
| 3β-HCA | 116.38 | 124.08 | 112.66 | 114.50 |
| 3O-CA | 2.45 | NM | NM | NM |
| 3βH-Δ$^{5}$-BA | 4.05 | 3.96 | 2.74 | 2.69 |
| 3βH-Δ$^{5,24}$-CA [1] | 1.83 | NM | NM | NM |
| 7α-HC | 12.59 | 21.44 | 10.39 | 11.24 |
| 7α-HCO | 38.46 | 25.96 | 28.68 | 26.52 |
| 7α,26-diHC [2] | 0.84 | NM | 1.10 | 1.17 |
| 7α,26-diHCO [2] | 9.05 | 7.70 | 8.67 | 8.88 |
| 3β,7α-diHCA [2] | 45.12 | 41.57 | 51.66 | 53.40 |
| 7αH,3O-CA [2] | 91.31 | 99.13 | 122.61 | 120.01 |
| 3β,7α,24-triHCA | 0.03 | NM | NM | NM |
| 7α,24-diH,3O-CA | 0.04 | NM | NM | NM |
| 7αH,24O-27-nor-C [1,9] | 0.00 | NM | NM | NM |
| 7αH-27-nor-C-3,24-diO [1,9] | 0.57 | 0.72 | NM | NM |
| 3β,7α-diH-Δ$^{5}$-BA | 4.86 | 3.84 | 2.19 | 3.32 |
| 7αH,3O-Δ$^{4}$-BA | 5.94 | 5.90 | 5.97 | 6.47 |
| 12α-HC [1] | 0.00 | NM | NM | NM |
| 7α,12α-diHC | 0.26 | NM | 0.11 | 0.15 |
| 7α,12α-diHCO | 0.17 | NM | 0.46 | 0.48 |
| 3β,7α,12α-triHCA [1] | 0.66 | NM | 0.25 | 0.35 |
| 7α,12α-diH,3O-CA [1] | 2.72 | NM | 3.33 | 3.82 |
| 7β-HC | 3.45 | 2.96 | 3.36 | 3.61 |
| 7β,26-diHC | 0.00 | NM | NM | NM |
| 3β,7β-diHCA[2] | 9.65 | 9.22 | 12.25 | 12.57 |
| 3β,7β,24-triHCA [1] | 0.00 | NM | NM | NM |
| 3β,7β-diH-Δ$^{5}$-BA | 1.97 | 2.29 | 2.92 | 4.15 |
| 7-OC | 4.92 | 9.27 | 5.23 | 5.21 |
| 26H,7O-C | 0.00 | NM | NM | NM |
| 3βH,7O-CA | 0.00 | NM | NM | NM |
| 3β,24-diH,7O-CA [1] | 0.00 | NM | NM | NM |
| 3βH,7O-Δ$^{5}$-BA [1] | 0.00 | NM | 0.38 | 0.39 |
| 3β,5α,6β-triol [10] | 4.22 | NM | 3.19 | 3.79 |
| 3β,5α,6β,26-tetrol [1,10] | 0.00 | NM | NM | NM |
| 3β,5α,6β-triHCa [1,10] | 0.00 | NM | NM | 0.56 |
| 3β,5α,6β,24-tetraHCa [1] | 0.00 | NM | NM | NM |
| 3β,5α,6β-triHBA [1] | 0.00 | NM | 0.13 | 0.20 |
| 25-HC | 1.52 | 1.10 | 1.09 | 1.10 |
| 7α,25-diHC | 0.10 | NM | 0.50 | 0.54 |
| 7α,25-diHCO | 1.66 | 1.92 | 2.12 | 2.21 |
| 3β,7α,25-triHCA | 0.00 | NM | NM | NM |
| 7α,25-diH,3O-CA | 0.80 | NM | NM | NM |
| 3β,25-diHCA) [1] | 8.69 | NM | 4.34 | 4.66 |
| 3β,7β,25-triHCA [1] | 0.00 | NM | NM | NM |
| 24S-diHC | 9.61 | 9.19 | 9.64 | 9.56 |
| 24S,25-EC [3] | 3.02 | NM | 0.84 | 0.78 |
| 22R-HC | 0.00 | NM | NM | NM |
| 24R-HC | DNQ | NM | 0.99 | 1.44 |
| 7α,24-diHC | 0.00 | NM | NM | NM |
| 7α,24-diHCO | 0.00 | NM | NM | NM |
| 22H,24O-C [1,4] | 12.94 | NM | 10.36 | 11.06 |
| 3β,22-diHCA [1,5] | 5.07 | NM | 3.49 | 3.27 |
| 3β,x-diHCA [1,6] | 0.83 | NM | NM | NM |
| 22,25-diH,24O-C [1,7] | 7.07 | NM | NM | NM |
| 3β,x-diHCA [1,8] | 0.00 | NM | NM | NM |
| 3β,7α,x-triHCA [1] | 0.39 | NM | NM | NM |
| 7α,x-diH,3O-CA [1] | 1.77 | NM | NM | |

Abbreviations. DNQ, detected but not quantified. NM, not measured.
Dates in day/month/year format.
[1]. Identification based on retention time, accurate mass and MS$^{3}$ but in the absence of authentic standards.
[2]. 25R- and 25S-epimers measured in combination.
[3]. 24S,25-EC isomerises to 24-oxocholesterol during the sample preparation process.
[4]. Alternative structures 3β,20-dihydroxycholest-5-en-22-one (20H,22O-C).
[5]. Alternative structures 3β,x,y-trihydroxycholest-5-en-z-one (x,y-diH,zO-C).
[6]. Alternative structures 3β,x,y-trihydroxycholest-5-en-z-one (x,y-diH,zO-C).
[7]. Alternative structures 3β,x-dihydroxycholest-5-en-26-oic acid (3β,x-diHCA).
[8]. Alternative structures 3β,x,y-dihydroxycholest-5-en-z-one (x,y-diH,zO-C).
[9]. 7αH,24O-27-nor-C and 7αH-27-nor-C-3,24-diO are decarboxylation prpducts of 3β,7α-diH,24O-CA and 7αH,3,24-diO-CA, respectively.
[10]. Observed as the $[M-H_2O]^+$ ion.
[11]. The 26-hydroxy group is often refered to as a 27-hydroxy. We prefer the systematic nomenclature where 26 preceeds 27.
[12]. NIST SRM 1950 analysed in this study.
[13]. NISR SRM 1950 measurement from Yutuc et al Anal Chim Acta. 2021 Apr 15;1154:338259. The internal standard mix in [12] and [13] are different.
[14]. NIST SRM 1950 from measured made in 2024 (data accessed 01/11/2024). The internal standard mix in [14] and [15] are the same but differ from those used in [12] and [13].
[15]. NIST SRM 1950 from measurements made 2024 - 2025 (data accessed 24/06/2025).
